# Autistic traits and alcohol consumption through adolescence and young adulthood

**DOI:** 10.1101/2025.11.24.25340869

**Authors:** Stephanie Page, Richard Parker, Kayleigh E Easey, Felicity Sedgewick, Dheeraj Rai, Evie Stergiakouli

**Author notes:** **Corresponding author**: Stephanie Page **Email**.

## Abstract

**Background:** Contrary to previous findings, emerging studies support the increased likelihood of hazardous alcohol consumption in autistic adults when compared to neurotypical individuals. However, support services specifically for autistic people drinking hazardously are lacking. Examining the relationship of autistic features with alcohol consumption patterns longitudinally is essential to identify appropriate developmental points to offer support.

**Methods:** We investigated this using both phenotypic and genetic exposures measured in participants from the Avon Longitudinal Study of Parents and Children (ALSPAC). Our exposures included three measures of autistic traits (a broad autism phenotype measure, Social Communication Disorders Checklist score and polygenic score reflecting genetic liability for autism). Our outcome was alcohol consumption, measured across five timepoints between the ages of 17 to 28 years. We used multilevel piecewise linear spline analyses to model both the mean and 10^th^, 50^th^ (median) and 90^th^ quantiles of alcohol consumption longitudinally.

**Results:** There was little evidence that genetic liability for autism influenced alcohol consumption. However, we found that individuals with higher autistic traits drank less at the mean and 10^th^, 50^th^ (median) and 90^th^ percentiles of alcohol consumption compared to those with lower autistic traits. We did not find evidence of a relationship between social communication differences and alcohol consumption at the mean, 10^th^ and 50^th^ (median) percentiles. Conversely, there was evidence to suggest that individuals with higher autistic traits drank more at the 90^th^ percentile compared to those with fewer social communication differences.

**Conclusion:** Our findings indicate that social communication differences are associated with increased alcohol consumption in heavy drinkers, and that the relationship between autistic traits and alcohol consumption varies dependent on how traits are measured. This may explain the plurality of previous findings. Further research is needed to develop a more nuanced understanding of these associations across subpopulations of autistic traits and alcohol consumption.

## Introduction

Research on alcohol consumption in the autistic community has yielded conflicting findings. There is evidence to suggest that autistic adults are less likely to drink alcohol and develop a dependence compared to their neurotypical counterparts.^1–9^ For example, in a nationwide Swedish cohort of twins (N=20,863), with data collected at different timepoints (15, 18 and 24 years), individuals with a proxy diagnosis of autism were less likely to drink hazardously than twins without a diagnosis, and were less likely to drink hazardously than twins who had the proxy diagnosis and co-occurring attention deficit hyperactivity disorder and/or learning difficulties.^10^

However, in a sample of 123,543 Swedish adults, autistic individuals (N=26,986) without diagnosed intellectual disability (ID) were twice as likely to experience substance use related problems (including alcohol) as their non-autistic relatives.^11^ The risk was even greater in autistic individuals with co-occurring ADHD (which is unsurprising given that people with ADHD have higher rates of substance use when compared with both autistic people and the general population.^12,13^ Further, in a community sample of 237 autistic adults, both non-drinkers and hazardous drinkers had higher levels of autistic traits.^14^ Similar to the general population, non-drinkers and hazardous drinkers also reported higher levels of depression, generalised anxiety and social anxiety. This indicates a potential role of mental health difficulties in alcohol consumption in autistic adults.^15^ Depression has also been found to be associated with increased drinking frequency within the past year for autistic people between the ages of 16 and 20 years.^16^ Given that depression is more prevalent in autistic people than non-autistic people generally, it is possible that depression may mediate the effect of autism on increased alcohol consumption.^17^ Though research is limited, the emerging evidence that alcohol consumption could be problematic for some autistic people warrants further investigation. It also suggests that it is important to explore associations across the distribution of alcohol consumption, rather than assuming that associations in moderate drinkers are necessarily the same as those at the more extreme ends. Support for autistic individuals using alcohol is scarce, yet there is evidence that functional outcomes for autistic people with substance use disorders are worse than for non-autistic individuals, including domestic housekeeping and participation in society.^13^ Thus, further research in this area is crucial if appropriate support is to be made available.

Studies conducted using samples from the United States (US) suggest that alcohol consumption tends to start in adolescence, peak between the ages of 18 and 22 and decline by the mid-20s.^18^ Sex differences in alcohol consumption over the course of a decade have also been reported in the general US population, with males reporting higher levels of hazardous drinking compared to females.^19^ Existing studies on the relationship between alcohol consumption and autistic features are limited by cross-sectional study designs that are unable to capture the potential variability of alcohol consumption from adolescence into adulthood. Exploring this relationship longitudinally would allow us to examine the temporal sequence of autistic traits, potential risk factors and alcohol consumption. Further, it could also identify opportunities for early prevention strategies by identifying potentially modifiable pathways.

Observational studies can be prone to bias from unmeasured confounding, sample selection or measurement error, meaning any uncovered associations may not reflect direct causal relationships.^20^ Triangulation can help ascertain how robust findings are to such limitations by incorporating evidence from methods with differing sources of potential bias, such as prospective cohort studies and methods from genetic epidemiology.^21,22^ Consistent findings when drawing upon combinations such as these may provide reassurance that an any observed effects are therefore less likely to be the result of potential biases, making any inferences more reliable. The aim of this study was there to investigate if alcohol consumption was associated with autistic traits in the general population between the ages of 17 to 28 years, at different percentiles of alcohol use, using phenotypic and genomic data to triangulate findings.

## Methods

### Participants

Participants were sourced from the Avon Longitudinal Study of Parents and Children (ALSPAC), a prospective longitudinal birth cohort study based in the UK.^23,24^ ALSPAC recruited 14,203 pregnant women based in Avon (in south-west England) with expected delivery dates between April 1, 1991 and December 31, 1992. Some women had multiple pregnancies, meaning the initial number of pregnancies enrolled was 14,541. N=13,988 children from these pregnancies were alive at one year of age. This was then increased to 15,447 pregnancies by enrolment initiatives.^25^ Of these children, 14,901 children were alive at one year of age. This study focused on the children of the pregnant women involved (known as Generation 1 (G1) of ALSPAC) but also used measures from mothers and partners from G0. Following additional phases of recruitment, 14,833 mothers were enrolled in ALSPAC as of September 2021. N=12,113 G0 partners have been in contact with the study (and 3,807 G0 partners are currently involved).^26^ As stated by the ALSPAC team, “Please note that the study website contains details of all the data that is available through a fully searchable data dictionary and variable search tool. Ethical approval for the study was obtained from the ALSPAC Ethics and Law Committee and the Local Research Ethics Committees. Informed consent for the use of all data collected was obtained from participants following the recommendations of the ALSPAC Ethics and Law Committee at the time. Participants can contact the study team at any time to retrospectively withdraw consent for their data to be used. Study participation is voluntary and during all data collection sweeps, information was provided on the intended use of data. The completion of a questionnaire, either on paper or online, was considered to be written consent from participants to use their data for research purposes. The informed consent obtained from ALSPAC (Avon Longitudinal Study of Parents and Children) participants does not allow the data to be made available through any third party maintained public repository. Supporting data are available from ALSPAC on request under the approved proposal number, B4193. Full instructions for applying for data access can be found here: http://www.bristol.ac.uk/alspac/researchers/access/. The ALSPAC study website contains details of all available data (http://www.bristol.ac.uk/alspac/researchers/our-data/).” ^27,28^

This study also used ALSPAC genetic data for G1. G1 were genotyped using the Illumina HumanHap550 quad chip genotyping platforms by 23andme, imputed to HapMap2 and subjected to standard quality control methods (the details of which can be found in the Supplementary Methods).^29^

### Measures

Measures were collected for G1 via self and parent-reported postal questionnaires. The timeline for data collection is given in Figure 1.

**Figure 1.**
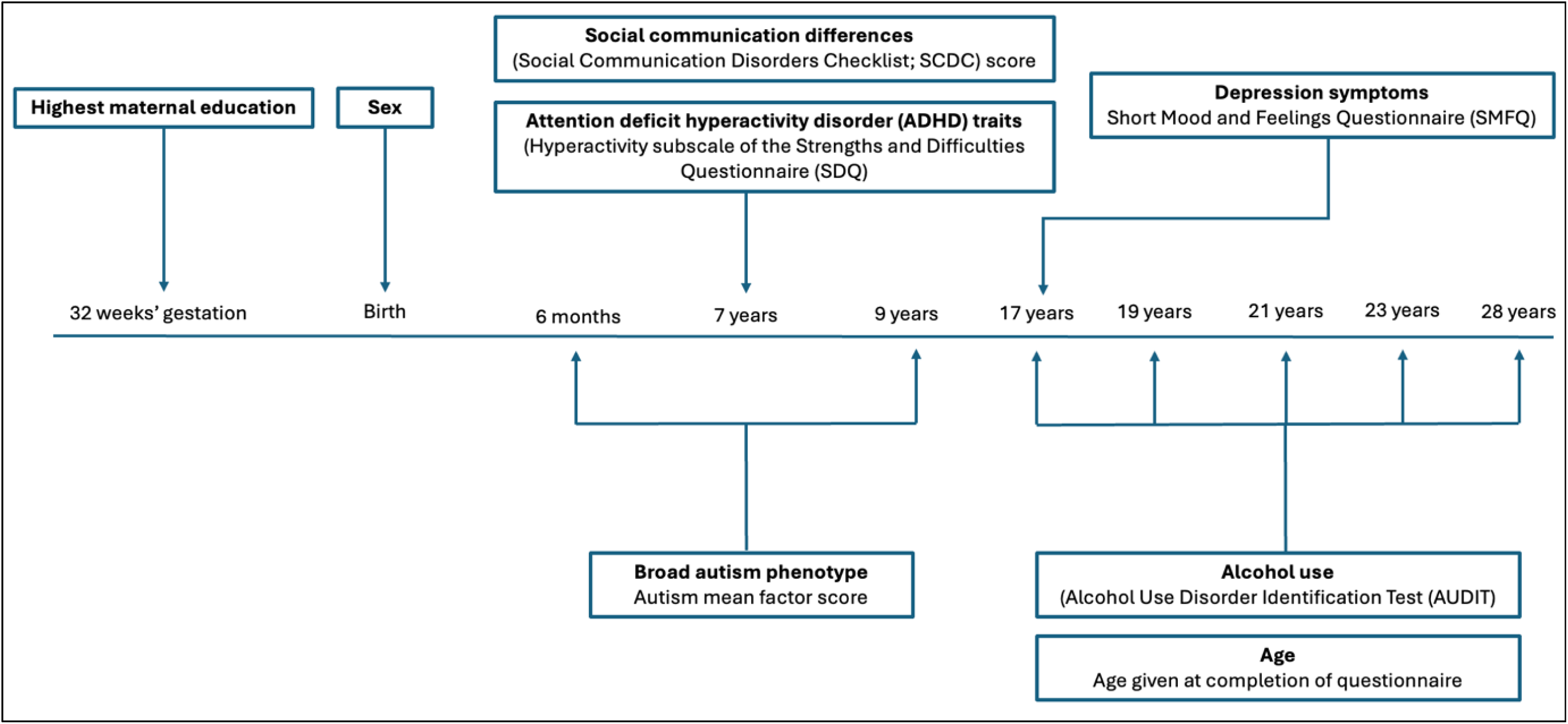
Timeline of data collection for main model variables.

### Exposures

Three different measures of autistic traits were analysed, independently, to triangulate our findings and capture different facets of autism. The analytical datasets consisted of participants with observed data for all the covariates described below, as well as an observation of the outcome on at least one occasion and an observation of the specific exposure included in that model. Given participants did not always have observed data for all three exposures, the sample sizes differed between models for each exposure. Details of how these samples were derived and corresponding sample sizes are given in Figure 2 and Supplementary Figures S40 and S41.

**Figure 2.**
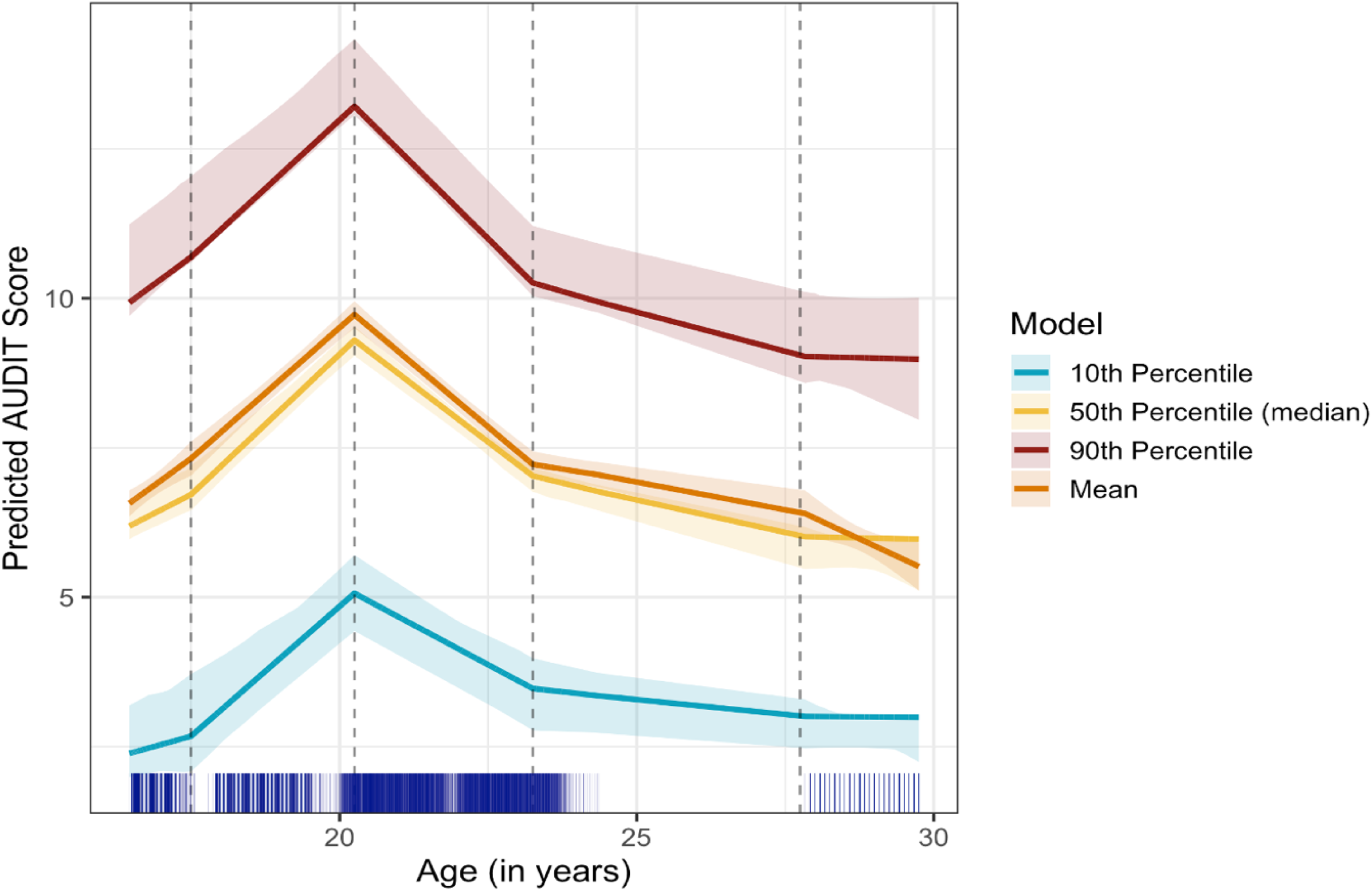
Trajectories of mean, 10^th^, 50^th^ (median) and 90th alcohol consumption between approximate ages 17 to 28 years in a sample where participants had complete data for all variables and at least one outcome timepoint (N=3,057). Vertical dashed lines indicate knot positions in the model. A rug is included underneath to show observation frequency.

The first exposure was a score derived from parent responses to the Social Communication Disorders Checklist (SCDC), a 12-item measure for autistic traits that was administered when the child was approximately seven years old.^30^ Scores range between 0-24, and a score of nine or more has shown good discriminant validity for pervasive developmental disorders in children and adolescents.^31^ Though it has not yet been validated in adults, the SCDC has been found to have similar neurodevelopmental and genetic correlates at 25 years to the SCDC measured at approximately age 11.^32^ The sample size where individuals had data for the SCDC variable, relevant covariates and at least one outcome timepoint was N=3,771.

Given that social communication differences are only one facet of autism, an additional phenotypic measure was used to reflect the broader autism phenotype. This was a mean factor score of 93 (predominantly parent-reported) traits associated with autism, collected between the first six months and nine years of the child’s life.^33^ A lower (more negative) score reflected higher levels of autistic traits (and vice versa). The sample size where individuals had data for the autism mean factor score, relevant covariates and at least one outcome timepoint was N=4,035.

Finally, a polygenic score for autism was calculated using ALSPAC genotypic data for G1. The raw genome-wide data were subject to standard quality control methods cited in the Supplementary Methods (S1). The risk alleles associated with autism that were included in the polygenic score were derived from a genome wide association study (GWAS) of autism deemed to be the most appropriate based on size.^34^ Only single nucleotide polymorphisms (SNPs) present in both the GWAS summary statistics and ALSPAC genotypic data were used in the calculation. Data were clumped to ensure independence (r^2^=0.01, kb=10,000, p=1). The effect sizes of these alleles were used as weights when calculating the sum of risk alleles for individuals in the target sample (ALSPAC G1) with 12 different p value thresholds, ranging from 0.5 to 1×10^-7^. The p-value threshold selected for the finalised autism-PRS used in analyses was 1×10^-5^. This was selected based on the outcome of regression analyses where the variance explained by each polygenic score (dependent on the p-value used during score calculation) was compared for both of our phenotypic autism measures (see Supplementary Tables S1-S2). To assist interpretability of model results, scores were standardised (details of which can be found in the Supplementary Methods S3). The polygenic scores for autism were calculated using the software R (version 4.3.2) and University of Bristol ACRC Blue Pebble with the PLINK package (version 1.90). The sample size where individuals had data for the polygenic score for autism, relevant covariates and at least one outcome timepoint was N=3,250.

### Outcomes

Measurements were taken at five different timepoints (approximately 17, 19, 21, 23 and 28 years old) through self-reported responses to the 10-item Alcohol consumption Disorders Identification Test (AUDIT) via postal questionnaires.^35^ A score between eight and 15 was taken to indicate hazardous or harmful alcohol consumption, while a score of 15 suggested alcohol dependence.^36^ The AUDIT has been shown to be a valid measure for detecting problem drinking in both adolescents and adults.^37,38^

### Covariates

Sex was included as a potential confounder and measured as sex recorded at birth. Socioeconomic status (SES) was also included as a potential confounder through a proxy measure of maternal education at 32 weeks’ gestation. Data on SES was also measured through a collection of parental variables (namely maternal occupational social class, homeownership status, parity, maternal marital status, age at delivery and three binary measures of maternal smoking pre and during pregnancy). Further details of these additional SES measures can be found under Supplementary Methods S8.

A score reflecting attention deficit hyperactivity disorder (ADHD) traits was included as a covariate of interest. Traits of ADHD were measured via parent responses to the Hyperactivity subscale of the Strengths and Difficulties Questionnaire (SDQ) at seven years of age.^14^ A score of six or more was taken to suggest elevated ADHD symptoms.^36^ The SDQ has shown good discriminant validity for both children and adults with ADHD vs. controls. ^39,40^

As autistic adults engaging in either hazardous drinking or teetotalism have reported higher levels of depression symptoms, a measure of depression symptoms was also included as a baseline covariate.^14^ Depression symptoms were derived from self-reported responses to the Short Mood and Feelings Questionnaire (SMFQ) at approximately age 17 years.^41^ A score of eight or more was taken to indicate depression. The SMFQ is a recommended screening tool for depression in children and adolescents and has been validated in young adults.^42^

Participants were asked to record their age when completing the questionnaires. Age was then centred on the lowest timepoint (17 years) to assist with results interpretation.

### Statistical analysis

A series of multilevel models, with measurement occasion nested within participants, were fitted to the data. The Akaike Information Criterion (AIC) was used to guide model selection, where a lower score (difference of 3 or more) was taken to indicate better model fit.^43^ See Supplementary Methods S4 for a detailed description of how models were derived.

Our final baseline model had both a random intercept and a random slope for age (as a linear term across its full range, centred at 17 years). It also included random intercepts and slopes for age across measurement occasions to allow for complex level 1 variation. The fixed part of the model included age as piecewise linear splines (with knots at years 17½, 20¼, 23¼ and 27½). It also included sex, maternal education, depression traits, ADHD traits, and two-way interactions between age and each of sex, depression, and ADHD. Each autism measure was then added separately, to explore the association between autistic traits – as measured by these complementary, but correlated, indices – and alcohol consumption over time. Given depression might be a mediator on any pathway between autistic traits and alcohol consumption, the estimated effect of autistic traits on the outcome may reflect its direct effect (excluding that mediated by depression), as opposed to total effect, when adjusting for depression. We therefore also fitted the same final baseline models, but with depression, and its interactions, excluded to check the sensitivity of the autistic trait effect estimates to its inclusion.^44^ Further, to investigate the potential for a non-linear relationship between each autism measure and alcohol consumption, the model fit of the final baseline model for each autism exposure was compared against the same model, but with the addition of interactions between the exposure and age splines. These analyses were conducted in R (version 4.3.2), with splines derived via lspline (version 1.0-0).^45,46^ We modelled mean alcohol consumption using the R2MLwiN package (version 0.8-8), which calls MLwiN (version 3.00).^47,48^ To explore alcohol consumption at the 10^th^, 50^th^ (median) and 90^th^ percentiles, we used linear quantile mixed modelling (LQMM) via the R package lqmm (version 1.5.8), using the same splines as when modelling the mean.^49^ Further details of these analyses, in addition to information on how we checked model assumptions, are available in the Supplementary Methods.

### Mediation analyses

Due to the possibility of depression lying on the causal pathway between autism and alcohol consumption, a counterfactual mediation framework was used to explore the potential mediating effect of depression symptoms on any such pathway.^50,51^ Individual effects are defined and further explained visually using directed acyclic graphs (DAGs) in Supplementary Table S3 and Supplementary Figures S1-S4. These analyses were carried out using Paramed, a user-written program for Stata (version 2.3.1. and version 18.0).^52,53^

## Results

Details of how the analytical samples were determined are given in Figure 1. Baseline descriptive statistics are given for the analytical sample with the greatest sample size (autism mean factor score; N=4,035) in Table 1. Equivalent tables for the SCDC score and polygenic score for autism are available in Supplementary Tables S44 and S45.

**Table 1.** Descriptive statistics of the analytical sample with the largest sample size (where the autism mean factor score is the exposure, N=4,035). Please note that descriptive statistics for the other two analytical samples can be found in Supplementary Tables S44 and S45.

| Variable | N (%) |
| --- | --- |
| **Sex** |  |
| *Males* | 1,682 (41.69) |
| *Females* | 2,353 (58.31) |
| **Maternal highest education 32 weeks' gestation** |  |
| *Up to A level* | 2195 (72.24) |
| *Degree* | 849 (21.04) |
| *Vocational* | 271 (6.72) |
| **Maternal home ownership status** |  |
| *Mortgaged or owned* | 3,428 (86.59) |
| Rented | 605 (15.29) |
| **Parity** |  |
| *0* | 1,943 (48.94) |
| *1* | 1,399 (35.24) |
| *2* | 479 (12.07) |
| *3+* | 149 (3.76) |
| **Maternal occupational social class** |  |
| Professional occupation | 321 (9.01) |
| Managerial or technical occupation | 1,340 (37.60) |
| Skilled occupation (manual or non-manual) | 1,611 (45.21) |
| Partly skilled occupation, unskilled occupation or armed forces | 292 (8.19) |
| **Maternal marital status** |  |
| Never married | 465 (11.65) |
| Divorced, separated or widowed | 177 (4.43) |
| First marriage | 3,092 (77.44) |
| Marriage two or three | 259 (6.49) |

Flowcharts detailing how each of the three analytical samples was determined are available in Supplementary Figures S40-42. Completeness/missingness for each outcome timepoint in each of the three samples is detailed in Supplementary Table S43.

### AUDIT trajectories

The unadjusted predicted mean, 10^th^, 50^th^ (median) and 90^th^ percentiles of alcohol consumption measured through AUDIT are represented in Figure 2. Details of how this model was derived are available in the Supplementary Methods S9.

As can be seen in Figure 2, across all percentiles and the mean, the steepest increase in alcohol consumption is in the second slope (between knots at ages 17½ years and 20¼ years). Alcohol consumption then peaks at around the age of 20¼ years and begins its steepest decline in the third slope between the ages of 20¼ years and 23¼ years. The decline following this is much shallower (between the ages of 23¼ years and 27½ years); however, it should be noted that there is a gap in measurement of five years between ages 23 and 28 years, so it is possible there is an additional pattern that is unobserved here. Mean alcohol consumption is then predicted to further decline after 27½ years, whilst for the 10^th^, 50^th^ (median) and 90^th^ percentiles it tends to plateau. AUDIT scores by age at measurement are also reported for the mean, 10^th^, 50^th^ (median) and 90^th^ percentiles in Supplementary Tables S4-6.

### Sex differences

Mean alcohol consumption over time is visualised in males and females separately in Supplementary Figure S5. AUDIT scores by age at measurement are also reported for the mean, 10^th^ and 90^th^ percentiles in males (Supplementary Table S5) and females (Supplementary Table S6). As can be seen in Supplementary Figure S5, there is a difference between alcohol consumption in males and females over time. Males’ alcohol consumption rises more steeply, on average, across the mid-to-late teens, and whilst a difference in mean consumption persists thereafter, the subsequent patterns of increase and decline are similar to those captured in females.

### Measures of autism and alcohol consumption trajectories

Comparing the fit of models with, and without, the inclusion of exposure*age interactions found no evidence that the association between the exposures and alcohol consumption changed across age (see Supplementary Tables S7 and S20). Thus, results from the original final baseline models (without such interactions) are reported in Table 2, with full estimates for each model given in Supplementary Tables S23-S26.

**Table 2.** Results of multilevel piecewise linear spline models of the mean, and also 10^th^, 50^th^, and 90^th^ quantiles, of the distribution of f-reported alcohol consumption (AUDIT), exploring its association with three measures of autism over time.

| Exposure | Mean or percentile of alcohol consumption across time | Estimated change in AUDIT score per 1-unit increase in exposure | CI 95% (lower, upper) |
| --- | --- | --- | --- |
| Mean factor score for autism | Mean | 1.05 | 0.67, 1.44 |
|  | 10<sup>th</sup> | 0.97 | 0.53, 1.40 |
|  | 50<sup>th</sup> (median) | 0.98 | 0.56, 1.41 |
|  | 90<sup>th</sup> | 0.98 | 0.61, 1.36 |
| Social Communication Disorders Checklist (SCDC) score at 7y | Mean | -0.01 | -0.05, 0.03 |
|  | 10<sup>th</sup> | -0.02 | -0.06, 0.02 |
|  | 50<sup>th</sup> (median) | -0.01 | -0.07, 0.05 |
|  | 90<sup>th</sup> | 0.09 | 0.02, 0.16 |
| Polygenic score for autism | Mean | -0.13 | -1.10, 0.84 |
|  | 10<sup>th</sup> | 0.06 | -0.95, 1.06 |
|  | 50<sup>th</sup> (median) | 0.06 | -0.76, 0.89 |
|  | 90<sup>th</sup> | 0.06 | -1.09, 1.22 |

Models were adjusted for sex, socioeconomic position, depression symptoms and attention deficit hyperactivity disorder (ADHD) traits.

### Autism mean factor score

As outlined above, for the autism mean factor score, lower (more negative) scores indicated higher levels of autistic traits, while higher scores indicated lower autistic traits. On average, the AUDIT score across the mean and the 10^th^, 50^th^ (median) and 90^th^ percentiles was predicted to be higher per 1-unit increase in the autism mean factor score, i.e. people with higher autistic traits reported consuming less alcohol than people with lower autistic traits (β (95%CI) 1.05 (0.67, 1.44); 0.97 (0.53, 1.40); 0.98 (0.56; 1.41); 0.98 (0.61, 1.36)). These estimates were slightly smaller, but substantively similar, when excluding depression from the models (see Supplementary Table S8).

### Social Communication Disorders Checklist (SCDC) score

There was no evidence that the AUDIT score across the mean, 10^th^ and 50^th^ (median) percentiles differed, on average, across levels of social communication difficulties (per 1-unit increase in SCDC score, AUDIT was predicted to change by: β (95%CI) –0.01 (–0.05, 0.03); –0.02 (–0.06, 0.02); –0.01 (–0.07, 0.05); respectively). Conversely, there was evidence indicating that, on average, the AUDIT score for the 90^th^ percentile was higher in those with greater social communication difficulties, compared to those with fewer social communication difficulties (β (95%CI) 0.09 (0.02, 0.16)). These substantive findings persisted when excluding depression from the models: i.e. there was no evidence of a change in the mean, 10^th^ or 50^th^ (median) percentiles of AUDIT across SCDC score, but there was evidence that the AUDIT score was higher at the 90^th^ percentile for those with greater social communication difficulties compared to those with fewer social communication difficulties. This estimate was greater than when depression was included in the model (see Supplementary Table S8).

### Polygenic scores

There was no evidence that, on average, the mean, 10^th^, 50^th^ (median) and 90^th^ percentiles of the AUDIT score differed across genetic liability for autism (per 1-unit increase in the polygenic score, AUDIT was estimated to change by: β (95%CI) –0.13 (–1.10, 0.84); 0.06 (–0.95, 1.06); 0.06 (–0.76; 0.89); 0.06 (–1.09; 1.22); respectively). There was similarly no evidence that genetic liability for autism was associated with AUDIT scores, at the mean, 10^th^, 50^th^, and 90^th^ percentiles, when excluding depression from the model (see Supplementary Table S8).

### Mediation analyses

Counterfactual mediation analyses did not reveal evidence for mediating effects of depression symptoms on the relationship between autism mean factor score, SCDC score or polygenic score and alcohol consumption (please see Supplementary Tables S9-S17 for results and S3 for effect definitions, also visualised using directed acyclic graphs in Supplementary Figures S1-S4).

### Missing data

It was beyond the scope of this study to address missing data via multiple imputation (and related) methods. This is due to the complexity of both patterns of missingness and statistical modelling used, and a lack of capacity for existing software to accommodate model complexity. However, chi squared comparisons showed that participants who were missing outcome data were more likely to report lower maternal education than those who had the outcome observed on at least one measurement occasion, as well as baseline covariates up to the age of 17 (Pearson *χ*^2^(4) = 687.49, p = <0.001). Individuals who had missing data in baseline covariates up to the age of 17 were also more likely to report lower maternal education than those who had complete data for this (Pearson *χ*^2^(4) = *688.94*, p = <0.001). Finally, males were more likely to have missing outcome data (Pearson *χ*^2^(1) = *112.47*, p = <0.001) as well as missing data in baseline covariates up to the age of 17 (Pearson *χ*^2^(1) = *101.85*, p = <0.001) (see Supplementary Tables S39-42).

### Sensitivity analyses

Model assumptions were tested and were not found to be violated for either multilevel linear or linear quantile mixed models (the details of which can be found in Supplementary Methods S5 and S7).

## Discussion

This is the first longitudinal study to examine the relationship between autistic traits and alcohol consumption across adolescence and young adulthood in a large cohort sample. Whilst there was no evidence to suggest the association of autistic traits with alcohol consumption changed across this age range, some exposures did have time-invariant effects. There was evidence to suggest that those with lower autistic traits drank at an increased rate compared to those with higher autistic traits. However, there was also evidence to suggest that heavy drinkers drank even more heavily when they had social communication difficulties compared to those with fewer social communication difficulties. There was no evidence to suggest a relationship between higher genetic liability for autism and alcohol consumption across time.

We found that individuals with higher autistic traits (indicated by a lower mean autism factor score) drank less than those with lower autistic traits across the mean and all three percentiles. Indeed, previous studies have also found that autistic people tend to drink less alcohol.^1–9^ However, the finding that individuals with greater social communication difficulties drank more heavily at the 90^th^ percentile of alcohol consumption aligns with previous findings.^14^ The autism mean factor score was chosen as an exposure to reflect the broad autism phenotype, while the SCDC score (a measure of social communication difficulties), represents an individual facet of autism that is potentially more relevant to alcohol consumption due to its social nature. It is therefore possible that the social nature of drinking alcohol drives individuals with social communication difficulties to drink more heavily than their neurotypical counterparts when drinking with others. Indeed, autistic adults have cited improvements in communication as motivation for drinking in an online survey.^54^ However, it should also be noted that social communication difficulties are not exclusively seen in autism; other conditions, such as ADHD, depression and social anxiety, are associated with greater social communication difficulties compared to those without such disorders.^55–58^ Further, our findings suggest that social communication difficulties specifically, rather than depression, can influence heavy alcohol consumption in those with autistic traits (given the lack of evidence to suggest that depression symptoms mediated the effect between any of the autism exposures and alcohol consumption over time). This does not align the finding that autistic adults who drink hazardously report higher levels of depression symptoms than those who do not drink hazardously, further highlighting the plurality of findings in this area.^14^ Additional research is needed to explore motivations such as these in more depth given the diversity of the autistic community. A pragmatic approach to this could be to draw upon qualitative research methods, capturing rich experiential data that quantitative methods cannot and therefore uncover further nuances around the complexity of alcohol consumption in the autistic community.^59^

Autism is highly heritable.^60^ Although genetic liability is not deterministic, there is evidence to suggest that genetic liability (quantified through polygenic scores) for autism is strongly associated with both autism diagnoses and autistic traits.^61^ However, polygenic risk scores currently explain only a small proportion of variance in autism and autistic traits.^62–64^ This could therefore explain why we did not detect any associations of genetic liability to autism with alcohol consumption in this sample.

These study findings reflect the complexity of existing evidence. Many members of the autistic community may be less likely to drink hazardously than their neurotypical counterparts, while others may be at a comparatively increased risk of problematic alcohol consumption, especially those with higher social communication difficulties.^1–9, 11, 14^ Our findings for the SCDC also support the association between autistic traits and alcohol consumption in adolescence as well as adulthood, which is contradictory to previous findings suggesting that autistic adolescents are not at risk of problematic alcohol consumption.^62–64^

This study has several strengths. It is the first study to model the relationship between autistic traits and alcohol consumption from adolescence into young adulthood longitudinally. It also draws upon measures that not only capture differing facets of autism, but involve divergent sources of potential bias, strengthening findings through triangulation.^65^ Further, by modelling alcohol consumption at the mean and 10^th^, 50^th^ (median) and 90^th^ percentiles across time, we were able to capture the potential complexity of the relationship between autistic traits and alcohol consumption across its full spectrum. This may provide insight into apparently contradictory findings from previous studies (which is arguably expected given the diversity of the autistic community generally). In addition, we were able to test for age-varying effects of the exposures on alcohol consumption. We did not find evidence for such effects, and so we were unable to identify timepoints which might be best suited to intervention. However, this does not mean that such effects do not exist. Measuring across an expanded age range, or more frequently, or using alternative measures of autistic traits may detect any such effect, should it exist.

The study also has limitations. Firstly, due to the larger number of females compared to male participants and higher socioeconomic status of the ALSPAC cohort, the findings of this study are at risk of selection bias – i.e. participants may not represent a random sample of the target population – and therefore selection bias cannot be ruled out ^66^. As well as selection into the cohort, subsequent attrition may introduce further selection bias, and indeed we found that those who had lower maternal education, and were male, were less likely to have complete data. However, due to the complexity of patterns of missingness, the statistical modelling used, and the lack of capacity for existing software to accommodate model complexity, it was beyond the scope of this study to address missing data via multiple imputation, and related, methods. Therefore, results are based on observed data analyses, including complete covariate data and all observed outcome measures for each participant. This may be unbiased if completeness does not depend on the outcome, but potentially biased if data are not missing at random (MAR) or are missing in variables other than the outcome.^67,68^ Similarly, the use of a European sample for the calculation of the polygenic score for autism hinders generalisability to non-European populations. Moreover, the self-reported nature of the alcohol measures used may be biased by social desirability, meaning that certain participants may be less likely to respond honestly if they are embarrassed to be misaligned with perceived societal norms.^69^ Finally, there is limited data available in ALSPAC to measure intellectual disability (ID) and this was therefore not taken into account in this study. Although a multisource approach has been used to identify cases of probable ID, the total number of participants this applies to remains low.^70^ A measure of IQ could have been included as a proxy measure for this analysis (though IQ itself is not the sole determinant of an ID diagnosis).^71^ It is possible that results are biased as individuals with social communication difficulties and co-occurring ID may be less likely to drink heavily than those without co-occurring ID.^11^

Whilst we found that higher autistic traits were associated with lower levels of alcohol consumption, our results also indicated that greater social communication difficulties were associated with higher levels of problematic drinking. As such, this study highlights the complexity of the relationship between autistic traits and drinking alcohol, as reflected in past, and sometimes contradictory, findings. Given the relative paucity of research in this area, however, further work will help better understand this complexity: for example, studying cohorts subject to different selection pressures; investigating measures of autism which may be sensitive to different facets; measuring co-occurring ID; sampling across different age ranges and/or more frequently; and using genomic data from non-European ancestries. Addressing these may help cultivate a sample that better represents the population as a whole and therefore improve generalisability of findings. Qualitative methods will also provide more contextual information regarding the nuances of this area. Given the heterogeneous presentation of the autistic community, motivations for drinking heavily are likely to vary, and it is important for research to acknowledge this diversity. As such, members from the autistic community whose health and quality of life are negatively impacted by alcohol consumption may be unwittingly neglected by existing health programmes. Better understanding of the relationship between autism and alcohol consumption will enable suitable support to be designed and targeted.

## Data Availability

The informed consent obtained from ALSPAC (Avon Longitudinal Study of Parents and Children) participants does not allow the data to be made available through any third party maintained public repository. Supporting data are available from ALSPAC on request under the approved proposal number, B4193. Full instructions for applying for data access can be found here: http://www.bristol.ac.uk/alspac/researchers/access/. The ALSPAC study website contains details of all available data (http://www.bristol.ac.uk/alspac/researchers/our-data/).

## Acknowledgements

We are extremely grateful to all the families who took part in this study, the midwives for their help in recruiting them, and the whole ALSPAC team, which includes data collection staff, data and administrations staff, technical managers and the technical staff with the Bristol Bioresource Laboratory, based within the University of Bristol. We would also like to express our gratitude to Dr Elinor Curnow, Dr Gemma Hammerton, Professor Jon Heron, Dr Kate Birnie, Dr Ana Gonçalves Soares and Dr Paul Madley-Dowd for their contributions to this study.

