## Supplementary Methods for "Autistic traits and alcohol consumption through adolescence and young adulthood"

S1. Quality control measures for genomic data

ALSPAC genomic data

The ALSPAC team carried out quality control methods after G1 (ALSPAC children) had been subjected to genotyping using the Illumina HumanHap550 quad genotyping array. Specific details of these quality control measures are available at [ALSPAC OMICs Data Catalogue (bristol.ac.uk)](https://proposals.epi.bristol.ac.uk/alspac_omics_data_catalogue.html#org979f2a0).

Autism Genome-wide Association Study (GWAS) data (Grove at al. 2019)

Grove et al. (2019) genotyped a Danish cohort before performing quality control methods on the data, which are further described in the results section of their paper referenced at the end of this document.

S2. Selection of p-value threshold for finalised autism-PRS

Results of regression analyses in which the phenotypic variance explained by each polygenic score (where age, sex and principal components are controlled for) are included in Supplementary Tables S1 and S2. Principal components were derived for HRC reference panel data by adapting and running code from the ALSPAC genetics directory used to derive principal components for 1000 genomes data.

S3. Standardisation of autism polygenic scores

To ease interpretability of the effect estimates for the autism polygenic score within the final baseline models, polygenic scores were standardised. First, the mean and standard deviation were calculated for the polygenic score. The mean was then subtracted from each individual score, and this was then divided by the standard deviation. Each individual original score was then replaced with the standardised version ready for analyses.

S4. Deriving piecewise linear spline models

In the random slopes models, the coefficient of age, as a linear term across its full range, was allowed to randomly-vary across participant (as allowing spline terms to instead randomly vary across participant lead to estimation difficulties). In the fixed part of the model (estimating population effects), any non-linearity in the association of age with the AUDIT score was initially assessed by fitting restricted cubic splines with differing numbers of knots, as well as age as a single linear term only, and comparing model fit (Harrell, 2015). This was initially carried out in two datasets split by sex. Conditional on the same function of age describing its relationship with AUDIT equally well in both sexes, the sexes were then combined in the same model, with an age*sex interaction. As part of determining knot placement, a restricted cubic spline model was fitted with a loop to explore the most appropriate number of potential knot points. Comparisons were drawn between the AIC for the model with ten knots and the random slopes model. Ten knots were then compared with three knots, and this continued in ascending order of knots (four, five, etc) until a chi squared test showed no evidence of a statistical difference. The lowest number of knots in the comparison was then selected as the appropriate number of knots for the restricted cubic spline model (N = 8). The population trend was plotted for this model (see Supplementary Figure S41). Once a satisfactory function had been derived, the resulting predictions were used to guide knot placement in a piecewise linear spline model, which – contingent on the model fit being judged as similarly satisfactory – benefits from coefficient estimates which are more readily interpretable, compared to those from a restricted cubic spline model. We then fitted a series of linear piecewise spline models, basing the location of knots on visual inspection of this restricted cubic spline plot (Howe et al., 2013).

S5. Testing model assumptions for piecewise linear spline models

To test the model assumptions of our multilevel linear models (normality, homoscedasticity and independence of residuals), a series of scatter plots, density plots and QQ plots were created and visually inspected (Supplementary Figures S6-S18). The results of these model checks suggested non-violated assumptions of homoscedasticity and independence of residuals, but non-normally distributed level 1 and 2 residuals. As a sensitivity analysis with regard to the non-normality of residuals, the corresponding models were rerun after the outcome (alcohol use) had been log transformed. Model checks for these are given in Supplementary Figures S19-S27. When comparing the output of the main and log transformed models, there were no substantive differences (Supplementary Table S18) and so results modelling the original scale are presented in the main paper.

S6. Linear quantile mixed modelling

To ensure model convergence, we used a range of quadrature knots (nK = 4 to 15), while the maximum number of iterations was 2000 and the tolerance level was set to 1e-02 to accommodate model complexity.

S7. Testing model assumptions for linear quantile mixed models

To test the model assumptions of our multilevel linear quantile mixed models (homoscedasticity and independence of residuals), scatter plots were created and visually inspected (available in Supplementary Figures S31-S39). We were satisfied that the output of these models did not violate the assumptions of our linear quantile mixed models.

S8. Additional measures of socioeconomic status (SES)

Maternal occupational social class was measured through the categories ‘Professional occupation’, ‘Managerial or technical occupation’, ‘Skilled occupation (non-manual)’, Skilled occupation (manual)’, ‘Partly skilled occupation’ and ‘Unskilled occupation’. Homeownership status was recorded when the child was approximately 8 months old and consisted of the categories ‘Mortgaged’, ‘Owned’, ‘Council rented’, ‘Rents privately (furnished)’, ‘Rents privately (unfurnished)’, ‘House association rented’ or ‘Other’. Parity was recorded by measuring the number of pregnancies mothers had carried to term. Age at delivery was measured by the mother’s age in years at their first birth.

S9. Deriving the model for Figure 2

As in the substantive models, the coefficient of age, as a linear term across its full range, was allowed to randomly-vary across participant. In the fixed part of the model (estimating population effects), any non-linearity in the association of age with the AUDIT score was included as piecewise linear splines. For plot simplicity, all measures were excluded apart from AUDIT scores and age, so as to model the relationship between AUDIT and age over time.
