## Supplementary Figures for "Autistic traits and alcohol consumption through adolescence and young adulthood"

Counterfactual mediation analyses

*Supplementary Figure S1*. Directed acyclic graph (DAG) of controlled direct effect (CDE).

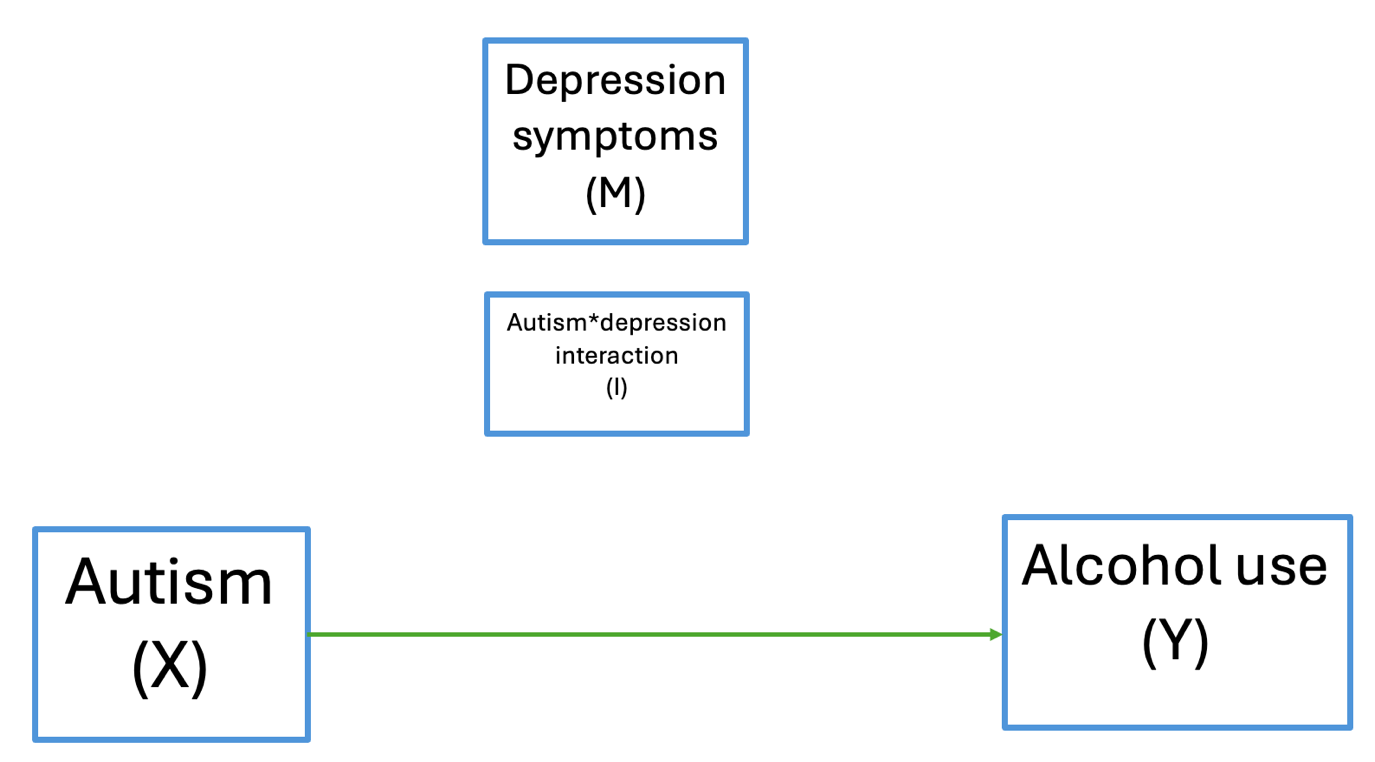

*Supplementary Figure S2*. Directed acyclic graph (DAG) of natural direct effect (NDE).

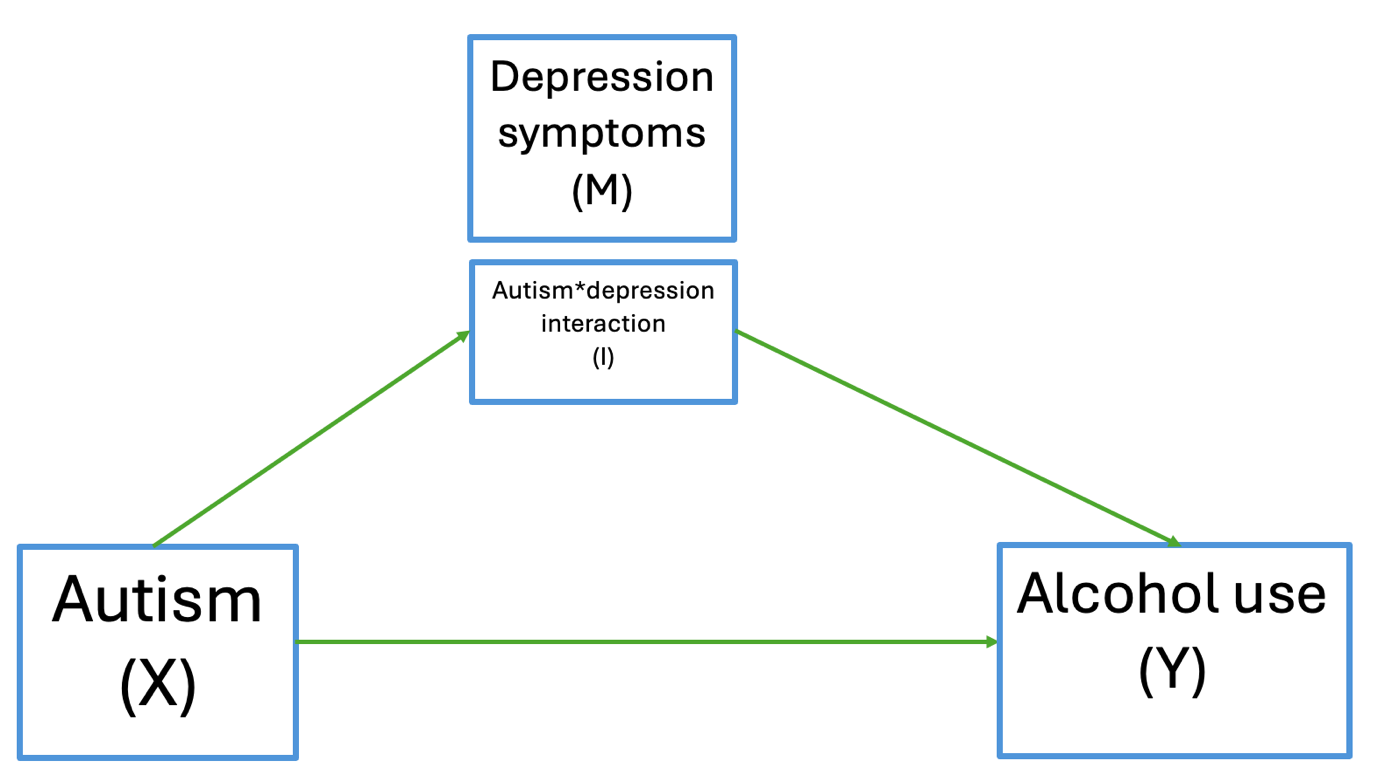

*Supplementary Figure S3*. Directed acyclic graph (DAG) of natural indirect effect (NIE).

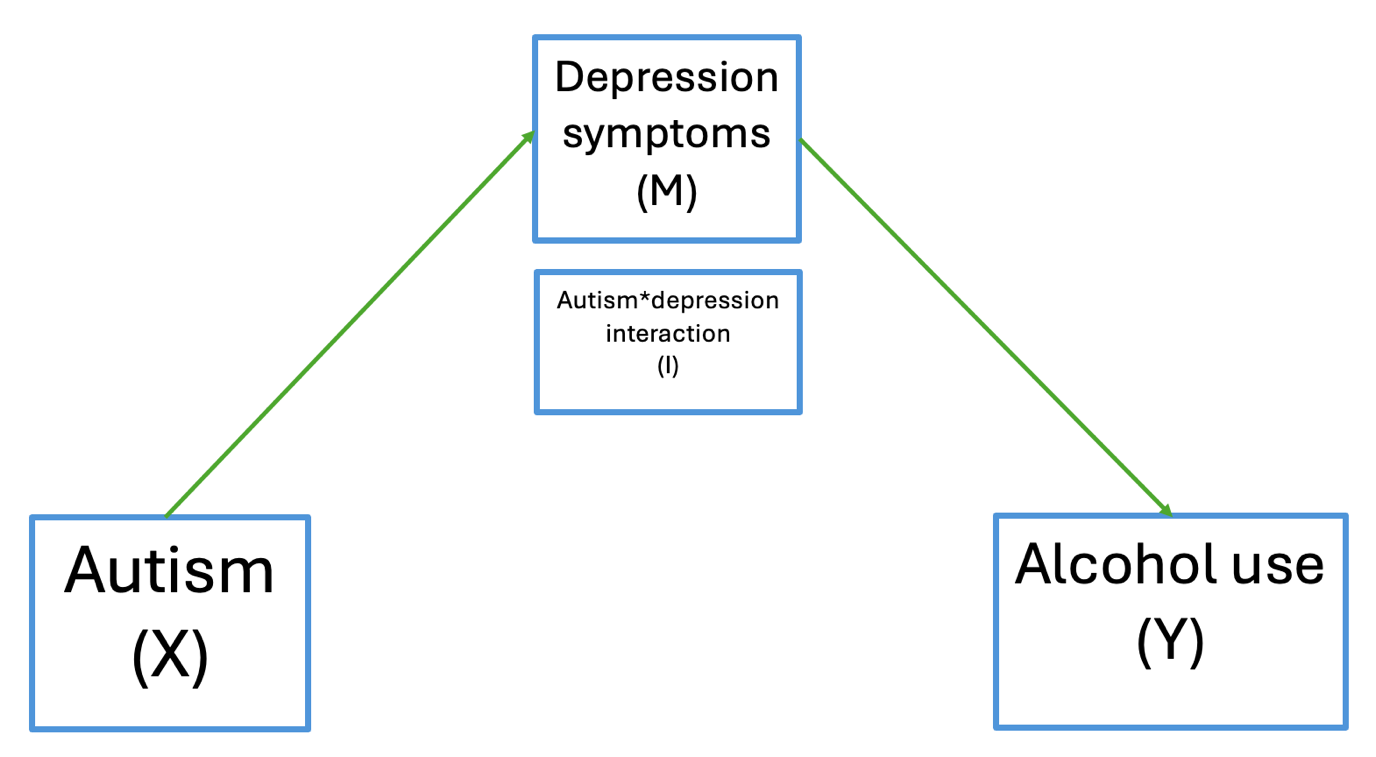

*Supplementary Figure S*4. Directed acyclic graph (DAG) of marginal total effect (MTE).

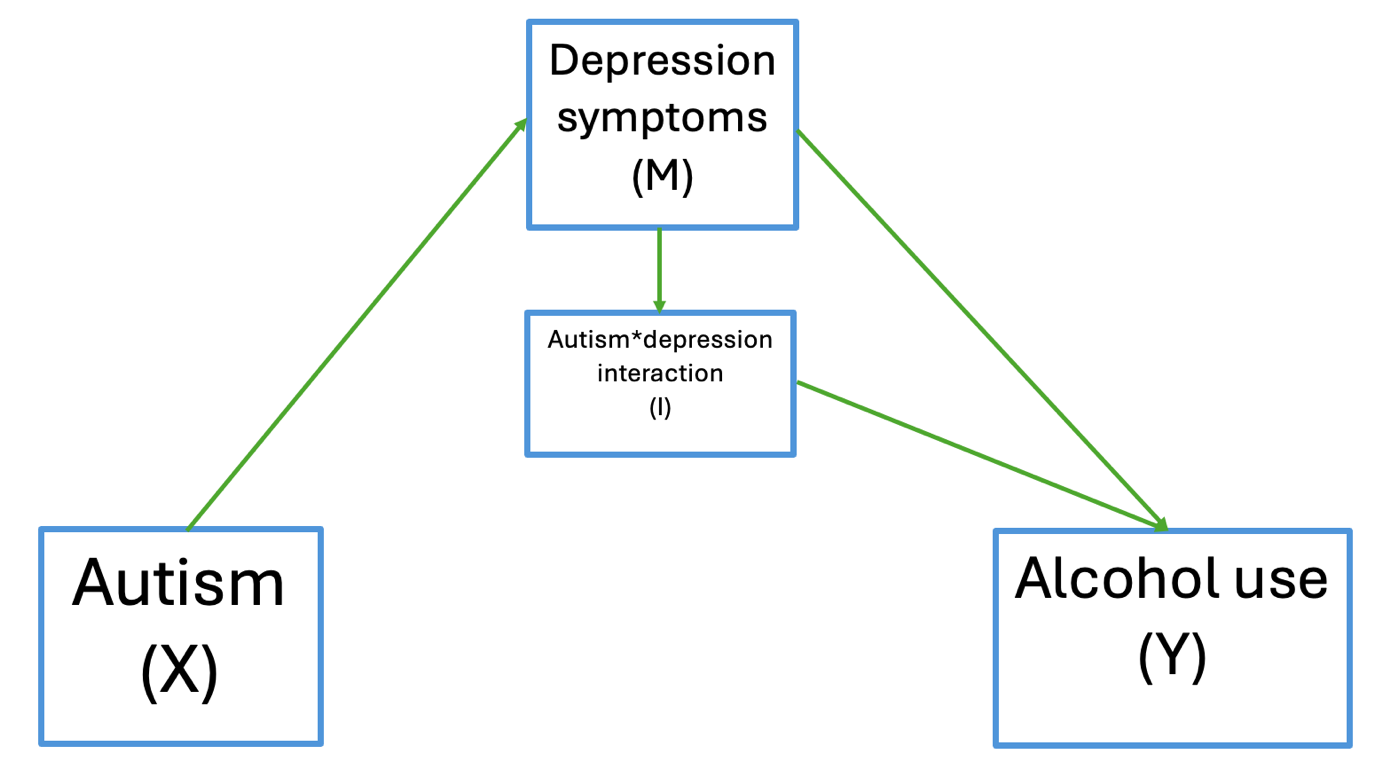

Sex differences

*Supplementary Figure S5*. Predicted mean alcohol use across time in males and females with 95% confidence intervals.

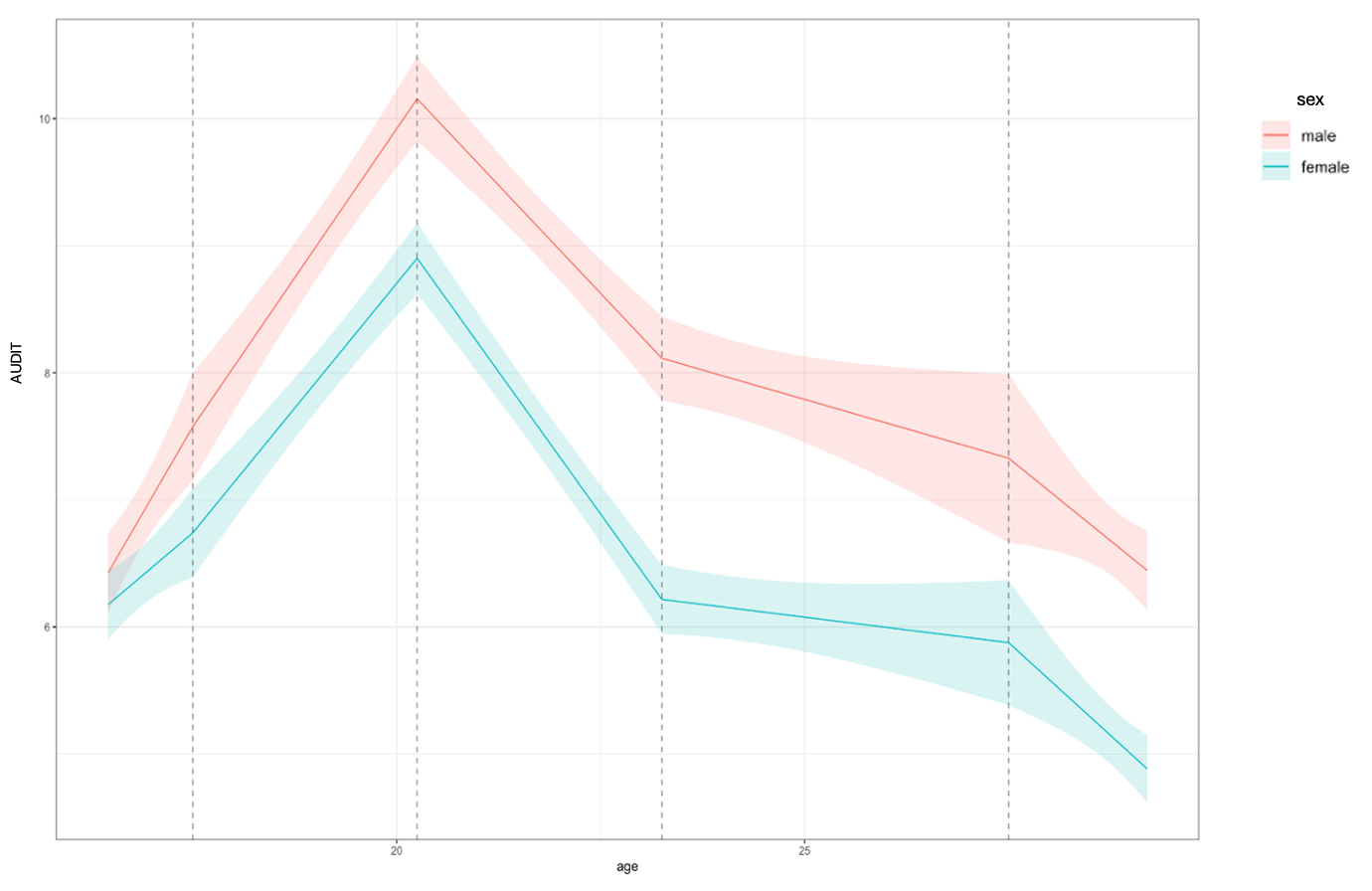

Sensitivity analyses

The following model checks were performed on the final baseline multilevel linear model looking at mean alcohol use over time with the autism mean factor score as the exposure (as this had the largest sample size of the three).

*Supplementary Figure S6*. Plot of individual-level predicted slopes (subsample of N=50 to assist visual clarity)

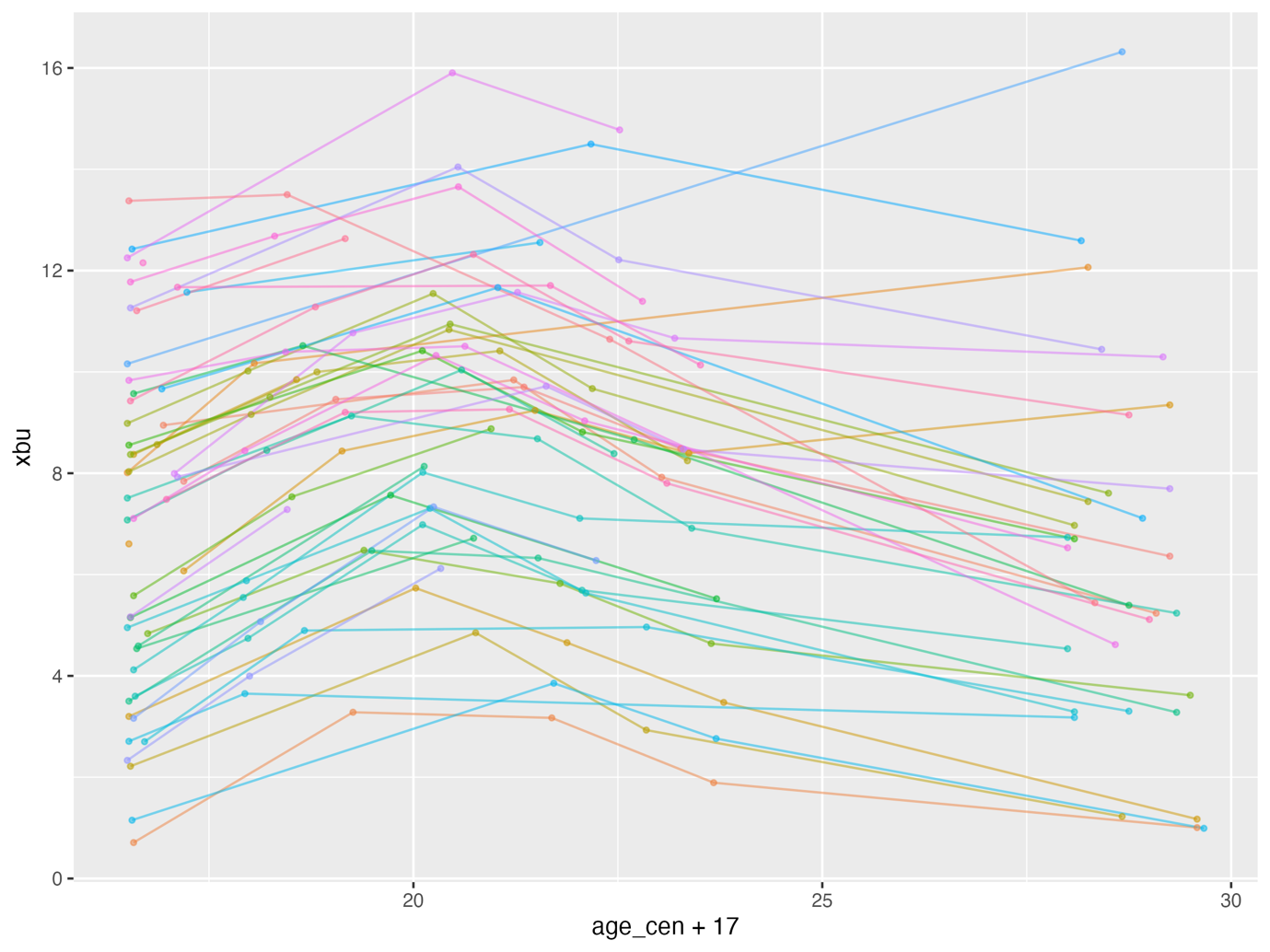

*Supplementary Figure S7*. Scatterplot of level 2 random intercept vs level 2 random slope residuals

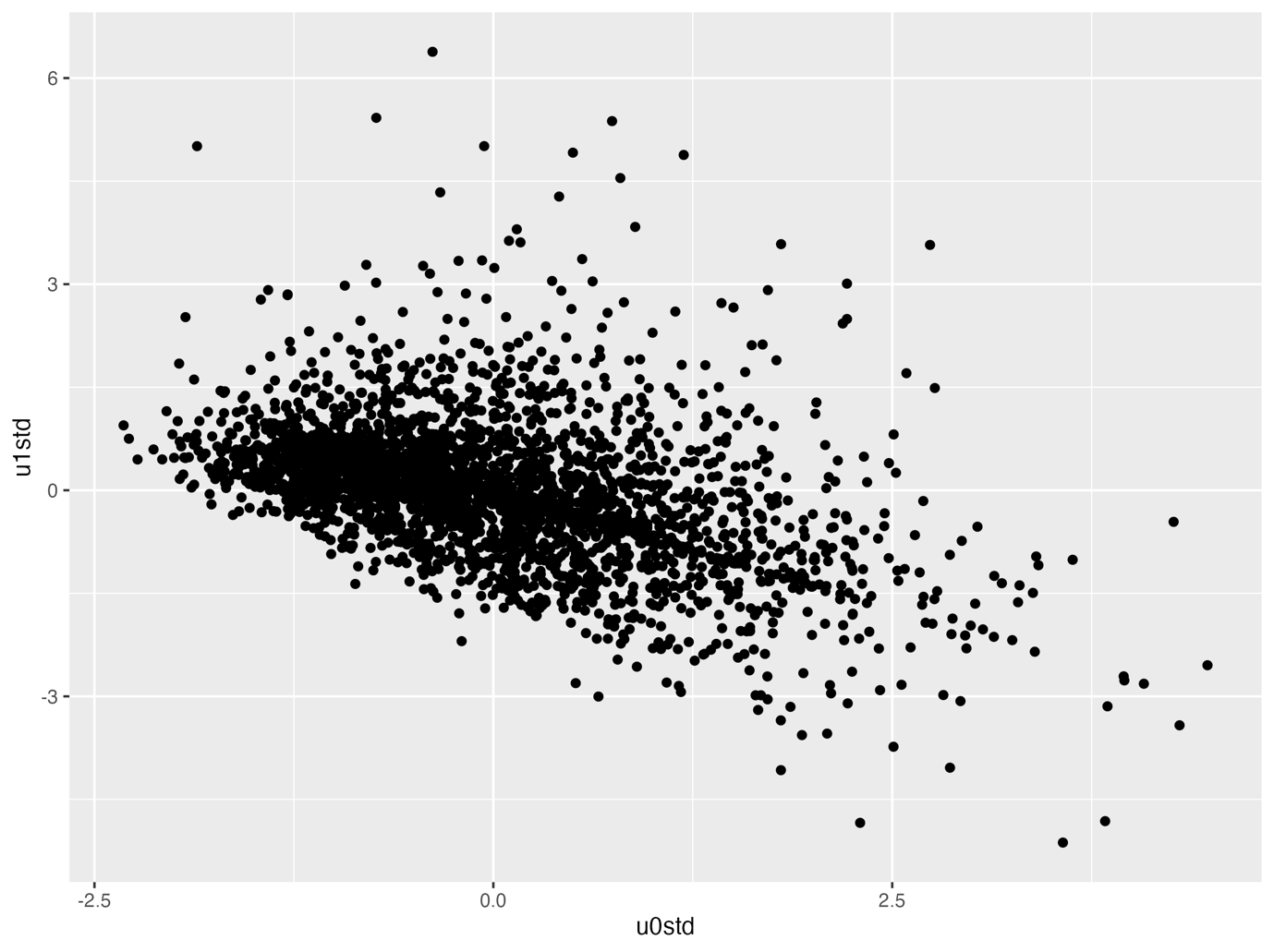

*Supplementary Figure S8*. QQ plot of level 1 residuals

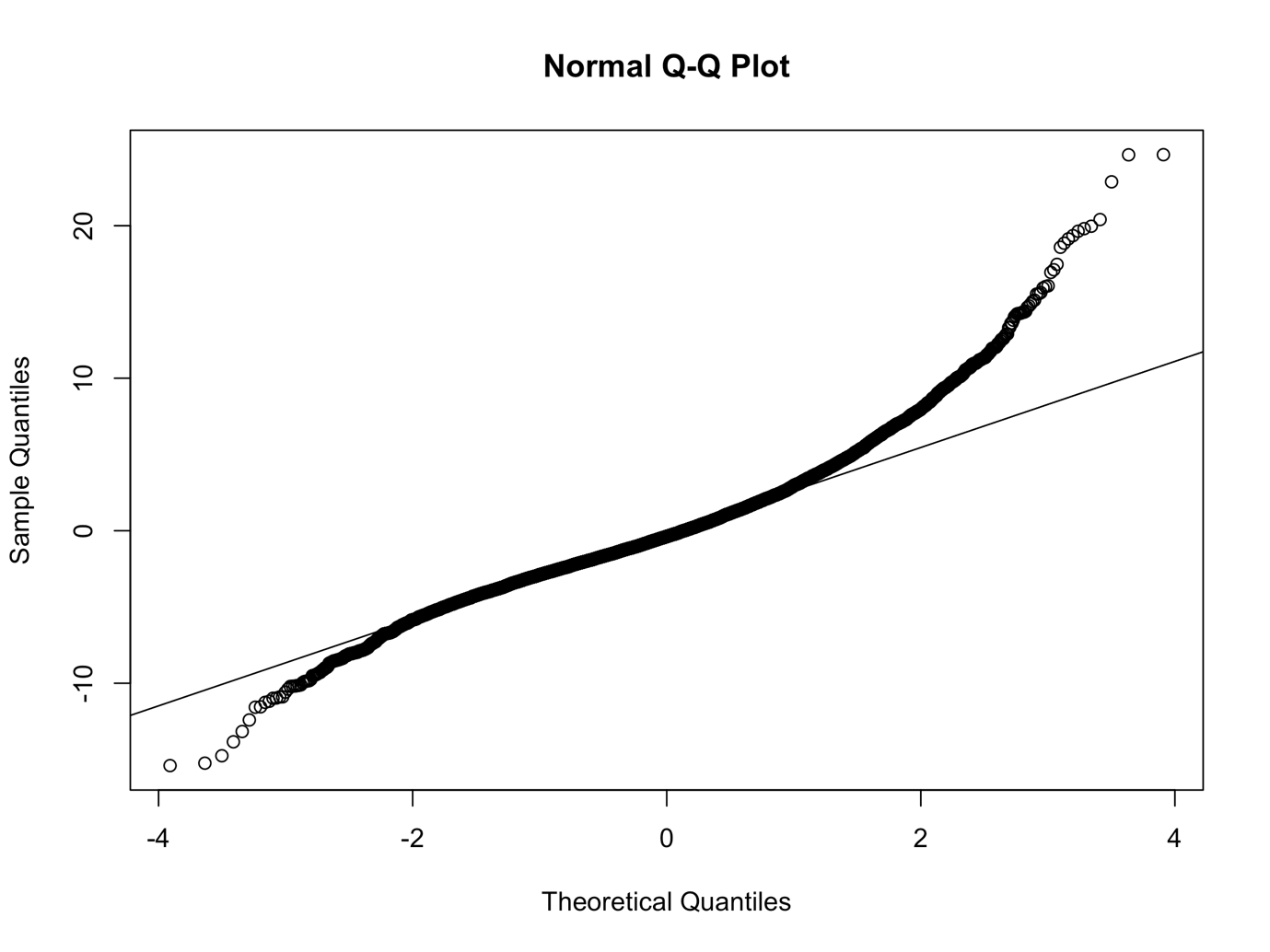

*Supplementary Figure S9.* Density plot of level 1 residuals

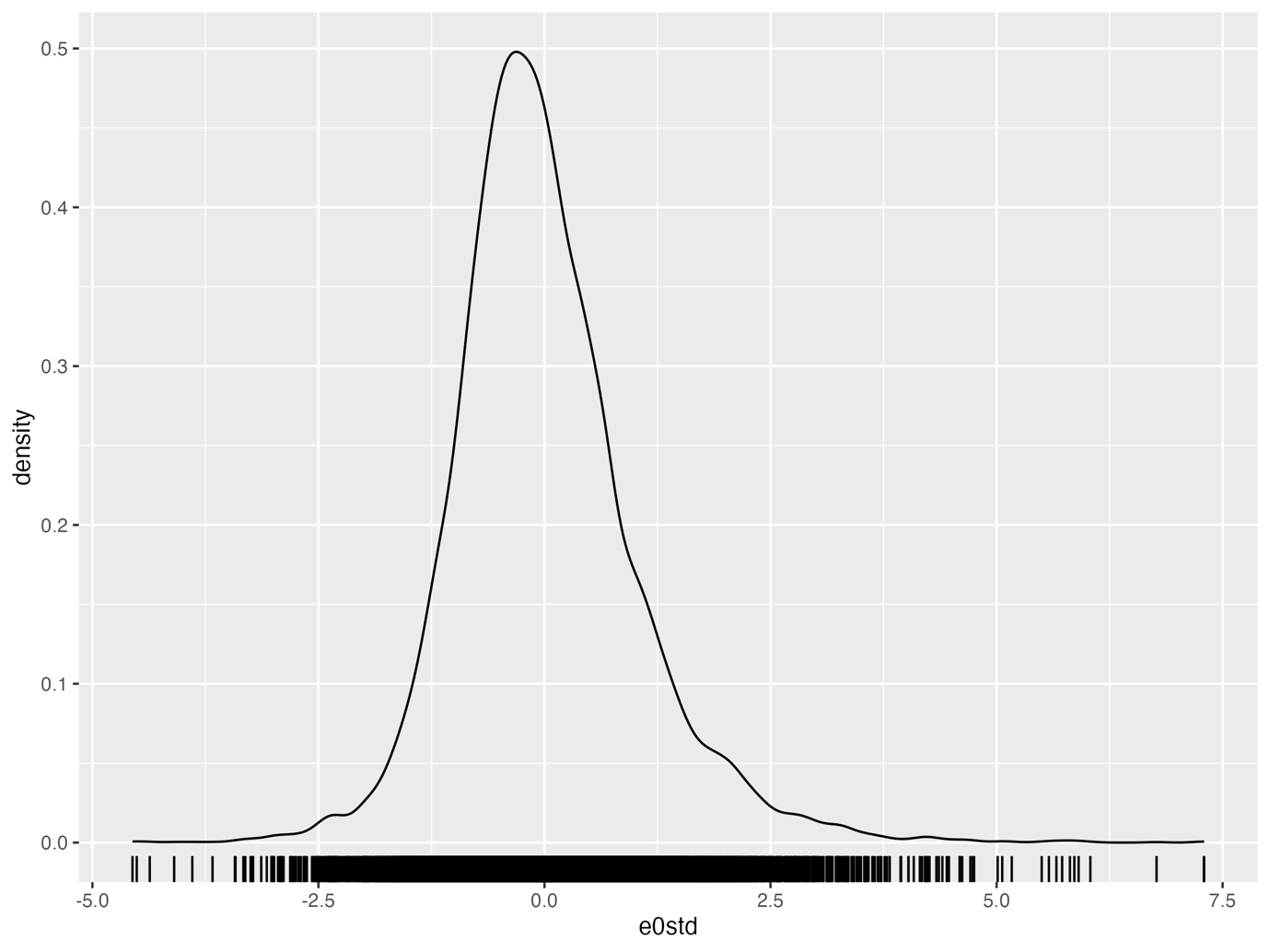

*Supplementary Figure S10*. Scatterplot of level 1 residuals vs. fitted values

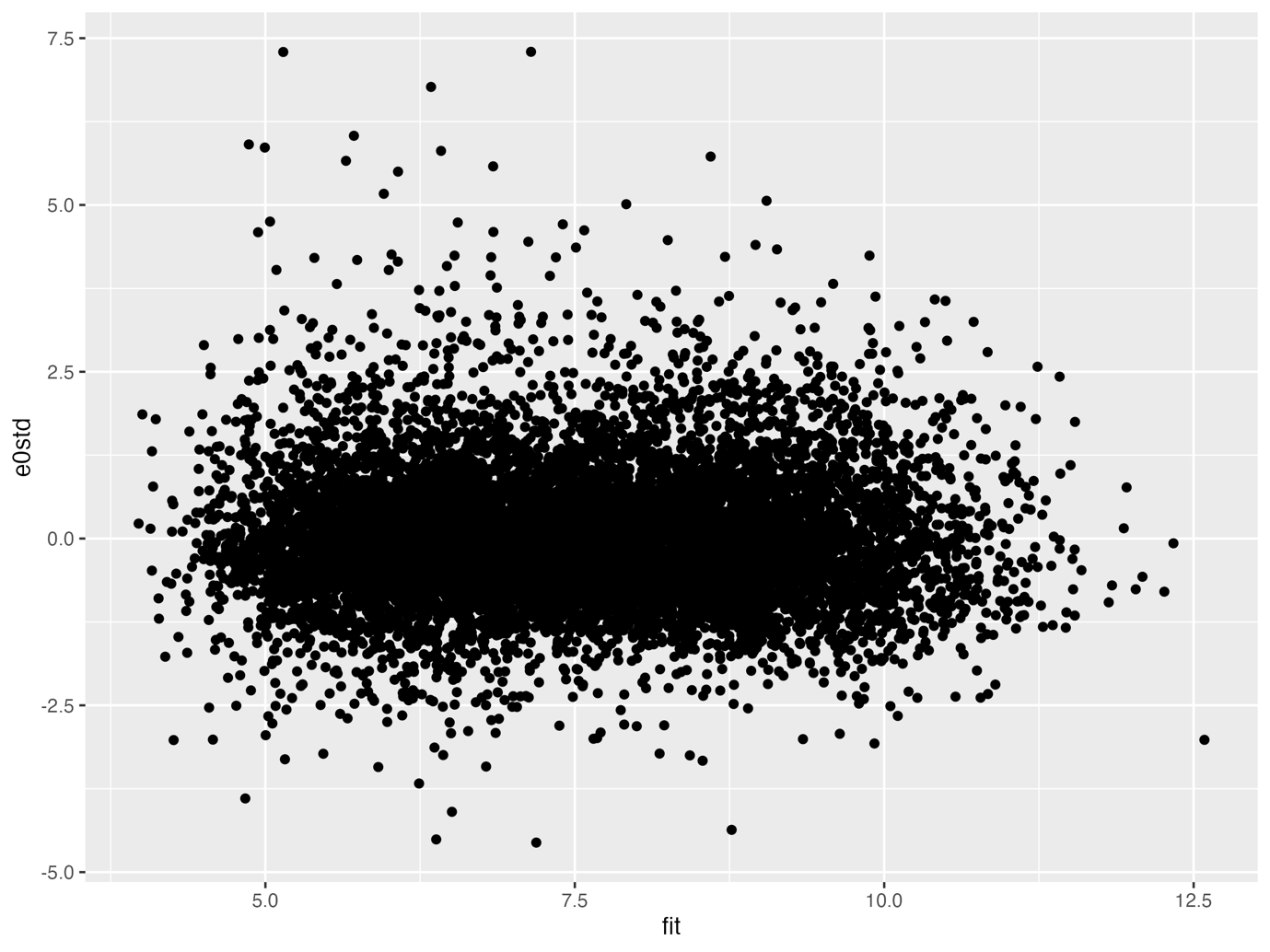

*Supplementary Figure S11*. QQ plot of level 2 random intercept residuals

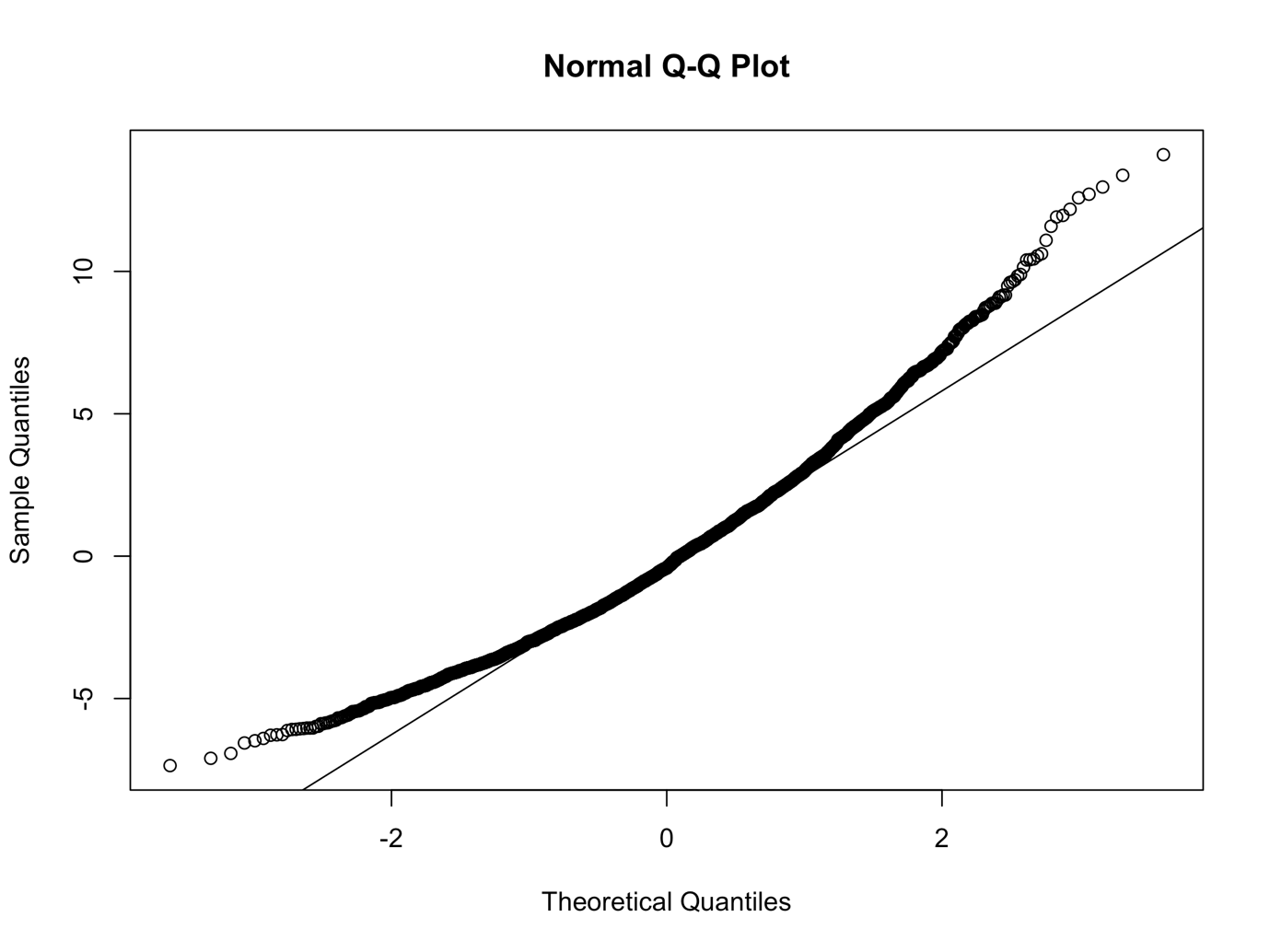

*Supplementary Figure S12*. Density plot of level 2 random intercept residuals

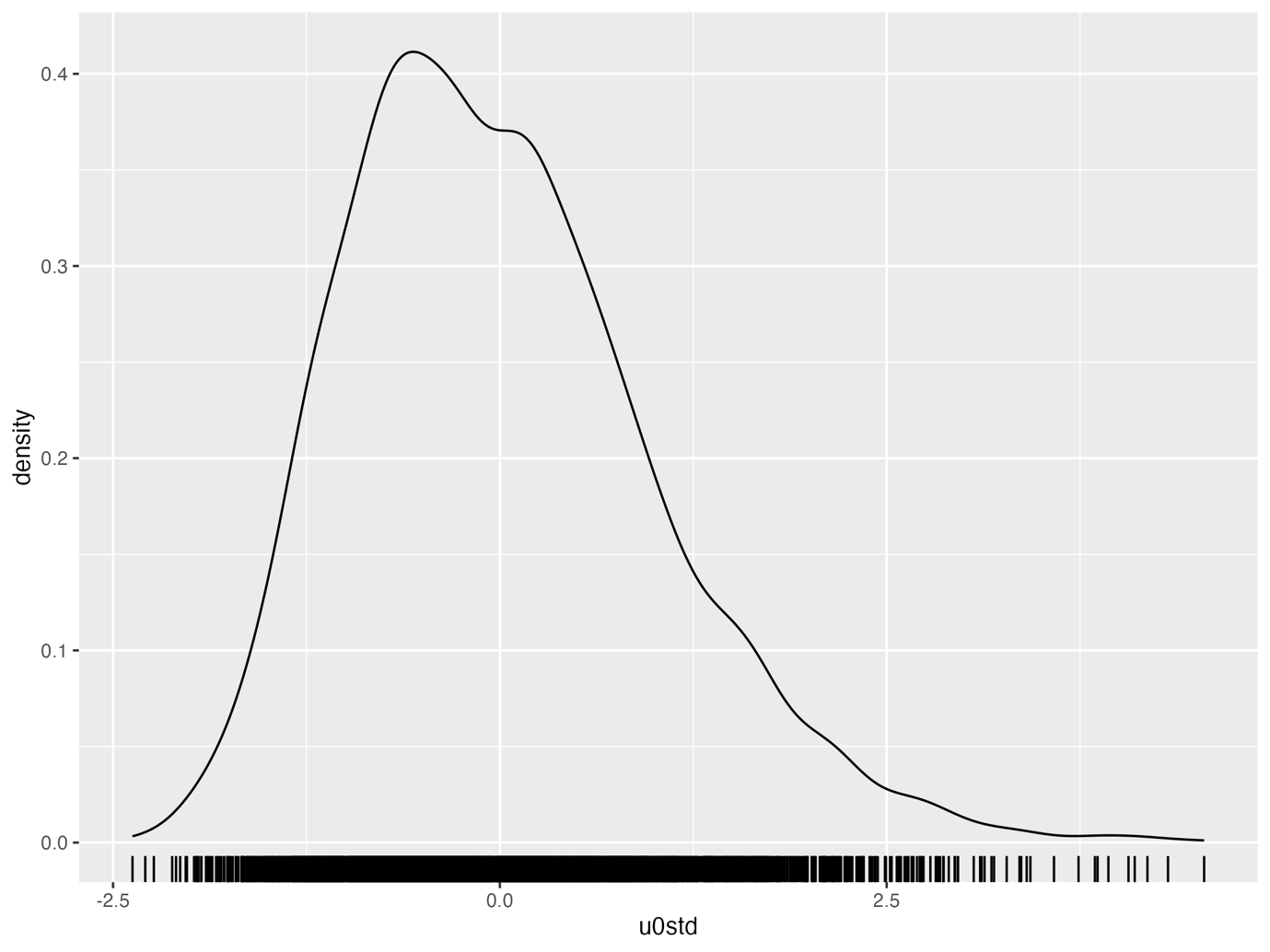

*Supplementary Figure S13*. QQ plot of Level 2 random slope residuals

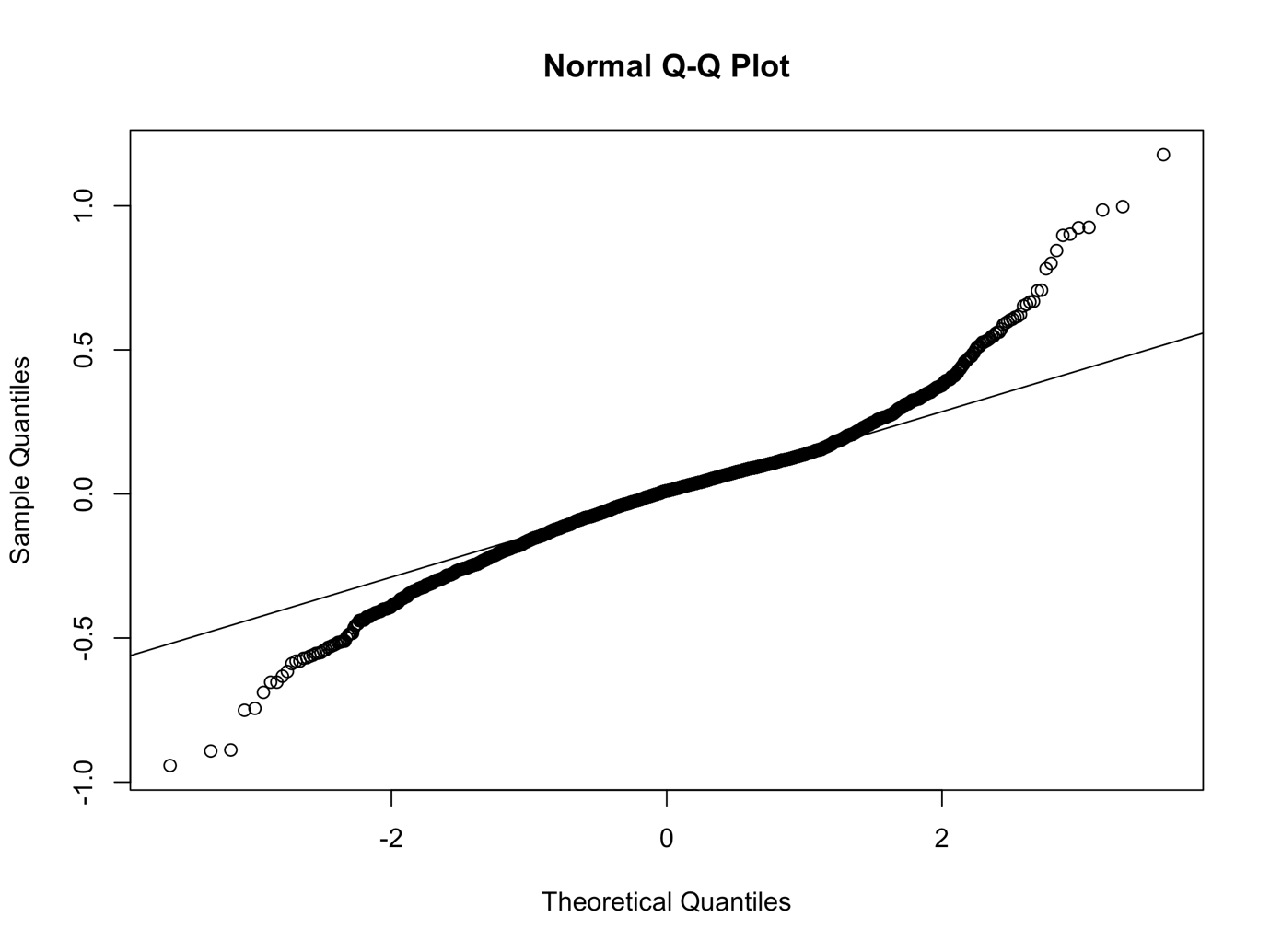

*Supplementary Figure S14*. Density plot of level 2 standardised residuals

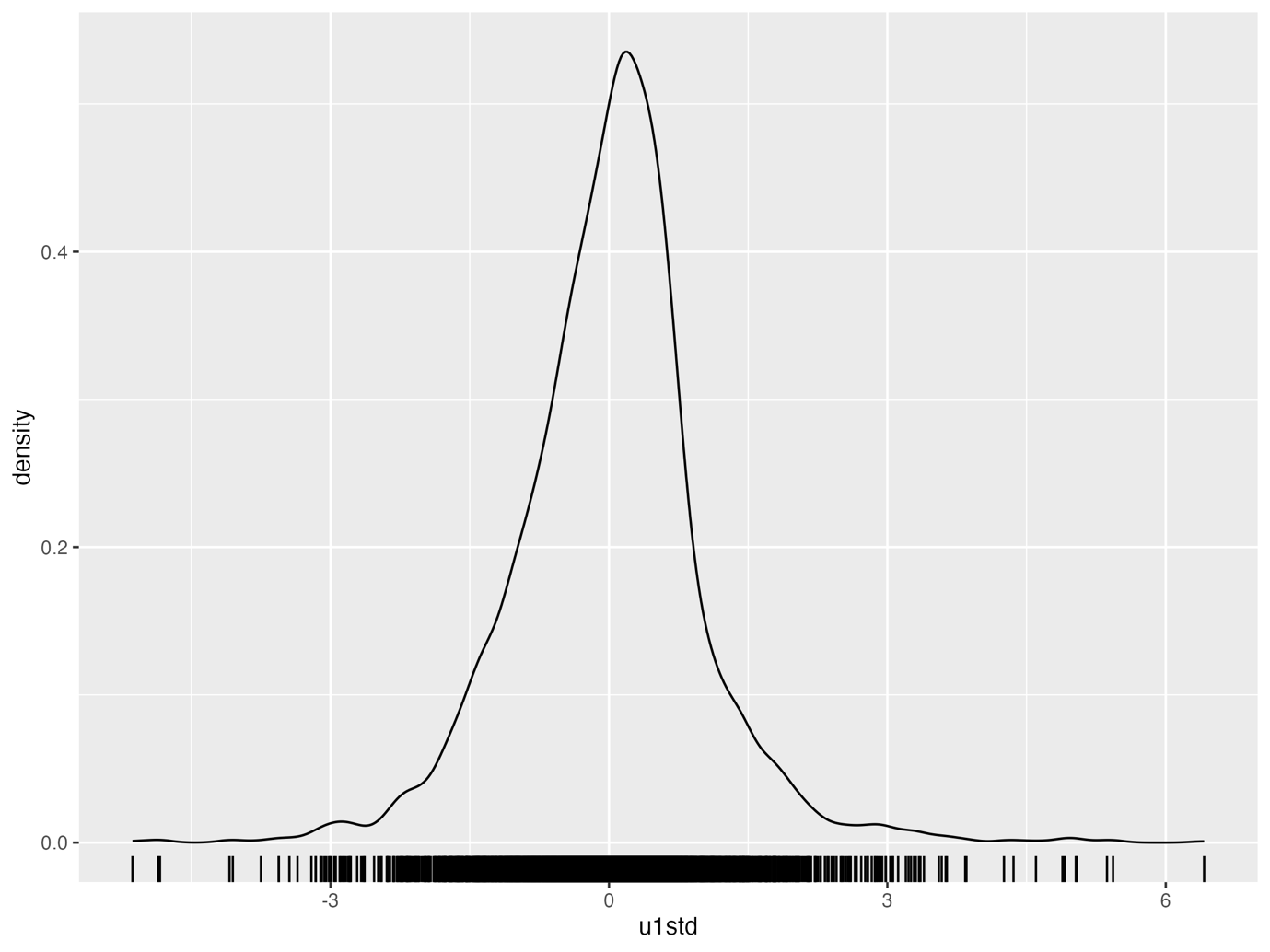

*Supplementary Figure S15*. Scatter plot of level 2 random slopes residuals vs. fitted values

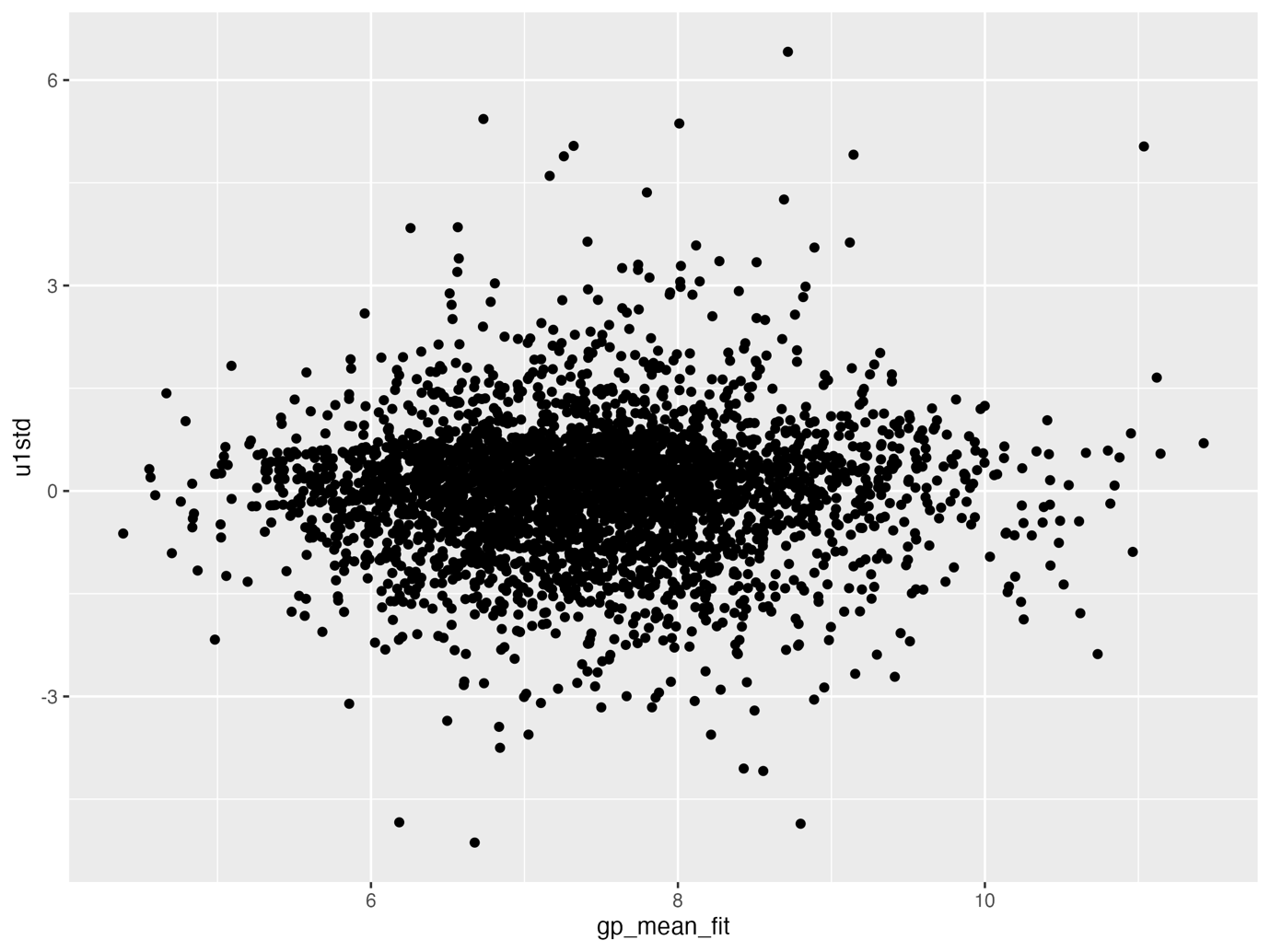

*Supplementary Figure S16.* Scatter plot of level 2 random intercept residuals vs. fitted values

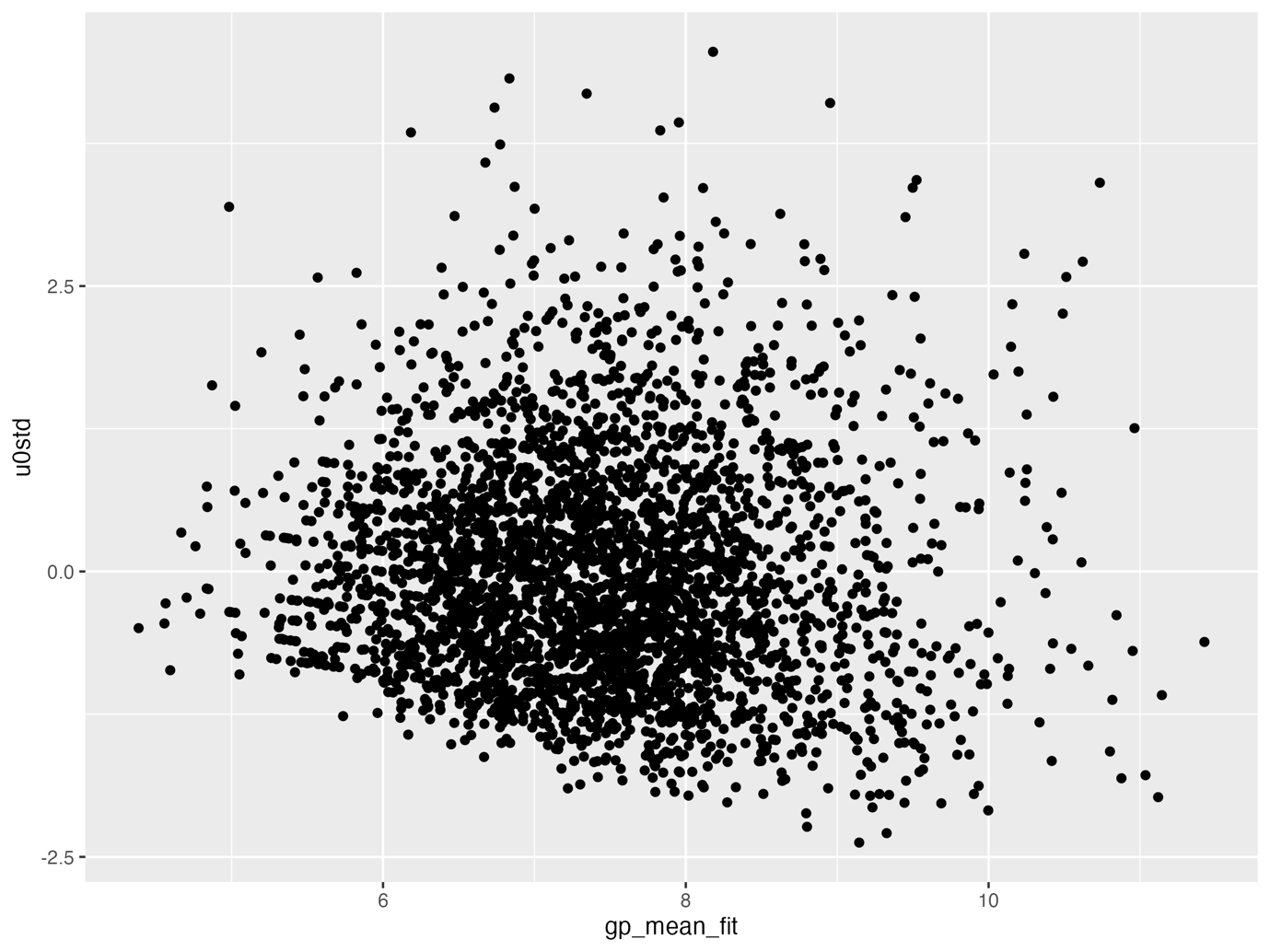

*Supplementary Figure S17*. Plot of variance functions (levels 1 and 2)

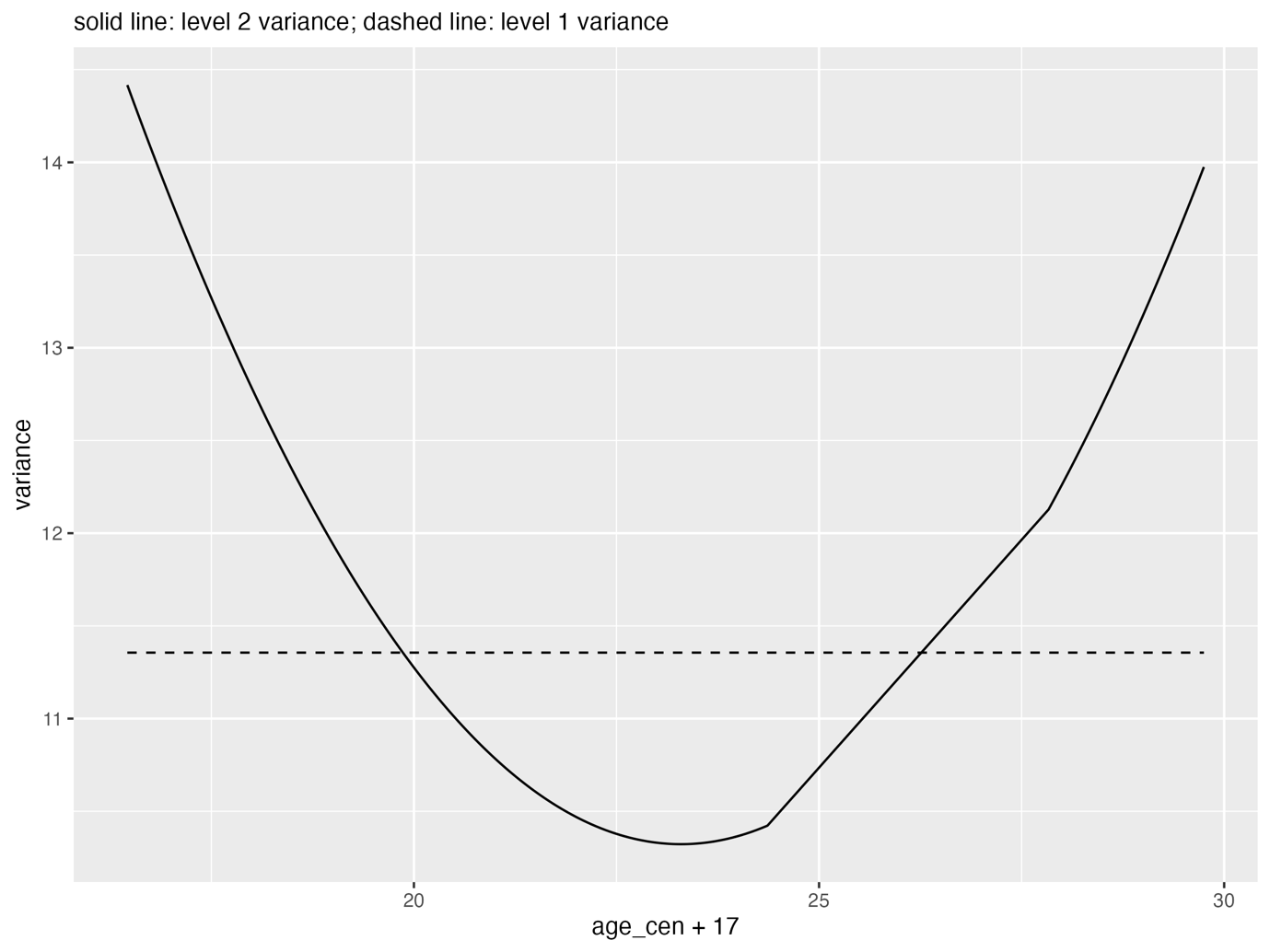

*Supplementary Figure S18*. Violin plot of residuals binned into intervals of age

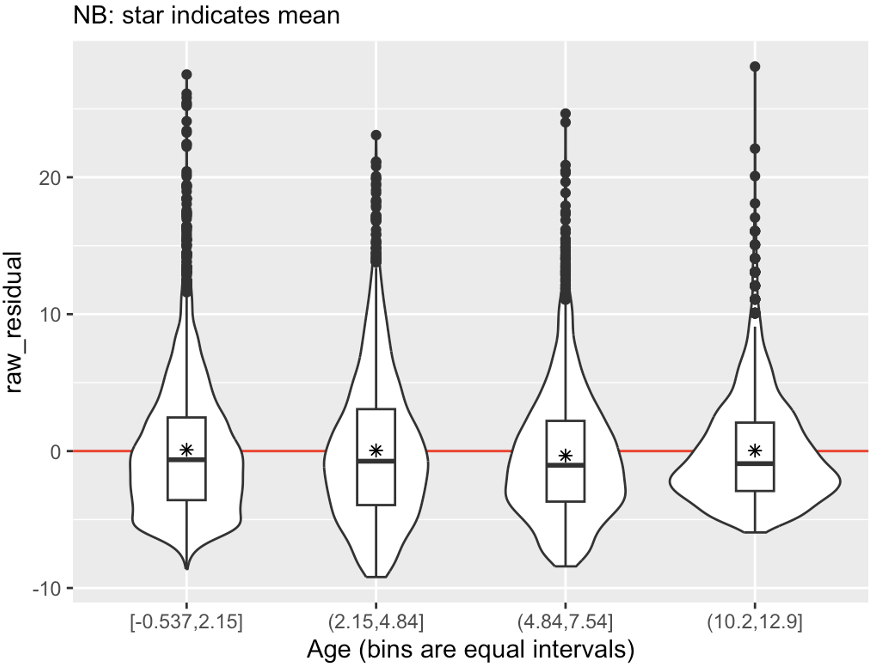

The following model checks were performed on the final baseline multilevel linear model with AUDIT log transformed for comparison with main analyses.

*Supplementary Figure S19*. QQ plot of level 1 residuals

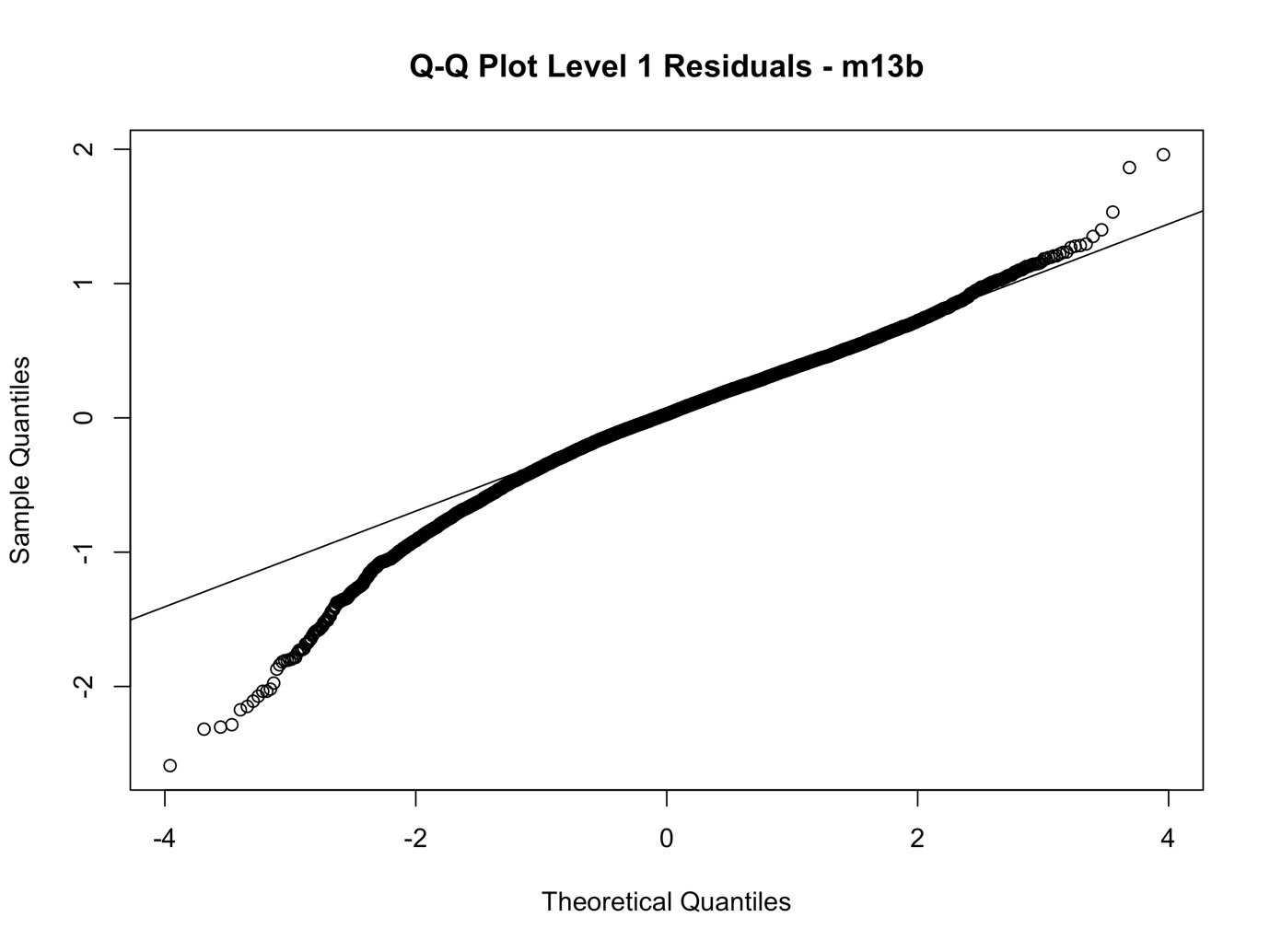

*Supplementary Figure S20.* Density plot of level 1 residuals

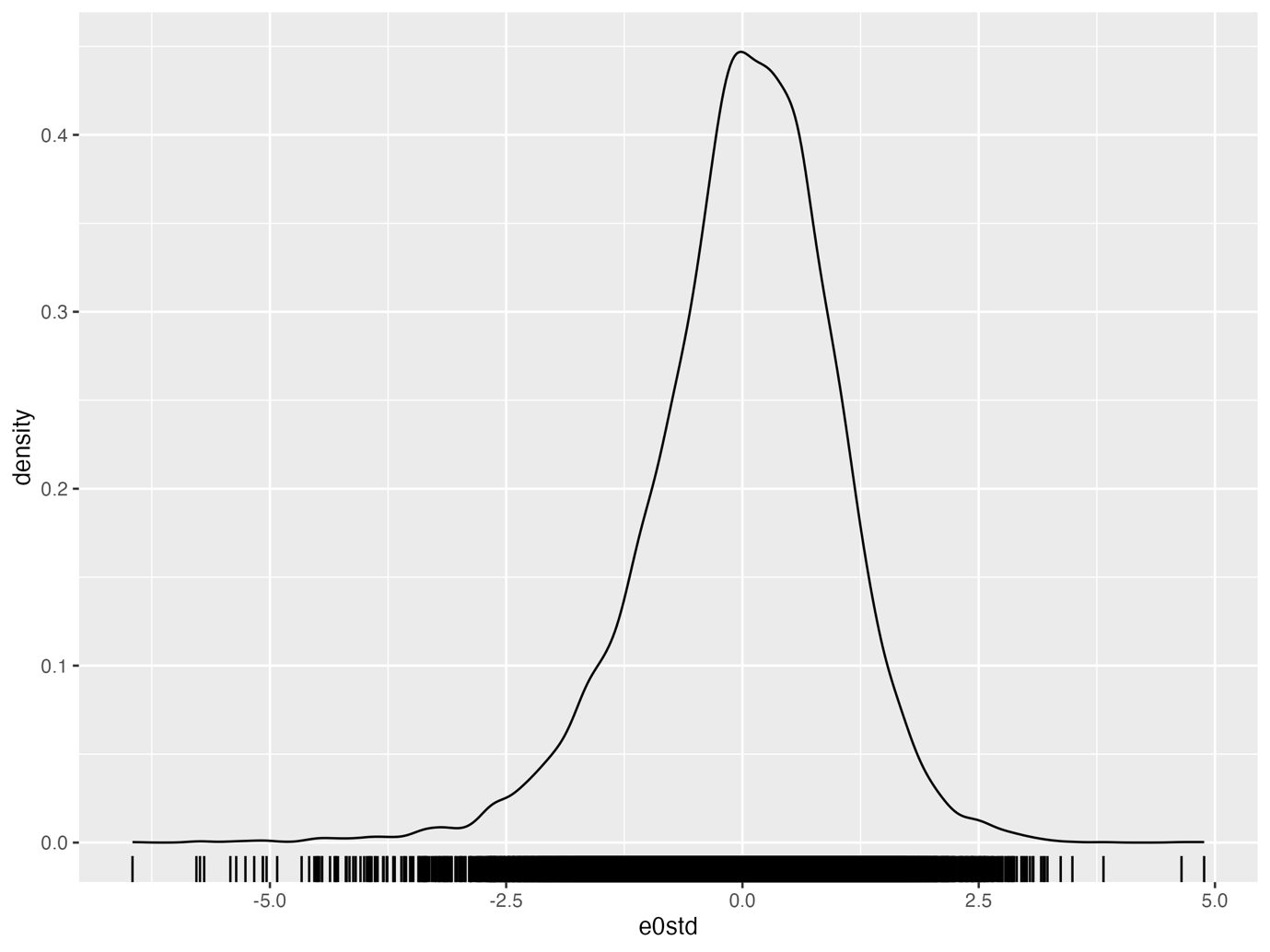

*Supplementary Figure S21*. QQ plot of level 2 random intercept residuals

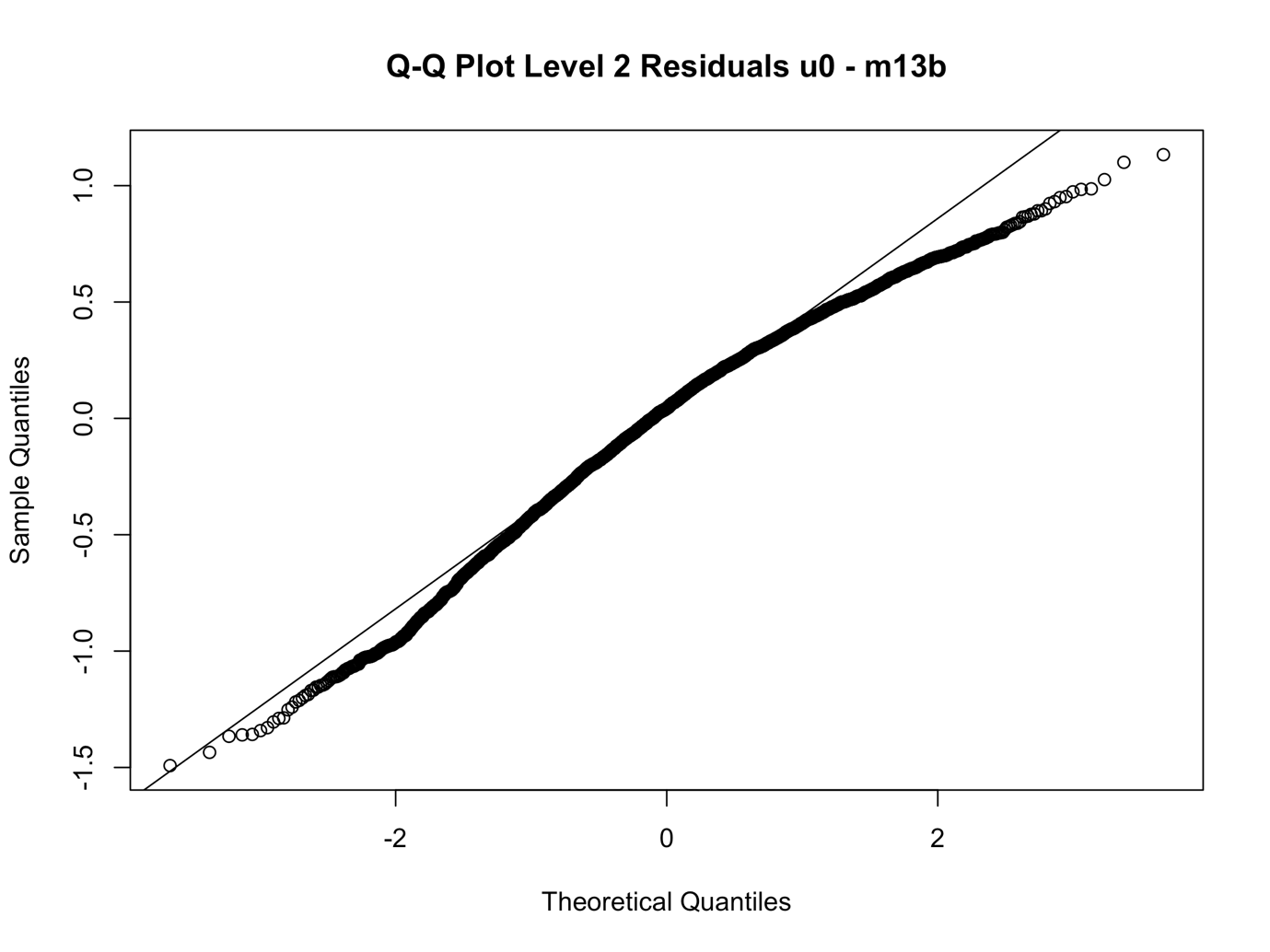

*Supplementary Figure S22*. Density plot of level 2 random intercept residuals

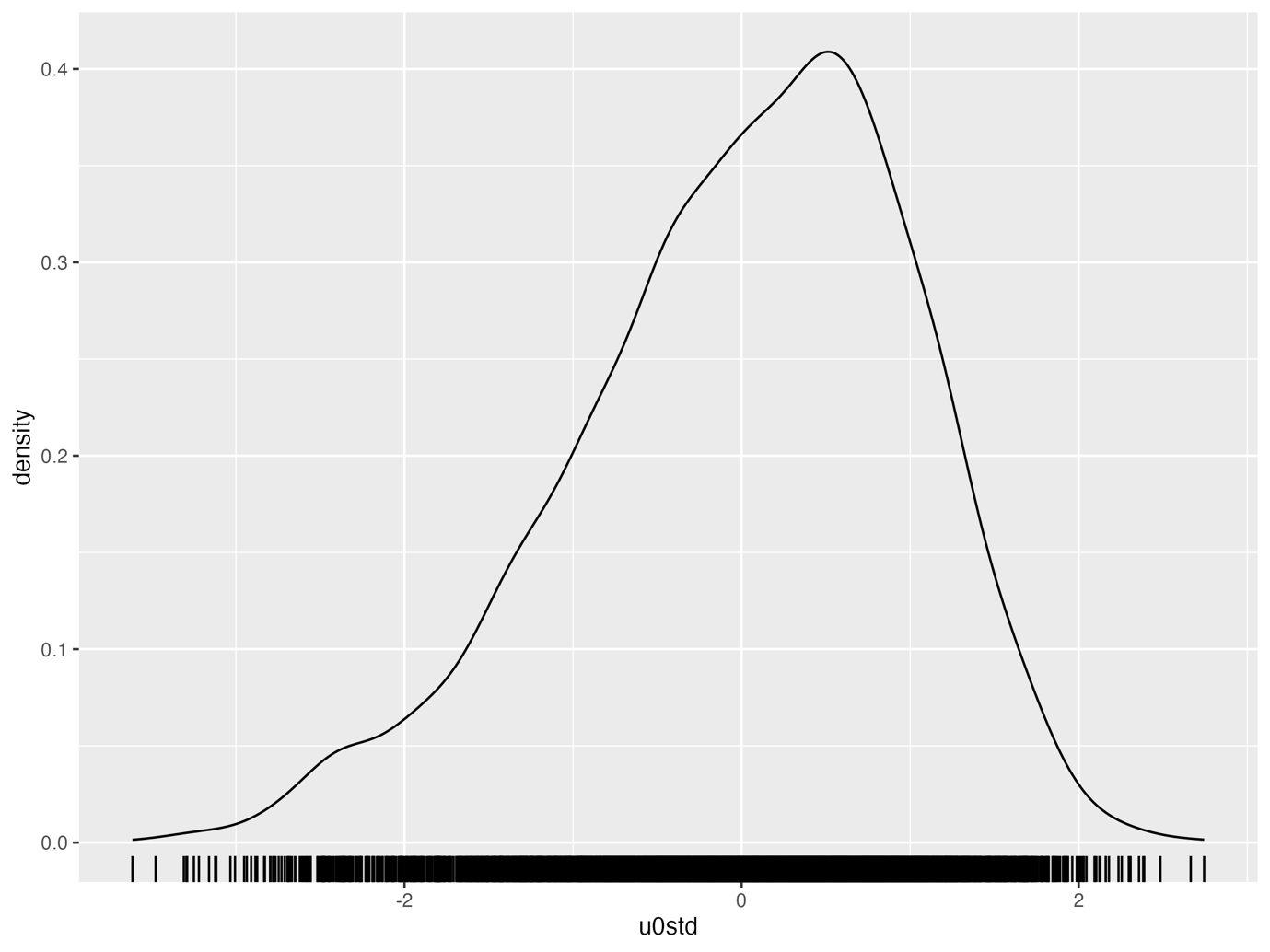

*Supplementary Figure S23*. QQ plot of Level 2 random slope residuals

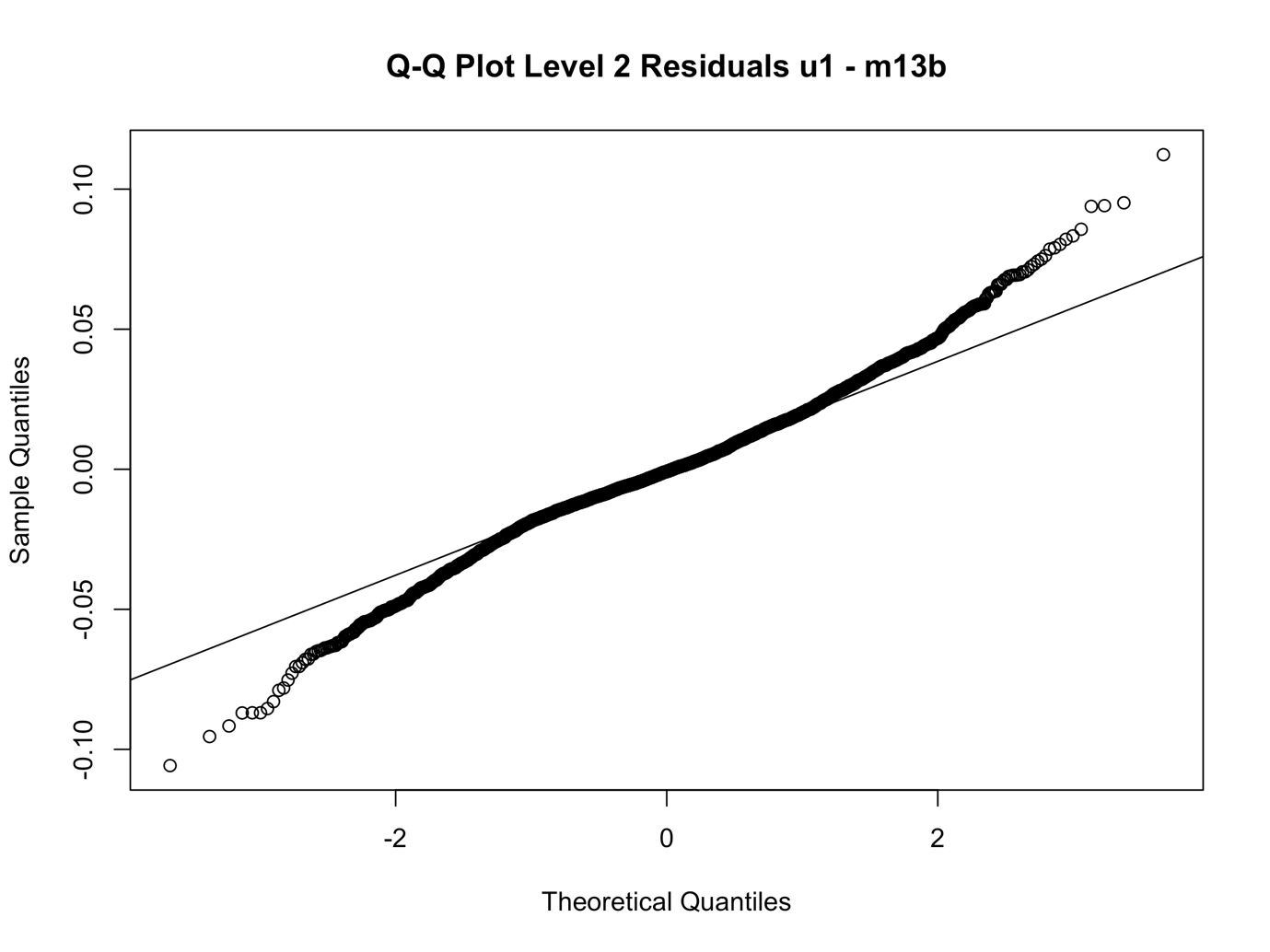

*Supplementary Figure S24.* Density plot of level 2 standardised residuals

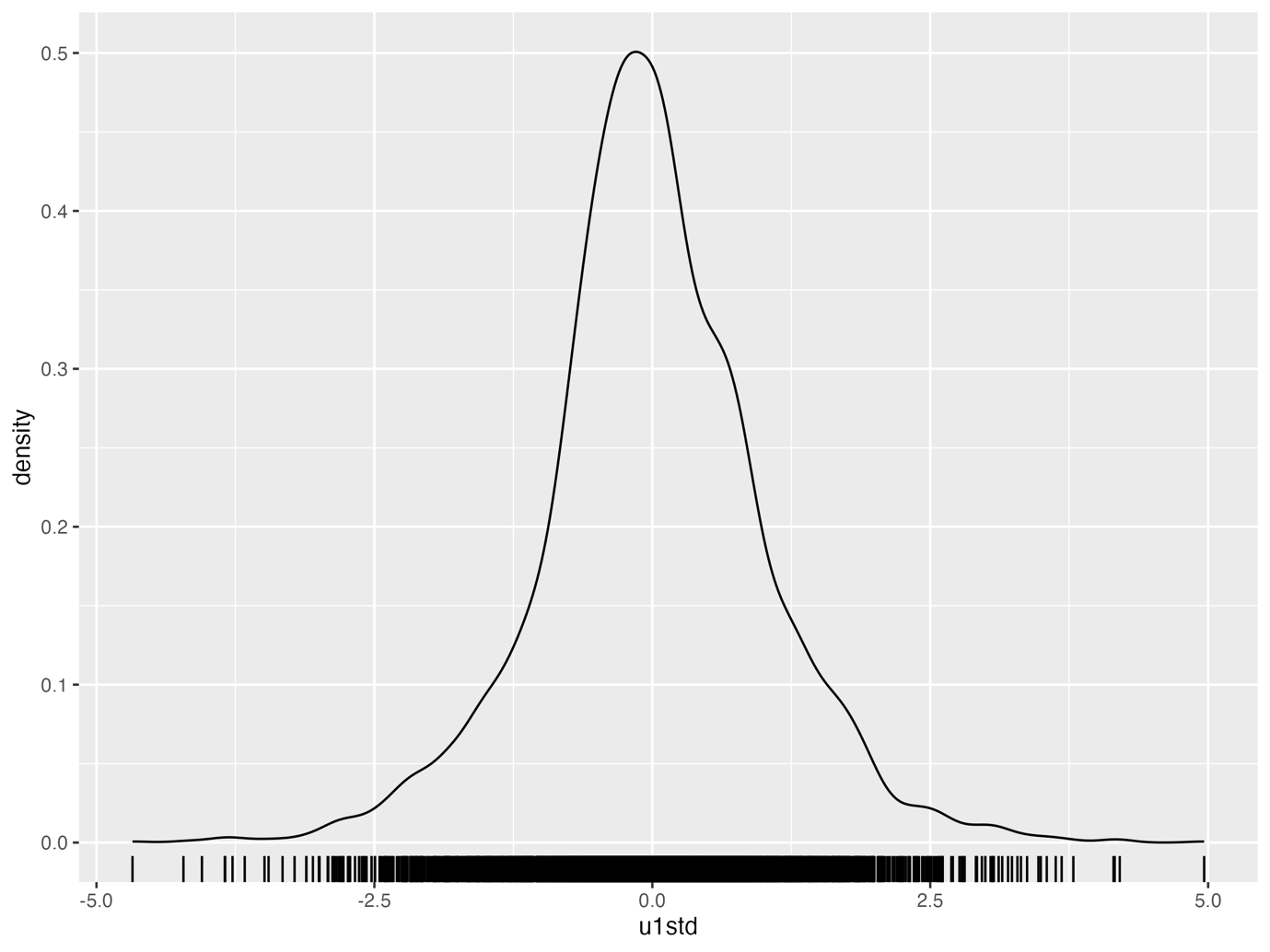

The following model checks were performed on the final linear quantile mixed models (LQMM) looking at AUDIT (Alcohol Use Disorders Identification Test) at the 10^th^, 50th and 90^th^ percentiles.

*Supplementary Figure S25*. Scatter plot of level 2 residuals from LQMM where an autism mean factor score was the exposure and 10^th^ percentile AUDIT scores were modelled.

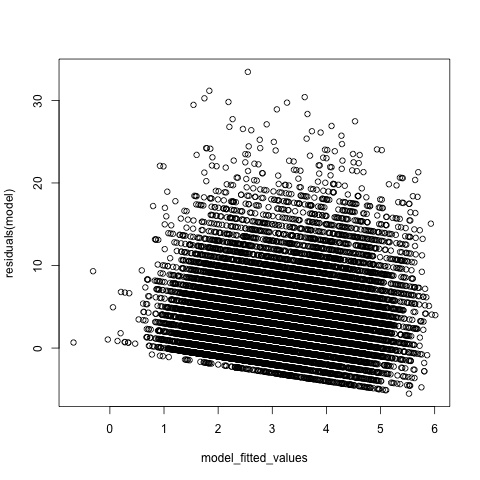

*Supplementary Figure S26*. Scatter plot of level 2 residuals from LQMM where an autism mean factor score was the exposure and 50^th^ percentile AUDIT scores were modelled.

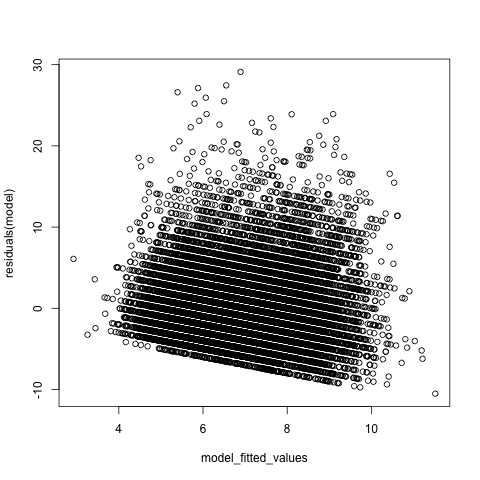

*Supplementary Figure S27.*  Scatter plot of level 2 residuals from LQMM where an autism mean factor score was the exposure and 90^th^ percentile AUDIT scores were modelled.

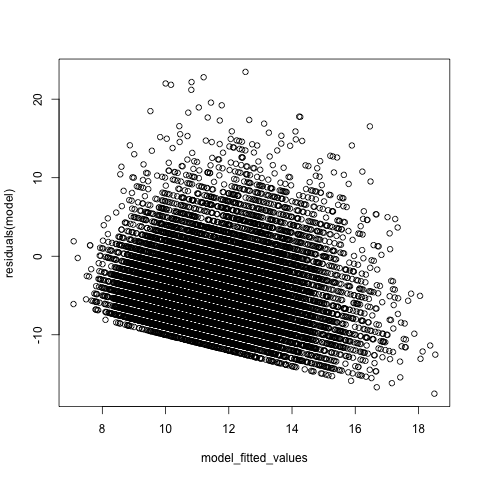

*Supplementary Figure S28*. Scatter plot of level 2 residuals from LQMM where scores derived from parent-reported responses to the Social Communication Disorders Checklist at 7 years were used as the exposure and 10^th^ percentile AUDIT scores were modelled.

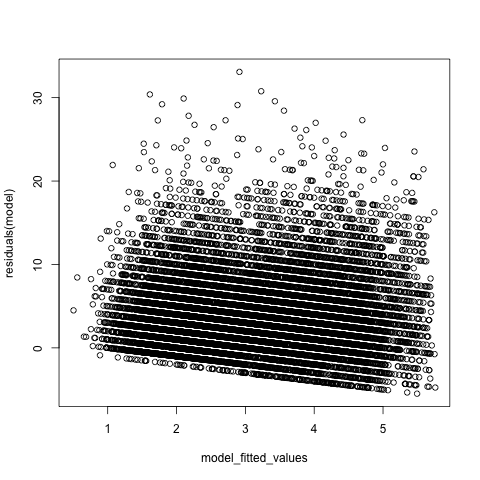

*Supplementary Figure S29*. Scatter plot of level 2 residuals from LQMM where scores derived from parent-reported responses to the Social Communication Disorders Checklist at 7 years were used as the exposure and 50^th^ percentile AUDIT scores were modelled.

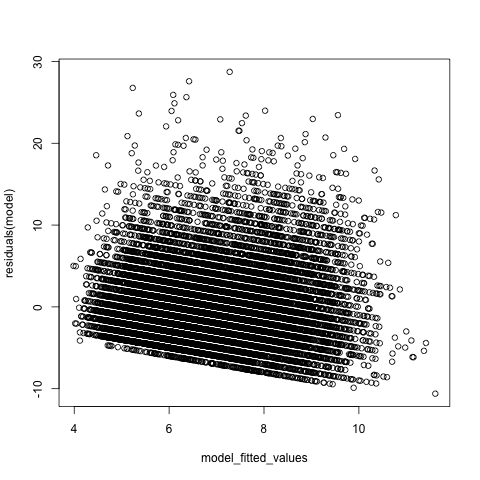

*Supplementary Figure S30.* Scatter plot of level 2 residuals from LQMM where scores derived from parent-reported responses to the Social Communication Disorders Checklist at 7 years were used as the exposure and 90^th^ percentile AUDIT scores were modelled.

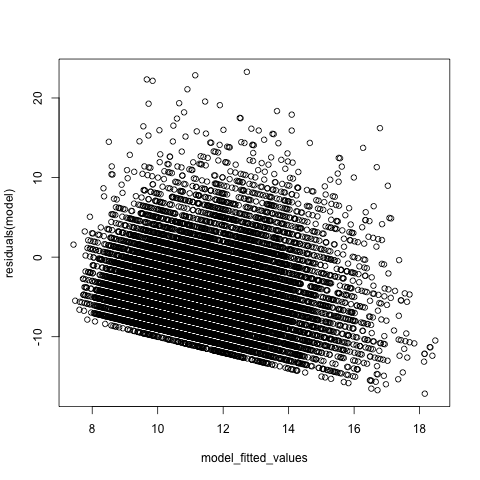

*Supplementary Figure S31*. Scatter plot of level 2 residuals from LQMM where a polygenic score for autism was the exposure and 10^th^ percentile AUDIT scores were modelled.

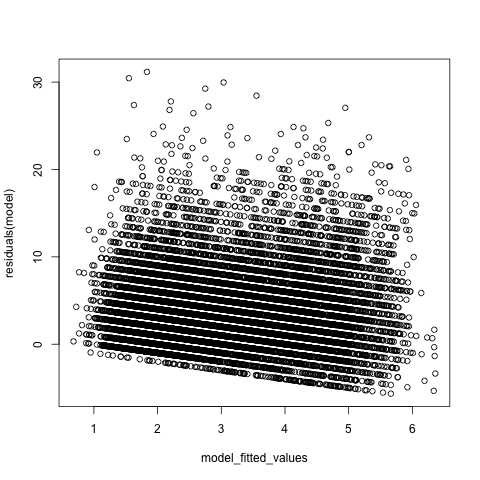

*Supplementary Figure S32.* Scatter plot of level 2 residuals from LQMM where a polygenic score for autism was the exposure and 50^th^ percentile AUDIT scores were modelled.

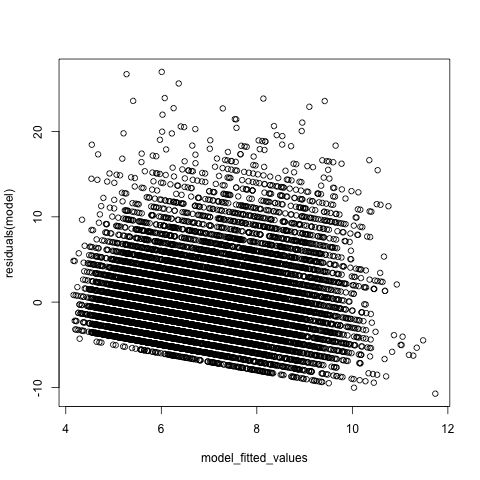

*Supplementary Figure S33.* Scatter plot of level 2 residuals from LQMM a polygenic score for autism was the exposure and 90^th^ percentile AUDIT scores were modelled.
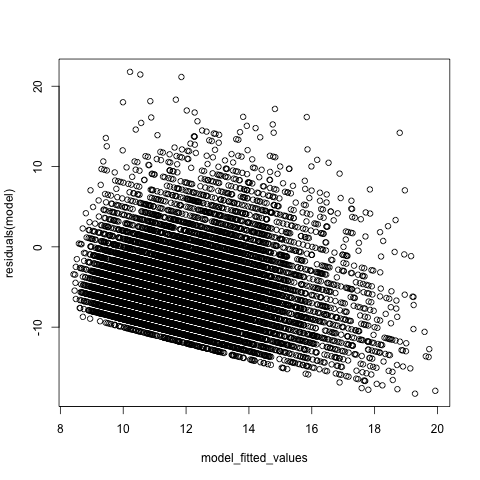

*Supplementary Figure S34*. Histogram of autism mean factor score

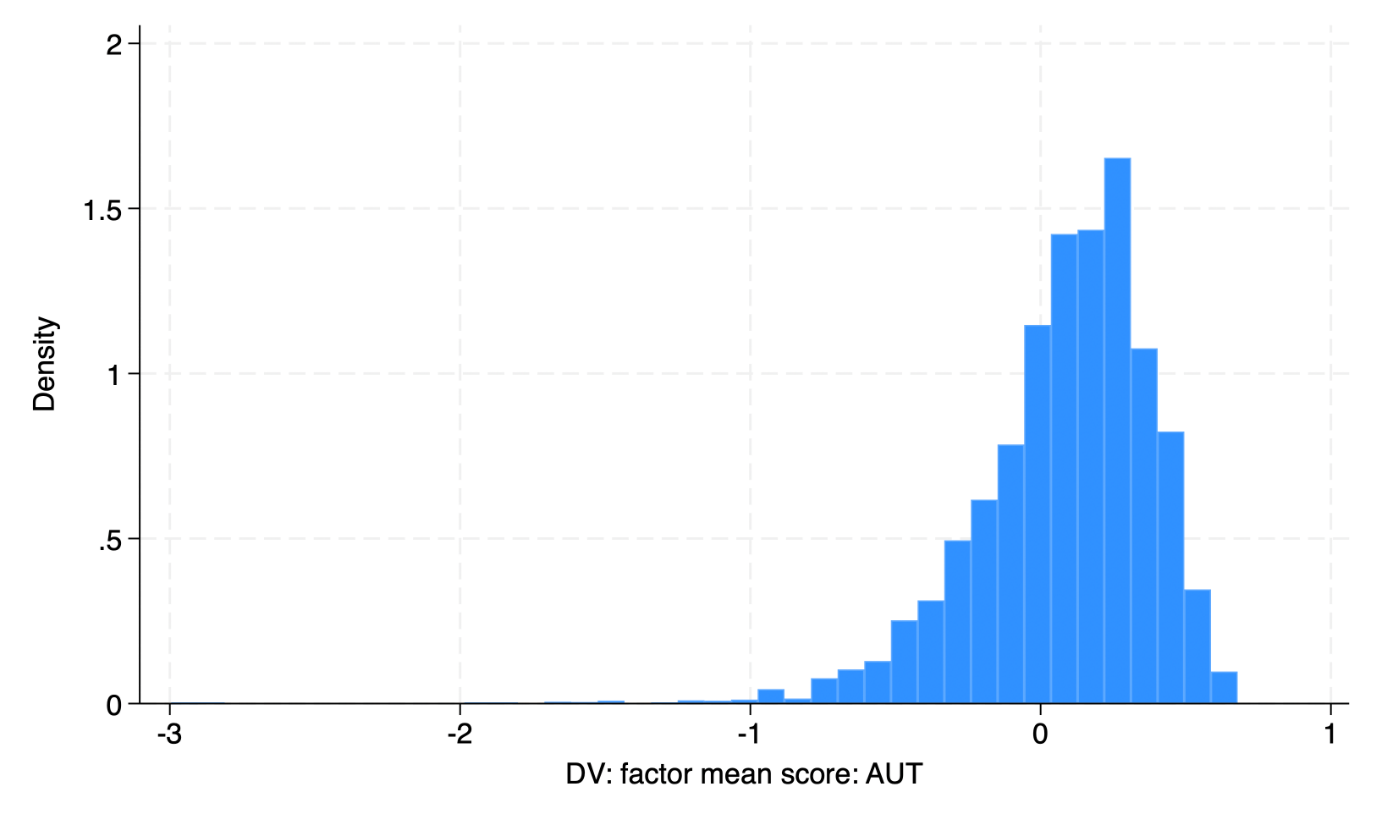

*Supplementary Figure S35*. Histogram of Social Communication Disorders Checklist (SCDC) score at 7 years

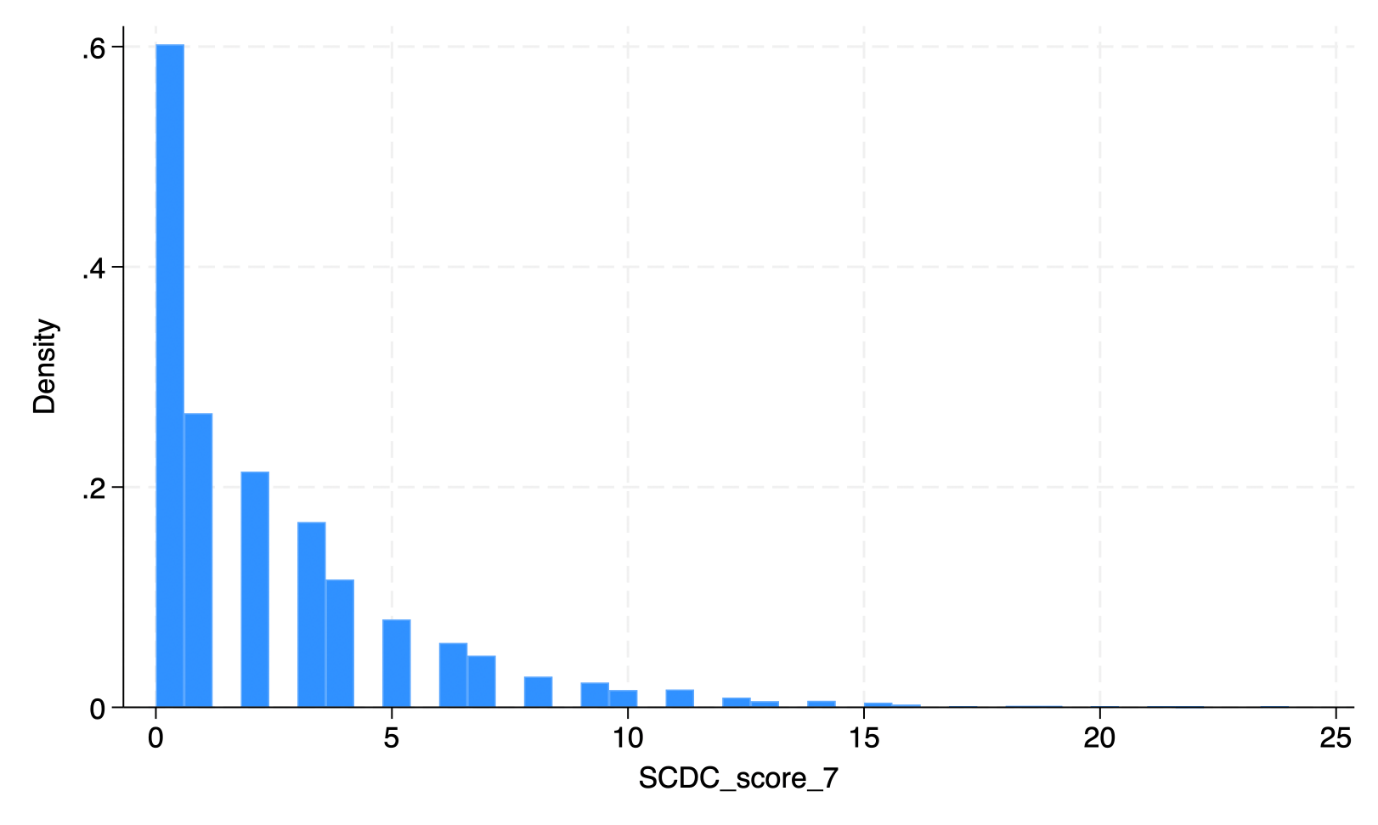

*Supplementary Figure S36*. Histogram of polygenic score for autism

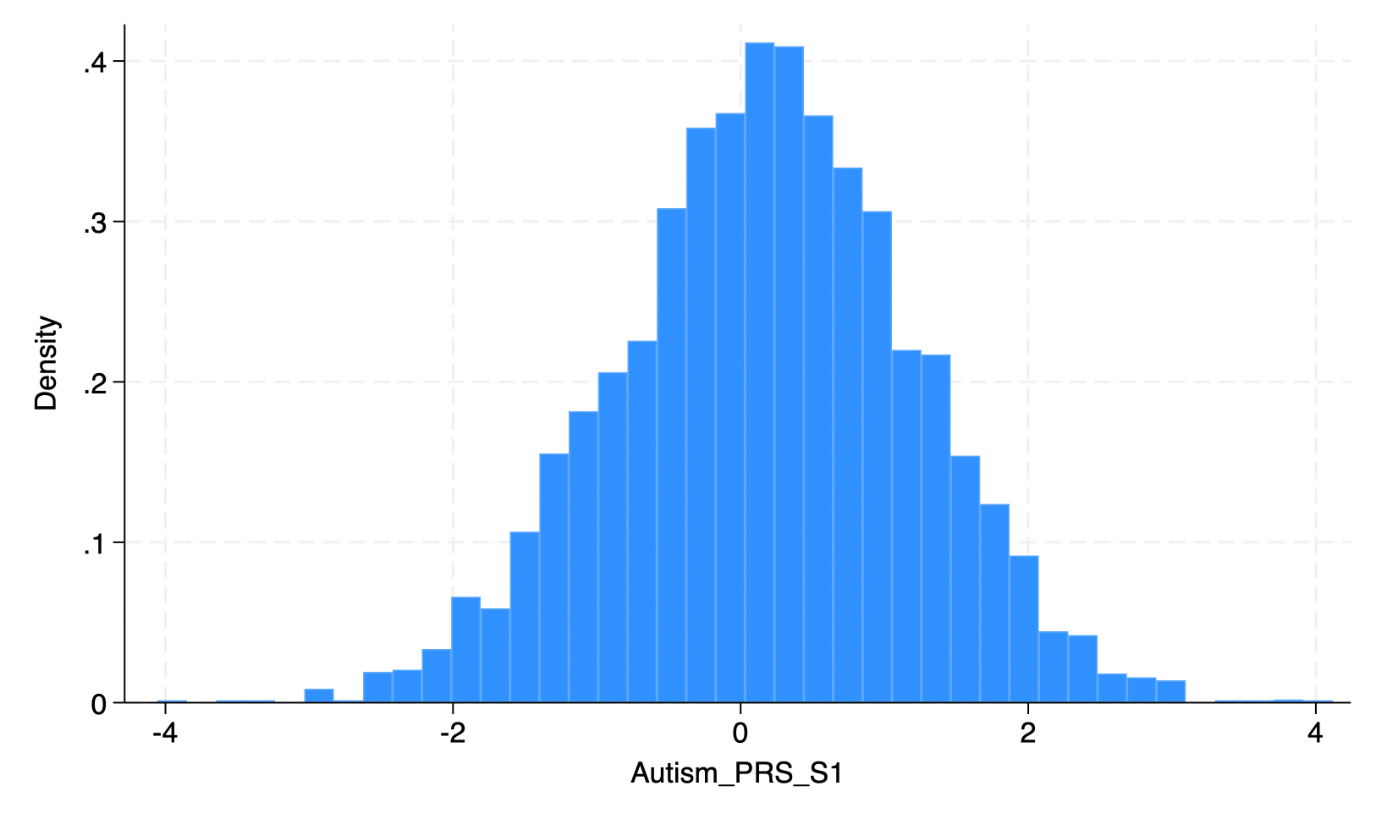

*Supplementary Figure S37.* Histogram of Strengths and Difficulties Questionnaire (SDQ) Hyperactivity Subscale score (measure of attention deficit hyperactivity disorder traits)

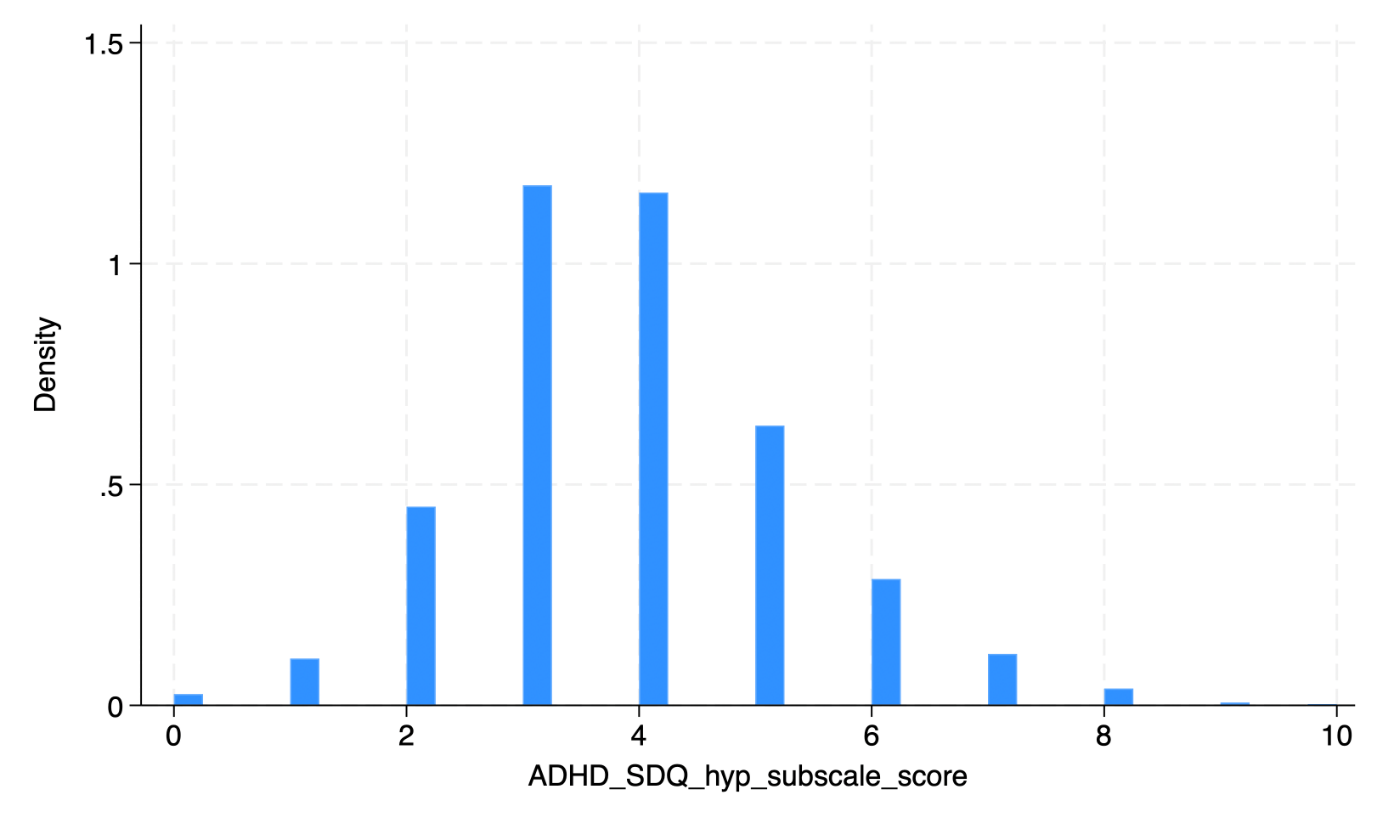

*Supplementary Figure S38.* Histogram of Short Mood and Feelings Questionnaire (SMFQ) score (measure of depression symptoms)

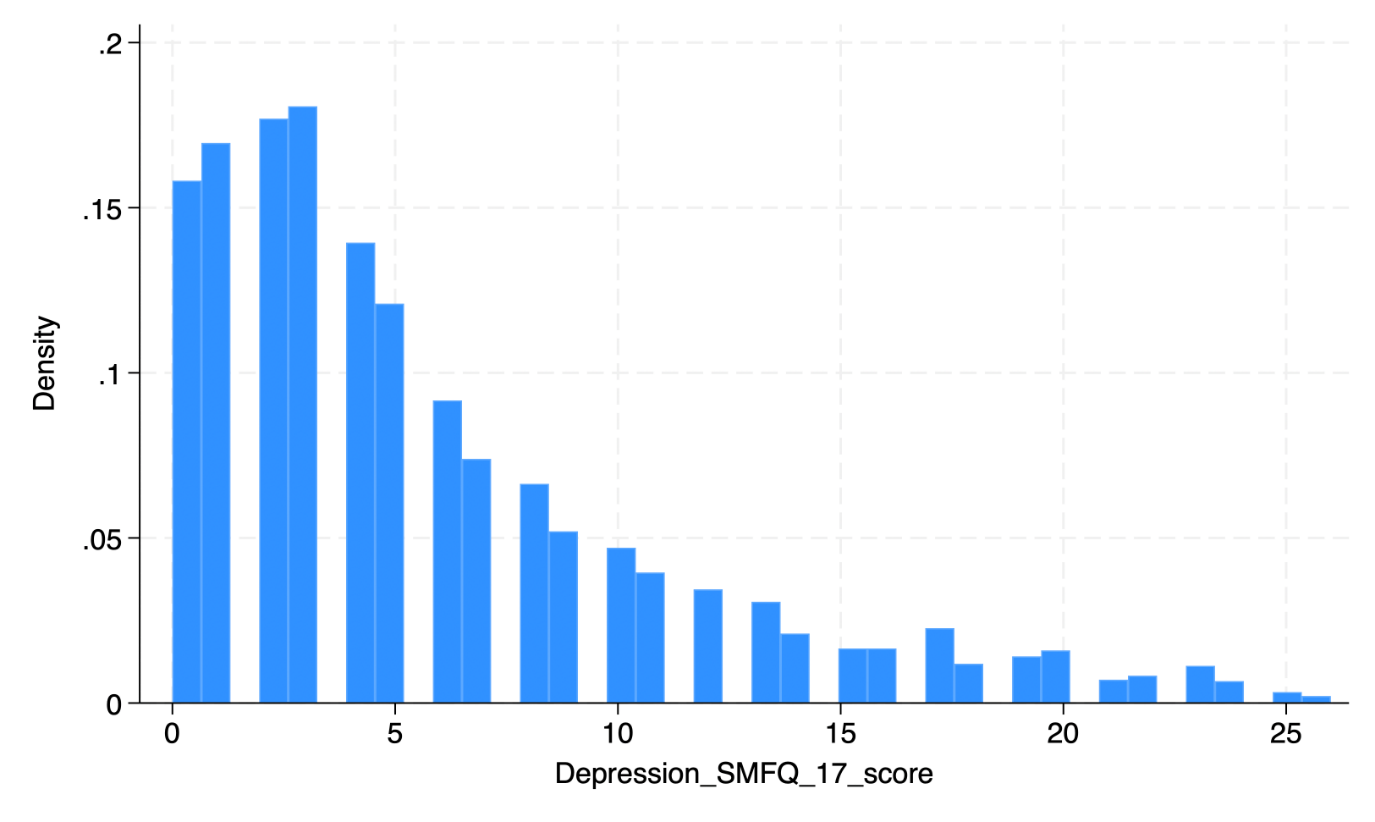

*Supplementary Figure S39.* Plot of population trend for restricted cubic spline model (N knots = 8).

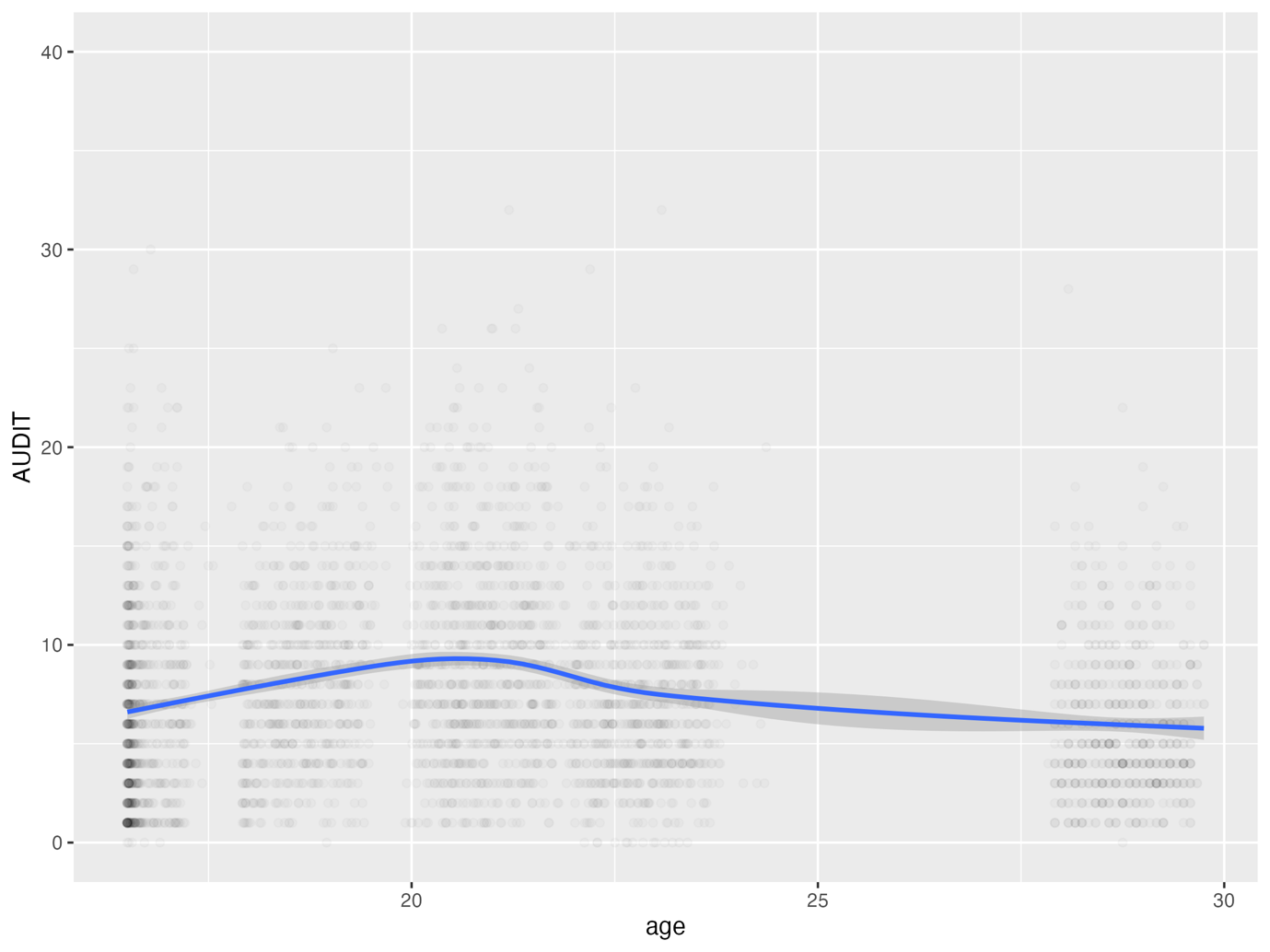

*Supplementary Figure 40.* Flowchart detailing how one of three analytical samples was determined (N=4,035). Please note: This is for individuals who had available data for the autism mean factor score as the exposure.

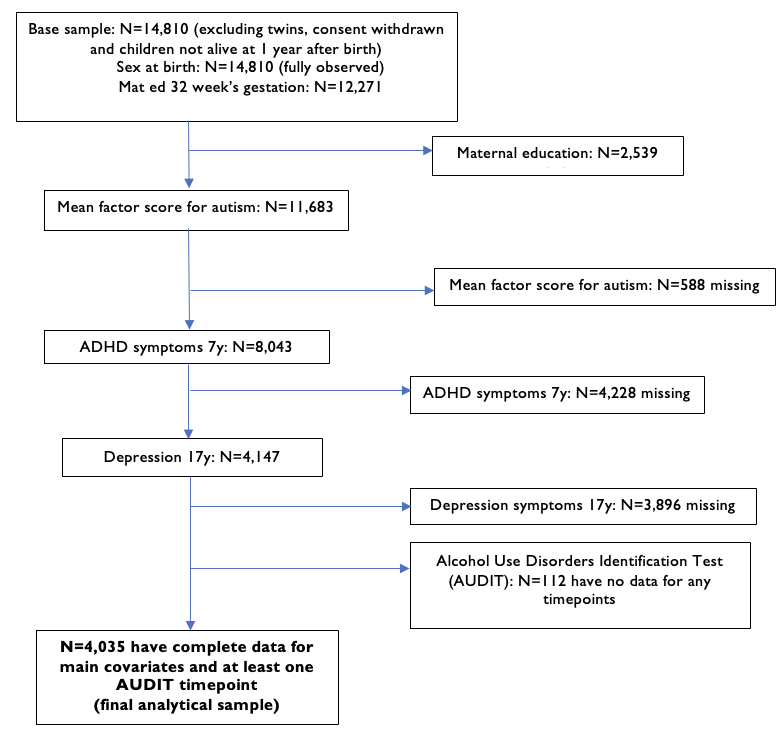

*Supplementary Figure S41.* Flowchart detailing how one of three analytical samples was determined (N=3,771). Please note: This is for individuals who had available data for the Social Communication Disorders Checklist (SCDC) score as the exposure.

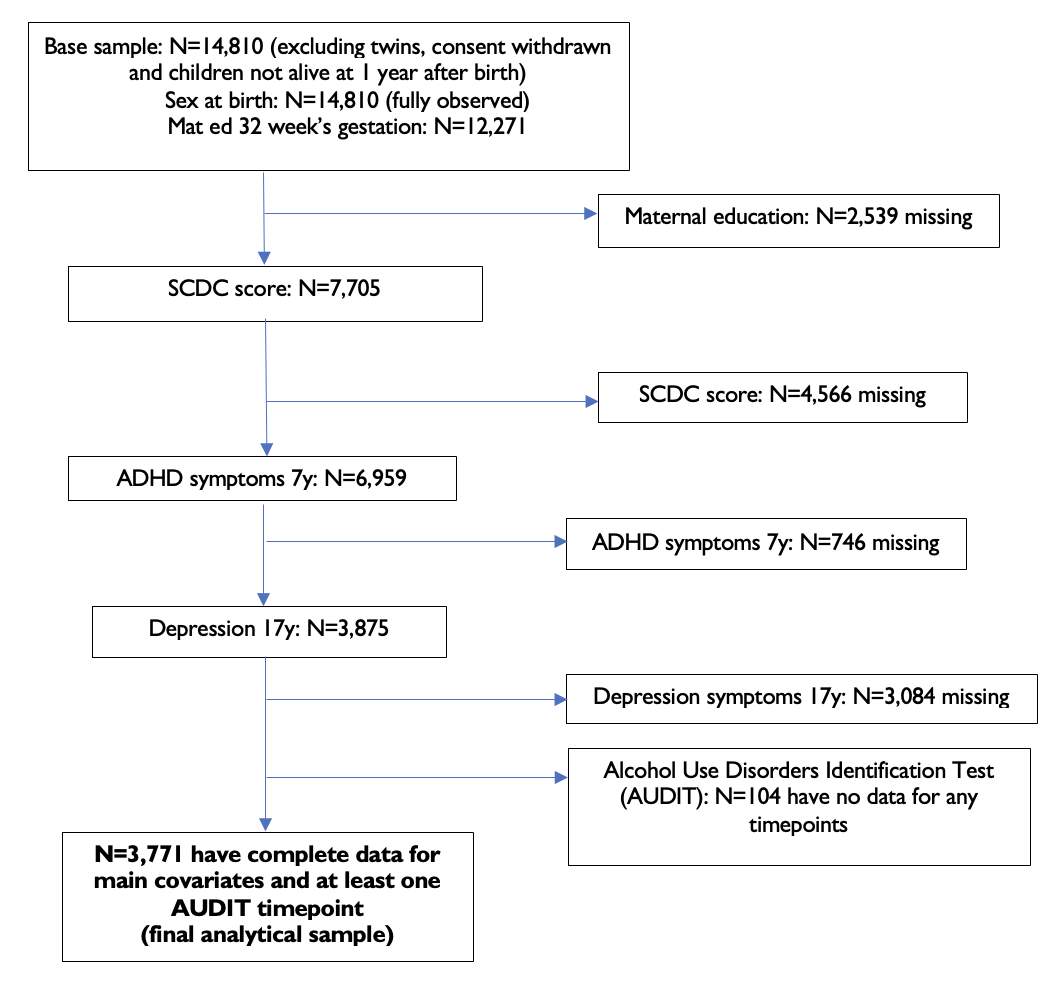

*Supplementary Figure S42.* Flowchart detailing how one of three analytical samples was determined (N=3,250). Please note: This is for individuals who had available data for the polygenic score for autism as the exposure.

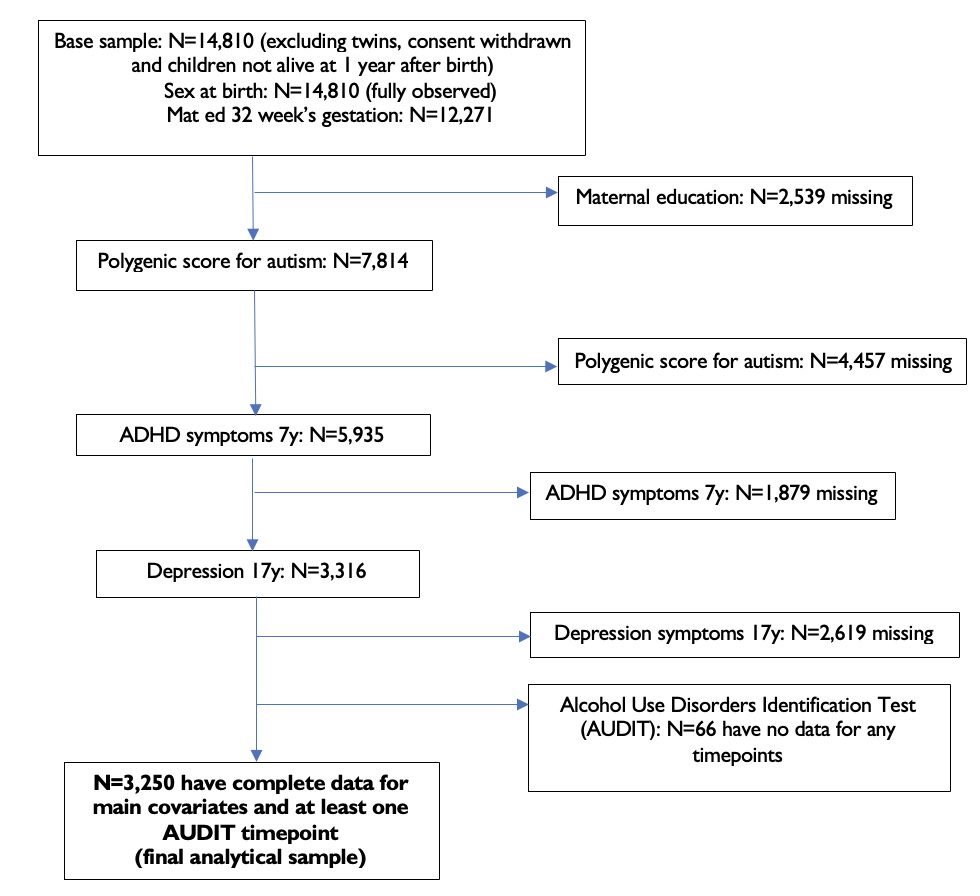
