## Supplementary Tables for "Autistic traits and alcohol consumption through adolescence and young adulthood"

| *Supplementary Table S1.* Phenotypic variance of autism mean factor score explained by autism polygenic score | |
| --- | --- |
| **P-value threshold** | **R²** |
| 0.5 | 0.033 |
| 0.1 | 0.032 |
| 0.05 | 0.032 |
| 0.01 | 0.032 |
| 0.001 | 0.032 |
| 1×10⁻⁴ | 0.032 |
| 1×10⁻⁵ | 0.031 |
| 1×10⁻⁶ | 0.031 |
| 1×10⁻⁷ | 0.031 |
| 1×10⁻⁸ | 0.031 |
| 1×10⁻⁹ | 0.031 |
| 1×10⁻¹⁰ | 0.031 |
| *Supplementary Table S2.* Phenotypic variance of SCDC score at 7 years explained by autism polygenic score |  |
| **P-value threshold** | **R²** |
| 0.1 | 0.022 |
| 0.05 | 0.022 |
| 0.01 | 0.022 |
| 0.001 | 0.022 |
| 1×10⁻⁴ | 0.022 |
| 1×10⁻⁵ | 0.022 |
| 1×10⁻⁶ | 0.026 |
| 1×10⁻⁷ | 0.022 |
| 1×10⁻⁸ | 0.022 |
| 1×10⁻⁹ | 0.022 |
| 1×10⁻¹⁰ | 0.022 |

*Supplementary Table S3*. Definitions of individual effects in a counterfactual mediation framework using Stata plug-in paramed (as described in Rijnhart et al., 2021; Liu et al., 2014).

| **Term** | **Abbreviation** | **Definition** |
| --- | --- | --- |
| Controlled direct effect | CDE | The effect of the exposure on the outcome in the absence of both the mediator and the interaction that operates in the presence of the mediator. |
| Natural direct effect | NDE | The effect of the exposure on the outcome where the value for the mediator is what it would be for each participant when the outcome = 0. |
| Natural indirect effect | NIE | Describes both the effect of the exposure on the outcome and the effect of the exposure on the outcome that operates through the mediator. |
| Pure natural direct effect | PNDE | The effect of the exposure on the outcome while also including the interaction that operates in the presence of the mediator. The PNDE does not capture the mediation effect itself as the mediator is held constant. |
| Pure natural indirect effect | PNIE | Describes the effect of the exposure on the outcome that only operates through the mediator. |
| Total natural direct effect | TNDE | The effect of the exposure on the outcome that includes the interaction effect which operates via the mediator and the interaction effect which does not operate via the mediator. |
| Total natural indirect effect | TNIE | The effect of the exposure on the outcome, both via the mediator and interaction that operates in the presence of the mediator. |
| Marginal total effect | MTE | The sum of the NDE and NIE where the mean value of confounders is used by default by Stata. |
| Total effect | TE | The sum of the NDE and NIE where values for the confounders have been specified. |

*Supplementary Table S4.* AUDIT scores by age at measurement at the mean, 10^th^, 50^th^ (median) and 90^th^ percentiles in those with data for all three exposures

| **Approximate age at completion (in years)** | 17 | 19 | 21 | 23 | 28 |
| --- | --- | --- | --- | --- | --- |
| **Percentile** |  |  |  |  |  |
| Mean | 6.98 | 8.26 | 9.11 | 7.31 | 5.99 |
| 10^th^ | 1 | 3 | 3 | 2 | 2 |
| 50^th^ (median) | 6 | 8 | 9 | 8 | 6 |
| 90th | 14 | 15 | 17 | 15 | 12 |

*Note: Scores are derived from summary statistics of raw data.*

*Supplementary Table S5.* AUDIT scores by age at measurement at the mean, 10^th^, 50^th^ (median) and 90^th^ percentiles in males with data for all three exposures

| **Approximate age at completion (in years)** | 17 | 19 | 21 | 23 | 28 |
| --- | --- | --- | --- | --- | --- |
| **Percentile** |  |  |  |  |  |
| Mean | 6.96 | 8.53 | 9.61 | 8.09 | 6.71 |
| 10^th^ | 1 | 3 | 3 | 2 | 2 |
| 50^th^ (median) | 6 | 7 | 8 | 6 | 5 |
| 90th | 14 | 14 | 16 | 12 | 10 |

*Note: Scores are derived from summary statistics of raw data.*

*Supplementary Table S6.* AUDIT scores by age at measurement at the mean, 10^th^, 50^th^ (median) and 90^th^ percentiles in females with data for all three exposures

| **Approximate age at completion (in years)** | 17 | 19 | 21 | 23 | 28 |
| --- | --- | --- | --- | --- | --- |
| **Percentile** |  |  |  |  |  |
| Mean | 6.99 | 8.10 | 8.79 | 6.88 | 5.57 |
| 10th | 1 | 3 | 3 | 2 | 2 |
| 50^th^ (median) | 6 | 8 | 8 | 7 | 5 |
| 90th | 14 | 14 | 17 | 13 | 11 |

*Note: Scores are derived from summary statistics of raw data.*

*Supplementary Table S7.* Observed vs missing values in all variables (N=14,810; in ascending order of missingness)

| **Variable** | **Label in ALSPAC** | **Missing values** | **Observed values** |
| --- | --- | --- | --- |
| Sex | kz021 | 0 | 14,810 |
| Marital status | a525 | 1,800 | 13,010 |
| Home ownership status | a006 | 1,858 | 12,952 |
| Autism mean factor score | clon207 | 1,904 | 12,906 |
| Highest maternal education 32 weeks' gestation | c645a | 2,539 | 12,271 |
| Maternal age at delivery | e695 | 3,170 | 11,640 |
| Maternal social class | c755 | 4,863 | 9,947 |
| ADHD traits 7y (SDQ subscale score) | Sum of variables kq321, kq329, kq334, kq340 and kq344 | 6,543 | 8,267 |
| SCDC score 7y | Sum of variables kr539 to kr550 | 6,855 | 7,955 |
| Depression symptoms 17y (SMFQ score) | Sum of variables ccs4500 to ccs4515 | 9,806 | 5,004 |

| AUDIT score 17y | Sum of variables ccs3540 to ccs3549 | 10,379 | 4,431 |
| --- | --- | --- | --- |
| AUDIT score 21y | CCU3095 | 10,913 | 3,897 |
| AUDIT score 28y | Sum of variables YPH5660 to YPH5750 | 10,949 | 3,861 |
| AUDIT score 23y | YPB4388a | 10,958 | 3,852 |
| DSM-IV criteria for alcohol abuse | Sum of variables CCU3104, CCU3108, CCU3116, CCU3113, CCU3109 and CCU3117 | 11,023 | 3,787 |
| AUDIT score 19y | Sum of variables cct5030 to cct5039 | 11,719 | 3,091 |
| Parity | b032 | 12,831 | 1,979 |
| Tobacco smoked regularly pre pregnancy | b663 | 13,059 | 1,751 |
| Tobacco smoked in first trimester | b665 | 13,059 | 1,751 |
| Tobacco smoked in last 2 weeks of pregnancy | b667 | 13,059 | 1,751 |

G1 sample recruited through phases 1-III, excluding those who had withdrawn consent, were not alive after 1 year of birth or were second in birth order from the same mother.

*Supplementary Table S8.* Results of multilevel piecewise linear spline and quantile regression models exploring the association between three measures of autism and alcohol use over time (depression excluded).

| **Exposure** | **Mean or percentile** | **Difference in AUDIT score** | **CI 95% (lower, upper)** |
| --- | --- | --- | --- |
| Mean factor score for autism | Mean | 0.79 | 0.34, 1.23 |
|  | 10th | 0.75 | 0.31, 1.18 |
|  | 50^th^ (median) | 0.78 | 0.33, 1.22 |
|  | 90th | 0.73 | 0.37, 1.10 |
| Social Communication Disorders Checklist (SCDC) score at 7y | Mean | 0.02 | -0.03, 0.06 |
|  | 10th | -0.04 | -0.08, 0.01 |
|  | 50^th^ (median) | 0.02 | -0.02, 0.07 |
|  | 90th | 0.13 | 0.05, 0.22 |
| Polygenic score for autism | Mean | 0.001 | -0.13, 0.13 |
|  | 10th | 0.02 | -0.06, 0.11 |
|  | 50^th^ (median) | 0.04 | -0.07, 0.16 |
|  | 90th | 0.09 | -0.11, 0.30 |

Models were adjusted for sex, socioeconomic position and attention deficit hyperactivity disorder (ADHD) traits.

| *Supplementary Table S9.* Parameter estimates from mediation analyses with the autism mean factor score (exposure), depression (mediator) and alcohol use (outcome) while controlling for confounders (sex and socioeconomic position, given through a proxy measure of maternal education). | | | | | |
| --- | --- | --- | --- | --- | --- |
| **Coefficient** | **Observed Coefficient** | **Standard error** | **P value** | **95% Conf. Interval (Lower)** | **95% Conf. Interval (Upper)** |
| CDE | 3.82 | 1.53 | 0.012 | 0.83 | 6.82 |
| NIE | -0.75 | 0.17 | <0.001 | -1.09 | -0.32 |
| MTE | 3.07 | 1.53 | 0.044 | 0.08 | 6.06 |
| This model assumes no exposure-mediator interaction. CDE = Controlled direct effect, NIE = Natural indirect effect, MTE = Marginal total effect. | | | | | |

| *Supplementary Table S10.* Parameter estimates from mediation analyses with the autism mean factor score (exposure), depression (mediator) and alcohol use (outcome) while controlling for confounders (sex and socioeconomic position, given through a proxy measure of maternal education). | | | | | |
| --- | --- | --- | --- | --- | --- |
| **Coefficient** | **Observed Coefficient** | **Standard error** | **P value** | **95% Conf. Interval (Lower)** | **95% Conf. Interval (Upper)** |
| CDE | 3.85 | 1.53 | 0.012 | 0.84 | 6.85 |
| NDE | 3.51 | 2.05 | 0.087 | -0.51 | 7.53 |
| NIE | -0.69 | 0.31 | 0.028 | -1.31 | -0.07 |
| MTE | 2.82 | 1.89 | 0.135 | -0.88 | 6.51 |
| This model assumes an exposure-mediator interaction. CDE = Controlled direct effect, NDE = Natural direct effect, NIE = Natural indirect effect, MTE = Marginal total effect. | | | | | |
| *Supplementary Table S11.* Parameter estimates from mediation analyses with the autism mean factor score (exposure), depression (mediator) and alcohol use (outcome) while controlling for confounders (sex and socioeconomic position, given through a proxy measure of maternal education). | | | | | |
| **Coefficient** | **Observed Coefficient** | **Standard error** | **P value** | **95% Conf. Interval (Lower)** | **95% Conf. Interval (Upper)** |
| CDE | 2.14 | 1.79 | 0.230 | -1.36 | 5.66 |
| NDE | 0.23 | 2.00 | 0.907 | -3.68 | 4.14 |
| NIE | -0.09 | 0.20 | 0.661 | -0.48 | 0.30 |
| MTE | 0.15 | 1.86 | 0.937 | -3.50 | 3.79 |
| This model assumes an exposure-mediator interaction. The mediator has been binarised to effectively remove depression from the population. CDE = Controlled direct effect, NDE = Natural direct effect, NIE = Natural indirect effect, MTE = Marginal total effect. | | | | | |
| *Supplementary Table S12.* Parameter estimates from mediation analyses with the Social Communication Disorders Checklist Score (SCDC, exposure), depression (mediator) and alcohol use (outcome) while controlling for confounders (sex and socioeconomic position, given through a proxy measure of maternal education). | | | | | |
| **Coefficient** | **Observed Coefficient** | **Standard error** | **P value** | **95% Conf. Interval (Lower)** | **95% Conf. Interval (Upper)** |
| CDE | -0.55 | 0.85 | 0.517 | -2.22 | 1.12 |
| NIE | 0.40 | 0.10 | <0.001 | 0.20 | 0.59 |
| TE | -0.15 | 0.85 | 0.856 | -1.82 | 1.51 |
| This model assumes no exposure-mediator interaction. CDE = Controlled direct effect, NIE = Natural indirect effect, MTE = Marginal total effect. | | | | | |
| *Supplementary Table S13.* Parameter estimates from mediation analyses with the Social Communication Disorders Checklist Score (SCDC, exposure), depression (mediator) and alcohol use (outcome) while controlling for confounders (sex and socioeconomic position, given through a proxy measure of maternal education). | | | | | |
| **Coefficient** | **Observed Coefficient** | **Standard error** | **P value** | **95% Conf. Interval (Lower)** | **95% Conf. Interval (Upper)** |
| CDE | -0.44 | 0.86 | 0.599 | -2.13 | 1.23 |
| NDE | -0.31 | 0.88 | 0.720 | -2.03 | 1.40 |
| NIE | -0.20 | 0.52 | 0.702 | -1.21 | 0.81 |
| MTE | -0.51 | 0.90 | 0.571 | -2.28 | 1.26 |
| This model assumes an exposure-mediator interaction. CDE = Controlled direct effect, NDE = Natural direct effect, NIE = Natural indirect effect, MTE = Marginal total effect. | | | | | |
| *Supplementary Table S14.* Parameter estimates from mediation analyses with the Social Communication Disorders Checklist Score (SCDC, exposure), depression (mediator) and alcohol use (outcome) while controlling for confounders (sex and socioeconomic position, given through a proxy measure of maternal education). | | | | | |
| **Coefficient** | **Observed Coefficient** | **Standard error** | **P value** | **95% Conf. Interval (Lower)** | **95% Conf. Interval (Upper)** |
| CDE | -0.48 | 1.06 | 0.650 | -2.55 | 1.59 |
| NDE | 0.16 | 0.75 | 0.827 | -1.31 | 1.64 |
| NIE | 0.42 | 0.40 | 0.294 | -0.36 | 1.20 |
| MTE | 0.58 | 0.90 | 0.518 | -1.18 | 2.34 |

This model assumes an exposure-mediator interaction. The mediator has been binarised to effectively remove depression from the population. CDE = Controlled direct effect, NDE = Natural direct effect, NIE = Natural indirect effect, MTE = Marginal total effect.

| *Supplementary Table S15.* Parameter estimates from mediation analyses with polygenic score for autism (exposure), depression (mediator) and alcohol use (outcome) while controlling for confounders (sex and socioeconomic position, given through a proxy measure of maternal education). | | | | | |
| --- | --- | --- | --- | --- | --- |
| **Coefficient** | **Observed Coefficient** | **Standard error** | **P value** | **95% Conf. Interval (Lower)** | **95% Conf. Interval (Upper)** |
| CDE | 0.51 | 1.07 | 0.634 | -1.60 | 2.62 |
| NIE | 0.02 | 0.10 | 0.876 | -0.18 | 0.21 |
| MTE | 0.53 | 1.08 | 0.625 | -1.59 | 2.65 |
| This model assumes no exposure-mediator interaction. CDE = Controlled direct effect, NIE = Natural indirect effect, MTE = Marginal total effect. | | | | | |
| *Supplementary Table S16.* Parameter estimates from mediation analyses with polygenic score for autism (exposure), depression (mediator) and alcohol use (outcome) while controlling for confounders (sex and socioeconomic position, given through a proxy measure of maternal education). | | | | | |
| **Coefficient** | **Observed Coefficient** | **Standard error** | **P value** | **95% Conf. Interval (Lower)** | **95% Conf. Interval (Upper)** |
| CDE | 0.54 | 1.08 | 0.617 | -1.57 | 2.64 |
| NDE | 0.64 | 1.09 | 0.552 | -1.48 | 2.77 |
| NIE | -0.01 | 0.06 | 0.880 | -0.12 | 0.10 |
| MTE | 0.64 | 1.08 | 0.556 | -1.48 | 2.75 |
| This model assumes an exposure-mediator interaction. CDE = Controlled direct effect, NDE = Natural direct effect, NIE = Natural indirect effect, MTE = Marginal total effect. | | | | | |
| *Supplementary Table S17.* Parameter estimates from mediation analyses with polygenic score for autism (exposure), depression (mediator) and alcohol use (outcome) while controlling for confounders (sex and socioeconomic position, given through a proxy measure of maternal education). | | | | | |
| **Coefficient** | **Observed Coefficient** | **Standard error** | **P value** | **95% Conf. Interval (Lower)** | **95% Conf. Interval (Upper)** |
| CDE | 2.56 | 1.27 | 0.045 | 0.06 | 5.06 |
| NDE | -0.52 | 1.19 | 0.663 | -2.86 | 1.82 |
| NIE | 0.52 | 0.44 | 0.244 | -0.35 | 1.39 |
| MTE | -0.003 | 1.01 | 0.998 | -1.99 | 1.98 |
| This model assumes an exposure-mediator interaction. The mediator has been binarised to effectively remove depression from the population. CDE = Controlled direct effect, NDE = Natural direct effect, NIE = Natural indirect effect, MTE = Marginal total effect. | | | | | |

| *Supplementary Table S18.* Output of final baseline multilevel linear models for each exposure where the outcome (AUDIT) has been log transformed | | |
| --- | --- | --- |
| **Exposure** | **Difference in AUDIT score** | **CI 95% (lower, upper)** |
| Mean factor score for autism | 0.20 | 0.14, 0.25 |
| Social Communication Disorders Checklist (SCDC) score at 7y | -0.01 | -0.01, -0.001 |
| Polygenic score for autism | -0.03 | -0.16, 0.11 |

Models were adjusted for sex, socioeconomic position, depression symptoms and attention deficit hyperactivity disorder (ADHD) traits.

*Supplementary Table 19.* Results of multilevel piecewise linear spline and quantile regression models exploring the association between three measures of autism and alcohol use over time (depression excluded).

| **Exposure** | **Mean or percentile** | **Difference in AUDIT score** | **CI 95% (lower, upper)** |
| --- | --- | --- | --- |
| Mean factor score for autism | Mean | 0.81 | 0.42, 1.20 |
|  | 10^th^ | 0.74 | 0.33, 1.16 |
|  | 50^th^ (median) | 0.77 | 0.37, 1.17 |
|  | 90th | 0.73 | 0.25, 1.21 |
| Social Communication Disorders Checklist (SCDC) score at 7y | Mean | 0.01 | -0.03, 0.06 |
|  | 10^th^ | -0.02 | -0.06, 0.01 |
|  | 50^th^ (median) | 0.01 | -0.04, 0.06 |
|  | 90th | 0.12 | 0.02, 0.22 |
| Polygenic score for autism | Mean | -0.10 | -1.08, 0.88 |
|  | 10th | 0.08 | -0.87, 1.02 |
|  | 50^th^ (median) | 0.09 | -0.83, 1.00 |
|  | 90th | 0.09 | -1.05, 1.22 |

Models were adjusted for sex, socioeconomic position, and attention deficit hyperactivity disorder (ADHD) traits.

*Supplementary Table 20.* Results of multilevel piecewise linear spline and quantile regression models exploring the association between three measures of autism and alcohol use over time (with exposure*age interactions included).

| **Exposure** | **Mean or percentile** | **Difference in AUDIT score** | **CI 95% (lower, upper)** |
| --- | --- | --- | --- |
| Mean factor score for autism | Mean | 19.54 | 2.33, 36.76 |
|  | 10^th^ | 20.57 | 2.99, 38.14 |
|  | 50^th^ (median) | 20.57 | 0.50, 40.64 |
|  | 90th | 20.57 | 0.61, 40.53 |
| Social Communication Disorders Checklist (SCDC) score at 7y | Mean | 0.07 | -1.76, 1.91 |
|  | 10^th^ | 0.62 | -1.52, 1.44 |
|  | 50^th^ (median) | 0.62 | -1.56, 2.81 |
|  | 90th | 0.63 | -1.17, 2.44 |
| Polygenic score for autism | Mean | 17.47 | -24.21, 59.15 |
|  | 10th | 28.48 | -13.79, 70.75 |
|  | 50^th^ (median) | 28.49 | -12.57, 69.54 |
|  | 90th | 28.49 | -24.63, 81.60 |

Models were adjusted for sex, socioeconomic position, depression symptoms and attention deficit hyperactivity disorder (ADHD) traits.

*Supplementary Table 21.* Results of multilevel piecewise linear spline and quantile regression models exploring the association between three measures of autism and alcohol use over time (with exposure*age interactions included and depression excluded).

| **Exposure** | **Mean or percentile** | **Difference in AUDIT score** | **CI 95% (lower, upper)** |
| --- | --- | --- | --- |
| Mean factor score for autism | Mean | 17.53 | 0.27, 34.79 |
|  | 10^th^ | 18.47 | -1.28, 38.22 |
|  | 50^th^ (median) | 18.47 | -3.35, 40.29 |
|  | 90th | 18.48 | 0.42, 36.54 |
| Social Communication Disorders Checklist (SCDC) score at 7y | Mean | 0.02 | -1.82, 1.87 |
|  | 10^th^ | 0.47 | -1.47, 2.41 |
|  | 50^th^ (median) | 0.46 | -2.58, 3.51 |
|  | 90th | 0.49 | -2.01, 3.00 |
| Polygenic score for autism | Mean | 19.68 | -22.40, 61.75 |
|  | 10th | 34.41 | -17.75, 86.58 |
|  | 50^th^ (median) | 34.41 | -15.28, 84.11 |
|  | 90th | 34.42 | -2.15, 70.98 |

Models were adjusted for sex, socioeconomic position and attention deficit hyperactivity disorder (ADHD) traits.

*Supplementary Table S22.* Comparison of model fit measured using the Akaike Information Criterion (AIC) across all models

| **Autism measure used** | **Specific model** | **AIC** |
| --- | --- | --- |
| Autism mean factor score | Final baseline model | 75498.25 |
|  | Final baseline model (depression removed) | 75666.32 |
|  | Final baseline model with exposure*age interactions included | 75496.66 |
|  | Final baseline model with exposure*age interactions included (depression removed) | 75668.93 |
|  | Final baseline model (outcome log transformed) | 21960.38 |
|  | Final baseline model (LQMM, 10th percentile) | 75888.25 |
|  | Final baseline model (LQMM, 50th percentile) | 76059.00 |
|  | Final baseline model (LQMM, 90th percentile) | 82310.63 |
|  | Final baseline model (LQMM, 10th percentile, depression removed) | 75012.04 |
|  | Final baseline model (LQMM, 50th percentile, depression removed) | 75230.93 |
|  | Final baseline model (LQMM, 90th percentile, depression removed) | 80103.44 |
|  | Final baseline model exposure*age interactions included (LQMM, 10th percentile) | 75996.39 |
|  | Final baseline model exposure*age interactions included (LQMM, 50th percentile) | 75245.99 |
|  | Final baseline model exposure*age interactions included (LQMM, 90th percentile) | 81572.35 |
|  | Final baseline model exposure*age interactions included (LQMM, 10th percentile, depression removed) | 74986.56 |
|  | Final baseline model exposure*age interactions included (LQMM, 50th percentile, depression removed) | 75441.17 |
|  | Final baseline model exposure*age interactions included (LQMM, 90th percentile, depression removed) | 80150.52 |
| SCDC score 7y | Final baseline model | 71043.02 |
|  | Final baseline model (depression removed) | 71196.92 |
|  | Final baseline model with exposure*age interactions included | 71045.72 |
|  | Final baseline model with exposure*age interactions included (depression removed) | 71196.56 |
|  | Final baseline model (outcome log transformed) | 20621.89 |
|  | Final baseline model (LQMM, 10th percentile) | 71622.29 |
|  | Final baseline model (LQMM, 50th percentile) | 71292.74 |
|  | Final baseline model (LQMM, 90th percentile) | 77048.32 |
|  | Final baseline model (LQMM, 10th percentile, depression removed) | 70801.06 |
|  | Final baseline model (LQMM, 50th percentile, depression removed) | 71053.56 |
|  | Final baseline model (LQMM, 90th percentile, depression removed) | 75331.93 |
|  | Final baseline model exposure*age interactions included (LQMM, 10th percentile) | 71507.73 |
|  | Final baseline model exposure*age interactions included (LQMM, 50th percentile) | 72346.07 |
|  | Final baseline model exposure*age interactions included (LQMM, 90th percentile) | 77389.75 |
|  | Final baseline model exposure*age interactions included (LQMM, 10th percentile, depression removed) | 71367.38 |
|  | Final baseline model exposure*age interactions included (LQMM, 50th percentile, depression removed) | 70774.51 |
|  | Final baseline model exposure*age interactions included (LQMM, 90th percentile, depression removed) | 76460.33 |
| Polygenic score for autism | Final baseline model | 61428.49 |
|  | Final baseline model (depression removed) | 61573.26 |
|  | Final baseline model with exposure*age interactions included | 61433.56 |
|  | Final baseline model with exposure*age interactions included (depression removed) | 61578.02 |
|  | Final baseline model (outcome log transformed) | 17618.76 |
|  | Final baseline model (LQMM, 10th percentile) | 62152.24 |
|  | Final baseline model (LQMM, 50th percentile) | 61810.6 |
|  | Final baseline model (LQMM, 90th percentile) | 68754.47 |
|  | Final baseline model (LQMM, 10th percentile, depression removed) | 61236.58 |
|  | Final baseline model (LQMM, 50th percentile, depression removed) | 61408.54 |
|  | Final baseline model (LQMM, 90th percentile, depression removed) | 65170.91 |
|  | Final baseline model exposure*age interactions included (LQMM, 10th percentile) | 61954.61 |
|  | Final baseline model exposure*age interactions included (LQMM, 50th percentile) | 61679.62 |
|  | Final baseline model exposure*age interactions included (LQMM, 90th percentile) | 68547.04 |
|  | Final baseline model exposure*age interactions included (LQMM, 10th percentile, depression removed) | 61340.61 |
|  | Final baseline model exposure*age interactions included (LQMM, 50th percentile, depression removed) | 61444.86 |
|  | Final baseline model exposure*age interactions included (LQMM, 90th percentile, depression removed) | 65345.76 |

*Supplementary Table S23.* Overall base trajectories of mean alcohol use

| **Exposure** |  | **Coefficient** | **CI 95% (lower, upper)** |
| --- | --- | --- | --- |
| Mean factor score for autism | Intercept | -8.63 | -25.73, 8.46 |
|  | Spline 1 | 0.82 | -0.19, 1.84 |
|  | Spline 2 | 1.24 | 0.84, 1.63 |
|  | Spline 3 | -0.62 | -0.93, -0.31 |
|  | Spline 4 | -0.16 | -0.49, 0.18 |
|  | Spline 5 | -0.66 | -1.56, 0.24 |
| Social Communication Disorders Checklist (SCDC) score at 7y | Intercept | -5.30 | -22.86, 12.26 |
|  | Spline 1 | 0.62 | -0.42, 1.66 |
|  | Spline 2 | 1.30 | 0.89, 1.71 |
|  | Spline 3 | -0.62 | -0.93, -0.30 |
|  | Spline 4 | -0.17 | -0.52, 0.17 |
|  | Spline 5 | -0.65 | -1.58, 0.29 |
| Polygenic score for autism | Intercept | -11.82 | -30.56, 6.91 |
|  | Spline 1 | 1.00 | -0.11, 2.12 |
|  | Spline 2 | 1.23 | 0.79, 1.67 |
|  | Spline 3 | -0.67 | -1.01, -0.33 |
|  | Spline 4 | -0.14 | -0.50, 0.22 |
|  | Spline 5 | -0.46 | -1.43, 0.51 |

Models were adjusted for sex, socioeconomic position, depression symptoms and attention deficit hyperactivity disorder (ADHD) traits.

*Supplementary Table S24.* Overall base trajectories of 10th percentile alcohol use

| **Exposure** |  | **Coefficient** | **CI 95% (lower, upper)** | |
| --- | --- | --- | --- | --- |
| Mean factor score for autism | Intercept | -7.75 | | -30.94, 15.45 |
|  | Spline 1 | 0.53 | | -0.86, 1.93 |
|  | Spline 2 | 1.16 | | 0.60. 1.72 |
|  | Spline 3 | -0.60 | | -0.94, -0.26 |
|  | Spline 4 | -0.01 | | -0.50, 0.48 |
|  | Spline 5 | -1.00 | | -2.21, 0.25 |
| Social Communication Disorders Checklist (SCDC) score at 7y | Intercept | -5.26 | | -22.55, 12.04 |
|  | Spline 1 | 0.38 | | -0.64, 1.41 |
|  | Spline 2 | 1.19 | | 0.76, 1.63 |
|  | Spline 3 | -0.59 | | -0.90, -0.28 |
|  | Spline 4 | -0.04 | | -0.45, 0.36 |
|  | Spline 5 | -0.94 | | -2.16, 0.28 |
| Polygenic score for autism | Intercept | -12.26 | | -32.58, 8.03 |
|  | Spline 1 | 0.80 | | -0.40, 2.01 |
|  | Spline 2 | 1.10 | | 0.63, 1.56 |
|  | Spline 3 | -0.67 | | -1.03, -0.30 |
|  | Spline 4 | -0.01 | | -0.44, 0.42 |
|  | Spline 5 | -0.70 | | -1.81, 0.41 |

Models were adjusted for sex, socioeconomic position, depression symptoms and attention deficit hyperactivity disorder (ADHD) traits.

*Supplementary Table S25.* Overall base trajectories of 50th percentile alcohol use

| **Exposure** |  | **Coefficient** | **CI 95% (lower, upper)** |
| --- | --- | --- | --- |
| Mean factor score for autism | Intercept | -7.73 | -27.57, 12.10 |
|  | Spline 1 | 0.75 | -0.42, 1.93 |
|  | Spline 2 | 1.18 | 0.70, 1.66 |
|  | Spline 3 | -0.61 | -0.94, -0.27 |
|  | Spline 4 | -0.003 | -0.41, 0.41 |
|  | Spline 5 | -0.98 | -2.03, 0.07 |
| Social Communication Disorders Checklist (SCDC) score at 7y | Intercept | -5.24 | -23.66, 13.17 |
|  | Spline 1 | 0.61 | -0.47, 1.68 |
|  | Spline 2 | 1.22 | 0.78, 1.65 |
|  | Spline 3 | -0.59 | -0.96, -0.22 |
|  | Spline 4 | -0.03 | -0.43, 0.37 |
|  | Spline 5 | -0.94 | -2.02, 0.13 |
| Polygenic score for autism | Intercept | -12.26 | -32.31, |
|  | Spline 1 | 1.02 | -0.18, |
|  | Spline 2 | 1.12 | 0.64, |
|  | Spline 3 | -0.66 | -1.02, |
|  | Spline 4 | 0.001 | -0.53, |
|  | Spline 5 | -0.70 | -2.14 |

Models were adjusted for sex, socioeconomic position, depression symptoms and attention deficit hyperactivity disorder (ADHD) traits.

*Supplementary Table S26.* Overall base trajectories of 90th percentile alcohol use

| **Exposure** |  | **Coefficient** | **CI 95% (lower, upper)** |
| --- | --- | --- | --- |
| Mean factor score for autism | Intercept | -7.71 | -26.44, 11.01 |
|  | Spline 1 | 1.00 | -0.11, 2.12 |
|  | Spline 2 | 1.16 | 0.68, 1.63 |
|  | Spline 3 | -0.65 | -0.96, -0.34 |
|  | Spline 4 | -0.01 | -0.41, 0.39 |
|  | Spline 5 | -0.98 | -2.07, 0.10 |
| Social Communication Disorders Checklist (SCDC) score at 7y | Intercept | -5.22 | -25.87, 15.42 |
|  | Spline 1 | 0.83 | -0.39, 2.06 |
|  | Spline 2 | 1.20 | 0.67, 1.73 |
|  | Spline 3 | -0.63 | -0.99, -0.27 |
|  | Spline 4 | -0.04 | -0.47, 0.39 |
|  | Spline 5 | -0.95 | -2.12, 0.23 |
| Polygenic score for autism | Intercept | -12.24 | -29.75, 5.26 |
|  | Spline 1 | 1.31 | 0.27, 2.35 |
|  | Spline 2 | 1.10 | 0.65, 1.56 |
|  | Spline 3 | -0.69 | -1.06, -0.32 |
|  | Spline 4 | 0.01 | -0.39, 0.41 |
|  | Spline 5 | -0.70 | -1.82, 0.43 |

Models were adjusted for sex, socioeconomic position, depression symptoms and attention deficit hyperactivity disorder (ADHD) traits.

*Supplementary Table S27.* Overall base trajectories of mean alcohol use (with depression excluded)

| **Exposure** |  | **Coefficient** | **CI 95% (lower, upper)** |
| --- | --- | --- | --- |
| Mean factor score for autism | Intercept | -5.40 | -22.25, 11.45 |
|  | Spline 1 | 0.67 | -0.33, 1.67 |
|  | Spline 2 | 1.15 | 0.76, 1.55 |
|  | Spline 3 | -0.65 | -0.95, -0.35 |
|  | Spline 4 | -0.12 | -0.45, 0.21 |
|  | Spline 5 | -0.73 | -1.61, 0.15 |
| Social Communication Disorders Checklist (SCDC) score at 7y | Intercept | -3.18 | -20.46, 14.09 |
|  | Spline 1 | 0.54 | -0.49, 1.56 |
|  | Spline 2 | 1.20 | 0.80, 1.60 |
|  | Spline 3 | -0.66 | -0.96, -0.35 |
|  | Spline 4 | -0.12 | -0.46, 0.21 |
|  | Spline 5 | -0.73 | -1.64, 0.17 |
| Polygenic score for autism | Intercept | -7.96 | -26.43, 10.52 |
|  | Spline 1 | 0.82 | -0.28, 1.92 |
|  | Spline 2 | 1.15 | 0.72, 1.58 |
|  | Spline 3 | -0.69 | -1.02, -0.36 |
|  | Spline 4 | -0.13 | -0.49, 0.22 |
|  | Spline 5 | -0.54 | -1.48, 0.41 |

Models were adjusted for sex, socioeconomic position, and attention deficit hyperactivity disorder (ADHD) traits.

*Supplementary Table S28.* Overall base trajectories of 10th percentile alcohol use (with depression excluded)

| **Exposure** |  | **Coefficient** | **CI 95% (lower, upper)** |
| --- | --- | --- | --- |
| Mean factor score for autism | Intercept | -4.31 | -23.66, 15.05 |
|  | Spline 1 | 0.38 | -0.78, 1.53 |
|  | Spline 2 | 1.07 | 0.55, 1.58 |
|  | Spline 3 | -0.55 | -0.94, -0.17 |
|  | Spline 4 | 0.09 | -0.36, 0.53 |
|  | Spline 5 | -0.95 | -2.17, 0.27 |
| Social Communication Disorders Checklist (SCDC) score at 7y | Intercept | -3.09 | -22.42, 16.25 |
|  | Spline 1 | 0.30 | -0.85, 1.46 |
|  | Spline 2 | 1.09 | 0.67, 1.50 |
|  | Spline 3 | -0.55 | -0.84, -0.26 |
|  | Spline 4 | 0.05 | -0.37, 0.48 |
|  | Spline 5 | -0.89 | -1.98, 0.21 |
| Polygenic score for autism | Intercept | -7.80 | -30.31, 14.72 |
|  | Spline 1 | 0.59 | -0.76, 1.94 |
|  | Spline 2 | 1.01 | 0.44, 1.58 |
|  | Spline 3 | -0.60 | -0.99, -0.21 |
|  | Spline 4 | 0.05 | -0.38, 0.47 |
|  | Spline 5 | -0.69 | -1.88, 0.50 |

Models were adjusted for sex, socioeconomic position and attention deficit hyperactivity disorder (ADHD) traits.

*Supplementary Table S29.* Overall base trajectories of 50th percentile alcohol use (with depression excluded)

| **Exposure** |  | **Coefficient** | **CI 95% (lower, upper)** |
| --- | --- | --- | --- |
| Mean factor score for autism | Intercept | -4.29 | -23.34, 14.76 |
|  | Spline 1 | 0.60 | -0.54, 1.74 |
|  | Spline 2 | 1.10 | 0.66, 1.54 |
|  | Spline 3 | -0.63 | -0.96, -0.29 |
|  | Spline 4 | -0.02 | -0.43, 0.39 |
|  | Spline 5 | -1.00 | -2.12, 0.12 |
| Social Communication Disorders Checklist (SCDC) score at 7y | Intercept | -3.07 | -24.79, 18.65 |
|  | Spline 1 | 0.52 | -0.77, 1.81 |
|  | Spline 2 | 1.12 | 0.59, 1.65 |
|  | Spline 3 | -0.62 | -1.01, -0.23 |
|  | Spline 4 | -0.03 | -0.46, 0.40 |
|  | Spline 5 | -0.93 | -2.05, 0.20 |
| Polygenic score for autism | Intercept | -7.78 | -24.99, 9.43 |
|  | Spline 1 | 0.80 | -0.22, 1.83 |
|  | Spline 2 | 1.05 | 0.56, 1.54 |
|  | Spline 3 | -0.65 | -0.97, -0.34 |
|  | Spline 4 | -0.04 | -0.46, 0.39 |
|  | Spline 5 | -0.73 | -1.96, 0.50 |

Models were adjusted for sex, socioeconomic position and attention deficit hyperactivity disorder (ADHD) traits.

*Supplementary Table S30.* Overall base trajectories of 90th percentile alcohol use (with depression excluded)

| **Exposure** |  | **Coefficient** | **CI 95% (lower, upper)** |
| --- | --- | --- | --- |
| Mean factor score for autism | Intercept | -4.26 | -22.73, 14.21 |
|  | Spline 1 | 0.86 | -0.24, 1.97 |
|  | Spline 2 | 1.08 | 0.64, 1.52 |
|  | Spline 3 | -0.70 | -1.07, -0.33 |
|  | Spline 4 | -0.04 | -0.46, 0.37 |
|  | Spline 5 | -0.99 | -2.19, 0.21 |
| Social Communication Disorders Checklist (SCDC) score at 7y | Intercept | -3.05 | -20.00, 13.91 |
|  | Spline 1 | 0.79 | -0.22, 1.79 |
|  | Spline 2 | 1.12 | 0.62, 1.61 |
|  | Spline 3 | -0.70 | -1.03, -0.36 |
|  | Spline 4 | -0.06 | -0.46, 0.35 |
|  | Spline 5 | -0.93 | -1.91, 0.06 |
| Polygenic score for autism | Intercept | -7.75 | -26.86, 11.36 |
|  | Spline 1 | 1.07 | -0.07, 2.21 |
|  | Spline 2 | 1.04 | 0.56, 1.52 |
|  | Spline 3 | -0.75 | -1.08, -0.41 |
|  | Spline 4 | -0.06 | -0.42, 0.30 |
|  | Spline 5 | -0.72 | -1.73, 0.28 |

Models were adjusted for sex, socioeconomic position and attention deficit hyperactivity disorder (ADHD) traits.

*Supplementary Table S31.* Exposure*age interactions and mean alcohol use

| **Exposure** |  | **Coefficient** | **CI 95% (lower, upper)** |
| --- | --- | --- | --- |
| Mean factor score for autism | Intercept | -9.52 | -26.61, 7.57 |
|  | Spline 1 | 0.87 | -0.14, 1.89 |
|  | Spline 2 | 1.23 | 0.83, 1.64 |
|  | Spline 3 | -0.62 | -0.93, -0.31 |
|  | Spline 4 | -0.16 | -0.50, 0.18 |
|  | Spline 5 | -0.64 | -1.54, 0.26 |
| Social Communication Disorders Checklist (SCDC) score at 7y | Intercept | -5.33 | -22.96, 12.29 |
|  | Spline 1 | 0.62 | -0.43, 1.67 |
|  | Spline 2 | 1.30 | 0.89, 1.71 |
|  | Spline 3 | -0.60 | -0.91, -0.28 |
|  | Spline 4 | -0.17 | -0.52, 0.18 |
|  | Spline 5 | -0.67 | -1.61, 0.27 |
| Polygenic score for autism | Intercept | -11.96 | -30.69, 6.78 |
|  | Spline 1 | 1.01 | -0.10, 2.12 |
|  | Spline 2 | 1.22 | 0.78, 1.66 |
|  | Spline 3 | -0.66 | -0.99, -0.32 |
|  | Spline 4 | -0.15 | -0.51, 0.22 |
|  | Spline 5 | -0.46 | -1.43, 0.51 |

Models were adjusted for sex, socioeconomic position, depression symptoms and attention deficit hyperactivity disorder (ADHD) traits.

*Supplementary Table S32.* Exposure*age interactions and 10th percentile alcohol use

| **Exposure** |  | **Coefficient** | **CI 95% (lower, upper)** |
| --- | --- | --- | --- |
| Mean factor score for autism | Intercept | -8.73 | -30.21, 12.75 |
|  | Spline 1 | 0.59 | -0.69, 1.87 |
|  | Spline 2 | 1.15 | 0.71, 1.59 |
|  | Spline 3 | -0.6 | -0.91, -0.30 |
|  | Spline 4 | -0.01 | -0.46, 0.44 |
|  | Spline 5 | -0.97 | -2.15, 0.21 |
| Social Communication Disorders Checklist (SCDC) score at 7y | Intercept | -5.70 | -25.13, 13.73 |
|  | Spline 1 | 0.41 | -0.73, 1.55 |
|  | Spline 2 | 1.18 | 0.78, 1.59 |
|  | Spline 3 | -0.55 | -0.96, -0.14 |
|  | Spline 4 | -0.06 | -0.55, 0.43 |
|  | Spline 5 | -0.90 | -2.24, 0.43 |
| Polygenic score for autism | Intercept | -12.39 | -32.20, 7.42 |
|  | Spline 1 | 0.81 | -0.37, 1.99 |
|  | Spline 2 | 1.09 | 0.63, 1.54 |
|  | Spline 3 | -0.65 | -1.07, -0.23 |
|  | Spline 4 | -0.01 | -0.49, 0.46 |
|  | Spline 5 | -0.69 | -1.86, 0.49 |

Models were adjusted for sex, socioeconomic position, depression symptoms and attention deficit hyperactivity disorder (ADHD) traits.

*Supplementary Table S33.* Exposure*age interactions and 50th percentile alcohol use

| **Exposure** |  | **Coefficient** | **CI 95% (lower, upper)** |
| --- | --- | --- | --- |
| Mean factor score for autism | Intercept | -8.72 | -28.08, 10.64 |
|  | Spline 1 | 0.82 | -0.33, 1.97 |
|  | Spline 2 | 1.18 | 0.70, 1.66 |
|  | Spline 3 | -0.61 | -1.01, -0.21 |
|  | Spline 4 | -0.0 | -0.39, 0.38 |
|  | Spline 5 | -0.98 | -1.89, -0.06 |
| Social Communication Disorders Checklist (SCDC) score at 7y | Intercept | -5.69 | -30.77, 19.39 |
|  | Spline 1 | 0.63 | -0.87, 2.12 |
|  | Spline 2 | 1.19 | 0.58, 1.80 |
|  | Spline 3 | -0.57 | -0.93, -0.22 |
|  | Spline 4 | -0.06 | -0.50, 0.37 |
|  | Spline 5 | -0.91 | -2.03, 0.21 |
| Polygenic score for autism | Intercept | -12.36 | -34.81, 10.10 |
|  | Spline 1 | 1.32 | -0.02, 2.65 |
|  | Spline 2 | 1.09 | 0.65, 1.53 |
|  | Spline 3 | -0.68 | -1.00, -0.36 |
|  | Spline 4 | 0.00 | -0.42, 0.42 |
|  | Spline 5 | -0.68 | -1.83, 0.46 |

Models were adjusted for sex, socioeconomic position, depression symptoms and attention deficit hyperactivity disorder (ADHD) traits.

*Supplementary Table S34.* Exposure*age interactions and 90th percentile alcohol use

| **Exposure** |  | **Coefficient** | **CI 95% (lower, upper)** |
| --- | --- | --- | --- |
| Mean factor score for autism | Intercept | -8.70 | -28.50, 11.11 |
|  | Spline 1 | 1.05 | -0.13, 2.22 |
|  | Spline 2 | 1.15 | 0.71, 1.59 |
|  | Spline 3 | -0.66 | -0.95, -0.36 |
|  | Spline 4 | -0.01 | -0.39, 0.36 |
|  | Spline 5 | -0.97 | -2.08, 0.13 |
| Social Communication Disorders Checklist (SCDC) score at 7y | Intercept | -5.67 | -24.15, 12.82 |
|  | Spline 1 | 0.88 | -0.23, 1.99 |
|  | Spline 2 | 1.19 | 0.75, 1.64 |
|  | Spline 3 | -0.60 | -0.92, -0.28 |
|  | Spline 4 | -0.06 | -0.48, 0.37 |
|  | Spline 5 | -0.91 | -2.13, 0.31 |
| Polygenic score for autism | Intercept | -12.36 | -34.81, 10.10 |
|  | Spline 1 | 1.32 | -0.02, 2.65 |
|  | Spline 2 | 1.09 | 0.65, 1.53 |
|  | Spline 3 | -0.68 | -1.00, -0.36 |
|  | Spline 4 | 0.00 | -0.42, 0.42 |
|  | Spline 5 | -0.68 | -1.83, 0.46 |

Models were adjusted for sex, socioeconomic position, depression symptoms and attention deficit hyperactivity disorder (ADHD) traits.

*Supplementary Table S35.* Exposure*age interactions and mean alcohol use (with depression excluded)

| **Exposure** |  | **Coefficient** | **CI 95% (lower, upper)** |
| --- | --- | --- | --- |
| Mean factor score for autism | Intercept | -5.74 | -22.57, 11.10 |
|  | Spline 1 | 0.69 | -0.31, 1.69 |
|  | Spline 2 | 1.15 | 0.76, 1.55 |
|  | Spline 3 | -0.64 | -0.94, -0.34 |
|  | Spline 4 | -0.12 | -0.45, 0.21 |
|  | Spline 5 | -0.73 | -1.61, 0.15 |
| Social Communication Disorders Checklist (SCDC) score at 7y | Intercept | -3.18 | -20.56, 14.20 |
|  | Spline 1 | 0.54 | -0.50, 1.57 |
|  | Spline 2 | 1.21 | 0.81, 1.62 |
|  | Spline 3 | -0.63 | -0.94, -0.33 |
|  | Spline 4 | -0.12 | -0.46, 0.22 |
|  | Spline 5 | -0.76 | -1.67, 0.16 |
| Polygenic score for autism | Intercept | -8.12 | -26.59, 10.36 |
|  | Spline 1 | 0.83 | -0.27, 1.93 |
|  | Spline 2 | 1.14 | 0.71, 1.58 |
|  | Spline 3 | -0.68 | -1.01, -0.35 |
|  | Spline 4 | -0.14 | -0.49, 0.22 |
|  | Spline 5 | -0.54 | -1.49, 0.41 |

Models were adjusted for sex, socioeconomic position and attention deficit hyperactivity disorder (ADHD) traits.

*Supplementary Table S36* Exposure*age interactions and 10th percentile alcohol use (with depression excluded)

| **Exposure** |  | **Coefficient** | **CI 95% (lower, upper)** |
| --- | --- | --- | --- |
| Mean factor score for autism | Intercept | -4.70 | -23.68, 14.29 |
|  | Spline 1 | 0.40 | -0.73, 1.53 |
|  | Spline 2 | 1.06 | 0.59, 1.52 |
|  | Spline 3 | -0.56 | -0.87, -0.24 |
|  | Spline 4 | 0.09 | -0.27, 0.46 |
|  | Spline 5 | -0.95 | -1.97, 0.08 |
| Social Communication Disorders Checklist (SCDC) score at 7y | Intercept | -3.50 | -24.02, 17.02 |
|  | Spline 1 | 0.30 | -0.93, 1.53 |
|  | Spline 2 | 1.09 | 0.57, 1.61 |
|  | Spline 3 | -0.54 | -0.87, -0.22 |
|  | Spline 4 | -0.02 | -0.46, 0.43 |
|  | Spline 5 | -0.85 | -1.96, 0.27 |
| Polygenic score for autism | Intercept | -7.95 | -28.32, 12.42 |
|  | Spline 1 | 0.59 | -0.63, 1.80 |
|  | Spline 2 | 1.01 | 0.60, 1.41 |
|  | Spline 3 | -0.59 | -0.93, -0.25 |
|  | Spline 4 | 0.03 | -0.38, 0.44 |
|  | Spline 5 | -0.67 | -1.75, 0.40 |

Models were adjusted for sex, socioeconomic position and attention deficit hyperactivity disorder (ADHD) traits.

*Supplementary Table S37.* Exposure*age interactions and 50th percentile alcohol use (with depression excluded)

| **Exposure** |  | **Coefficient** | **CI 95% (lower, upper)** |
| --- | --- | --- | --- |
| Mean factor score for autism | Intercept | -4.29 | -23.34, 17.76 |
|  | Spline 1 | 0.60 | -0.54, 1.74 |
|  | Spline 2 | 1.10 | 0.66, 1.54 |
|  | Spline 3 | -0.63 | -0.96, -0.29 |
|  | Spline 4 | -0.02 | -0.43, 0.39 |
|  | Spline 5 | -1.00 | -2.12, 0.12 |
| Social Communication Disorders Checklist (SCDC) score at 7y | Intercept | -3.49 | -24.84, 17.86 |
|  | Spline 1 | 0.54 | -0.72, 1.80 |
|  | Spline 2 | 1.13 | 0.64, 1.62 |
|  | Spline 3 | -0.59 | -0.96, -0.21 |
|  | Spline 4 | -0.06 | -0.45, 0.33 |
|  | Spline 5 | -0.89 | -2.01, 0.23 |
| Polygenic score for autism | Intercept | -7.93 | -27.96, 12.09 |
|  | Spline 1 | 0.81 | -0.39, 2.01 |
|  | Spline 2 | 1.04 | 0.56, 1.53 |
|  | Spline 3 | -0.64 | -1.00, -0.28 |
|  | Spline 4 | -0.05 | -0.50, 0.40 |
|  | Spline 5 | -0.72 | -1.88, 0.44 |

Models were adjusted for sex, socioeconomic position and attention deficit hyperactivity disorder (ADHD) traits.

*Supplementary Table S38.* Exposure*age interactions and 90th percentile alcohol use (with depression excluded)

| **Exposure** |  | **Coefficient** | **CI 95% (lower, upper)** |
| --- | --- | --- | --- |
| Mean factor score for autism | Intercept | -4.26 | -22.73, 14.21 |
|  | Spline 1 | 0.86 | -0.24, 1.97 |
|  | Spline 2 | 1.08 | 0.64, 1.52 |
|  | Spline 3 | -0.70 | -1.07, -0.33 |
|  | Spline 4 | -0.04 | -0.46, 0.37 |
|  | Spline 5 | -0.99 | -2.19, 0.21 |
| Social Communication Disorders Checklist (SCDC) score at 7y | Intercept | -3.46 | -20.41, 13.49 |
|  | Spline 1 | 0.81 | -0.19, 1.81 |
|  | Spline 2 | 1.09 | 0.63, 1.55 |
|  | Spline 3 | -0.65 | -0.95, -0.34 |
|  | Spline 4 | -0.07 | -0.37, 0.24 |
|  | Spline 5 | -0.88 | -1.75, -0.00 |
| Polygenic score for autism | Intercept | -7.91 | -26.35, 10.54 |
|  | Spline 1 | 1.07 | -0.03, 2.17 |
|  | Spline 2 | 1.03 | 0.52, 1.54 |
|  | Spline 3 | -0.73 | -1.09, -0.36 |
|  | Spline 4 | -0.07 | -0.52, 0.38 |
|  | Spline 5 | -0.71 | -1.88, 0.46 |

Models were adjusted for sex, socioeconomic position and attention deficit hyperactivity disorder (ADHD) traits.

*Supplementary Table S39.* Chi square comparison of sex between participants with base covariates up to the age of 17 years only vs. Those with base covariates and at least one outcome timepoint (up to the age of 28 years)

| **Sex** | **No outcome data** | **Has outcome data** | **Total** |
| --- | --- | --- | --- |
| Male | 6,263 | 1,300 | 7,563 |
| Female | 5,490 | 1,757 | 7,247 |
| **Total** | **11,753** | **3,057** | **14,810** |

*Pearson χ²(1) = 112.47, p = < 0.001*

*Supplementary Table S40.* Chi square comparison of maternal education between participants with base covariates up to the age of 17 years only vs. Those with base covariates and at least one outcome timepoint (up to the age of 28 years)

| **Mum's Highest Qualification** | **No outcome data** | **Has outcome data** | **Total** |
| --- | --- | --- | --- |
| CSE | 2,204 | 277 | 2,481 |
| Vocational | 1,025 | 187 | 1,212 |
| O level | 3,234 | 1,014 | 4,248 |
| A level | 1,860 | 891 | 2,751 |
| Degree | 891 | 688 | 1,579 |
| **Total** | **9,214** | **3,057** | **12,271** |

*Pearson χ**²(4) = 687.49, p < 0.001*

*Supplementary Table S41.* Chi square comparison of sex between participants with complete data for base covariates up to the age of 17 years only vs. Those with missing data in base covariates up to the age of 17 years only

| **Sex** | **Missing base data** | **Complete base data** | **Total** |
| --- | --- | --- | --- |
| Male | 6,222 | 1,341 | 7,563 |
| Female | 5,472 | 1,775 | 7,247 |
| **Total** | **11,694** | **3,116** | **14,810** |

*Pearson χ²(1) = 101.85, p = < 0.001*

*Supplementary Table S42.* Chi square comparison of maternal education between participants with complete data for base covariates up to the age of 17 years only vs. Those with missing data in base covariates up to the age of 17 years only

| **Mum's Highest Qualification** | **Missing base data** | **Complete base data** | **Total** |
| --- | --- | --- | --- |
| CSE | 2,193 | 288 | 2,481 |
| Vocational | 1,020 | 192 | 1,212 |
| O level | 3,215 | 1,033 | 4,248 |
| A level | 1,848 | 903 | 2,751 |
| Degree | 879 | 700 | 1,579 |
| **Total** | **9,155** | **3,116** | **12,271** |

*Pearson χ²(4) = 688.94, p < 0.001*

*Supplementary Table S43*. Completeness/missingness for each outcome timepoint in individuals who had observed data for all other substantive variables in each of the three samples.

| **Exposure** | **Outcome timepoint** | **N with data available** | **N with data missing** |
| --- | --- | --- | --- |
| Mean factor score for autism | 17 years | 3,672 | 475 |
|  | 19 years | 2,275 | 1,872 |
|  | 21years | 2,552 | 1,595 |
|  | 23 years | 2,459 | 1,688 |
|  | 28 years | 2,302 | 1,845 |
| SCDC score | 17 years | 3,426 | 449 |
|  | 19 years | 2,143 | 1,732 |
|  | 21years | 2,420 | 1,455 |
|  | 23 years | 2,329 | 1,546 |
|  | 28 years | 2,165 | 1,710 |
| Polygenic score for autism | 17 years | 2,972 | 344 |
|  | 19 years | 1,854 | 1,462 |
|  | 21years | 2,080 | 1,236 |
|  | 23 years | 2,015 | 1,301 |
|  | 28 years | 1,892 | 1,424 |

*Supplementary Table S44.* Descriptive statistics of the analytical sample where the SCDC score is the exposure (N=3,250).

| **Variable** | **N (%)** |
| --- | --- |
| **Sex** |  |
| *Males* | 1,587 (42.08) |
| *Females* | 2,184 (57.92) |
| **Maternal highest education 32 weeks' gestation** |  |
| *Up to A level* | 2,707 (71.79) |
| *Degree* | 818 (21.69) |
| *Vocational* | 246 (6.52) |
| **Maternal home ownership status** |  |
| *Mortgaged or owned* | 3,224 (87.00) |
| *Rented* | 482 (13.00) |
| **Parity** |  |
| *0* | 1,833 (49.39) |
| *1* | 1,294 (34.87) |
| *2* | 444 (11.96) |
| *3+* | 140 (3.78) |
| **Maternal occupational social class** |  |
| Professional occupation | 312 (9.34) |
| Managerial or technical occupation | 1,268 (37.95) |
| Skilled occupation (manual or non-manual) | 1,496 (44.78) |
| Partly skilled occupation, unskilled or armed forces | 265 (2.19) |
| **Maternal marital status** |  |
| Never married | 423 (11.33) |
| Divorced, separated or widowed | 159 (4.25) |
| First marriage | 2,914 (78.02) |
| Marriage two or three | 239 (6.40) |

*Supplementary Table S45.* Descriptive statistics of the analytical sample where the polygenic score for autism is the exposure (N=3,371)

| **Variable** | **N (%)** |
| --- | --- |
| **Sex** |  |
| *Males* | 1,364 (41.97) |
| *Females* | 1,886 (58.03) |
| **Maternal highest education 32 weeks' gestation** |  |
| *Up to A level* | 2,335 (71.85) |
| *Degree* | 710 (21.85) |
| *Vocational* | 205 (6.31) |
| **Maternal home ownership status** |  |
| *Mortgaged or owned* | 2,804 (87.46) |
| *Rented* | 402 (12.54) |
| **Parity** |  |
| *0* | 1,544 (48.19) |
| *1* | 1,162 (36.27) |
| *2* | 383 (11.95) |
| *3+* | 115 (3.59) |
| **Maternal occupational social class** |  |
| Professional occupation | 253 (8.78) |
| Managerial or technical occupation | 1,113 (38.65) |
| Skilled occupation (manual or non-manual) | 1,276 (44.31) |
| Partly skilled occupation, unskilled or armed forces | 238 (8.27) |
| **Maternal marital status** |  |
| Never married | 343 (10.64) |
| Divorced, separated or widowed | 144 (4.45) |
| First marriage | 2,533 (78.54) |
| Marriage two or three | 203 (6.36) |
